## Supplementary Figures for "Detailed disease progression of 213 patients hospitalized with Covid-19 in the Czech Republic: An exploratory analysis"

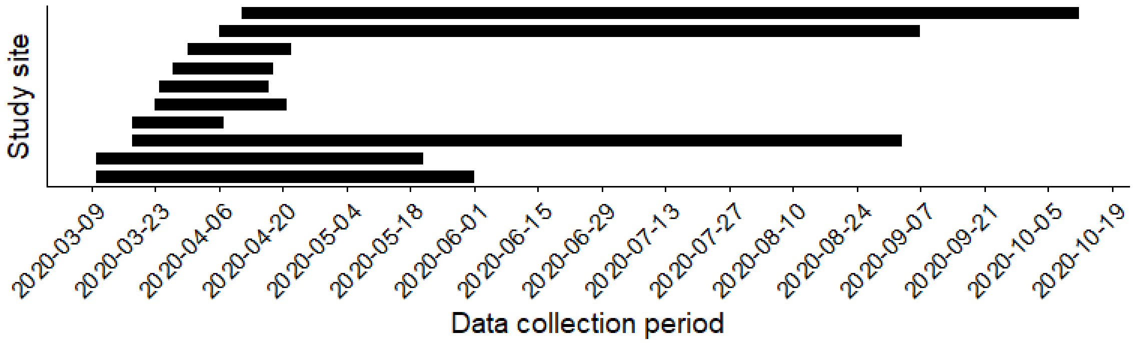


*Figure 1: Data collection periods at individual sites, showing the range of admission dates of patients included in the study. Note that we cannot provide additional information to link the sites here with data shown elsewhere as that would increase the risk of deanonymization of the patients.*

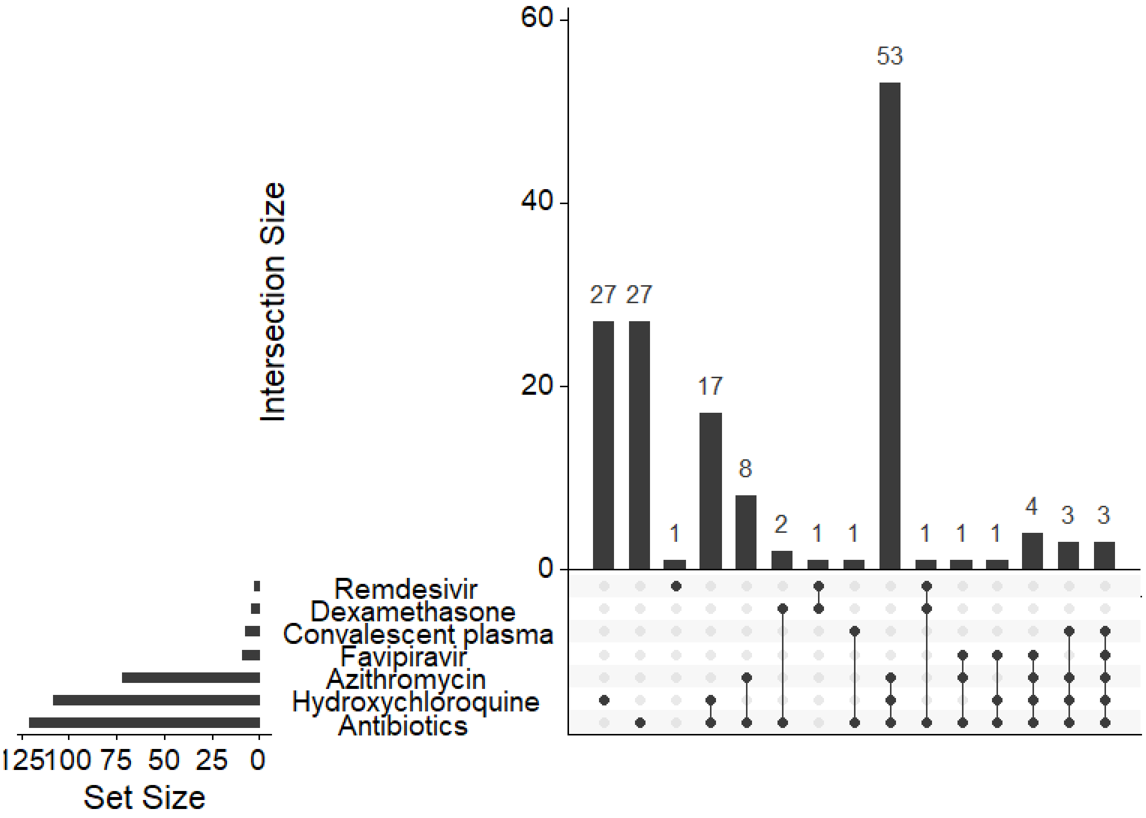


*Figure 2: Upset plot of treatment combinations - each vertical bar displays the number of patients that received the combination indicated by filled dots in the matrix. Horizontal bars show the total number of patients receiveing the given treatment.*

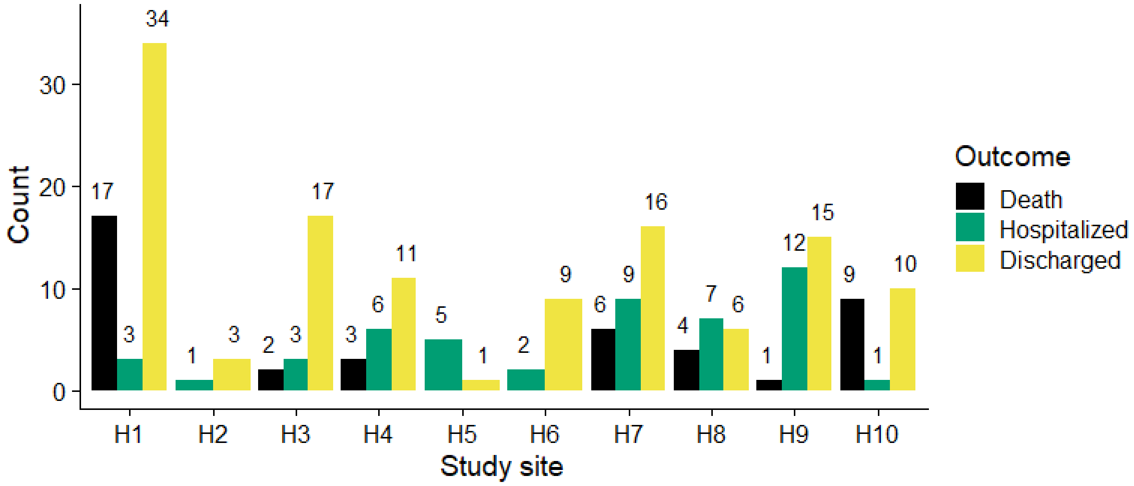


*Figure 3: Number of patients and outcomes at the individual sites. The numbers above bars are the exact counts. Hospitalized = still hospitalized at the end of data collection at the site or transferred to other site and lost to followup. Sites are anonymized to preserve patient privacy.*

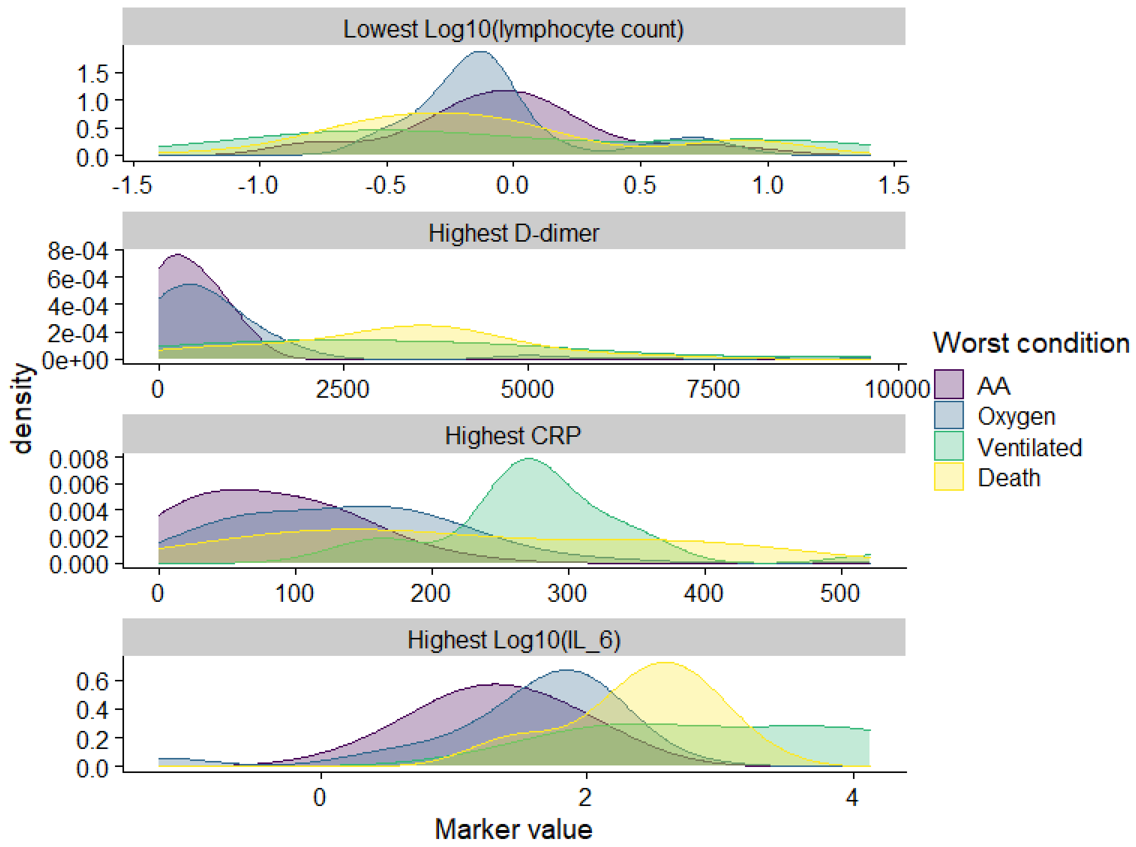


*Figure 4: Density plots of worst marker values per patient, stratified by worst condition experienced by the patient. For each patient that had a given marker measured, the worst value was taken. Additionally the patients are classified by the worst condition (regardless of the timing relative to the worst marker levels). For each set of patients and marker an empirical density plot of the worst marker values is shown.*

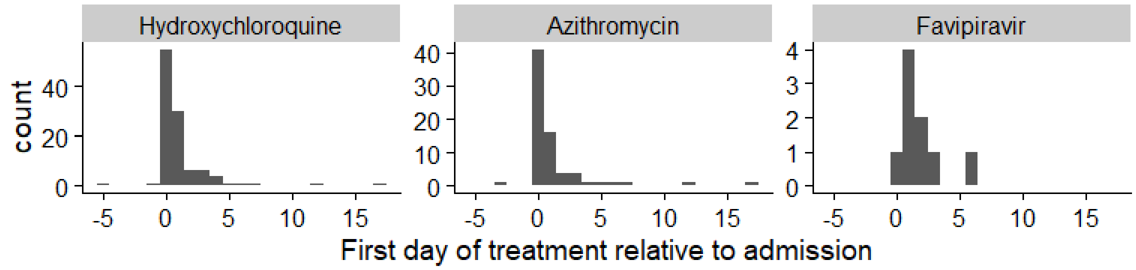


*Figure 5: Histogram of timing of first treatment relative to admission into one of the study sites. Two patients initiated treatment before admission, which is shown as the negative numbers.*

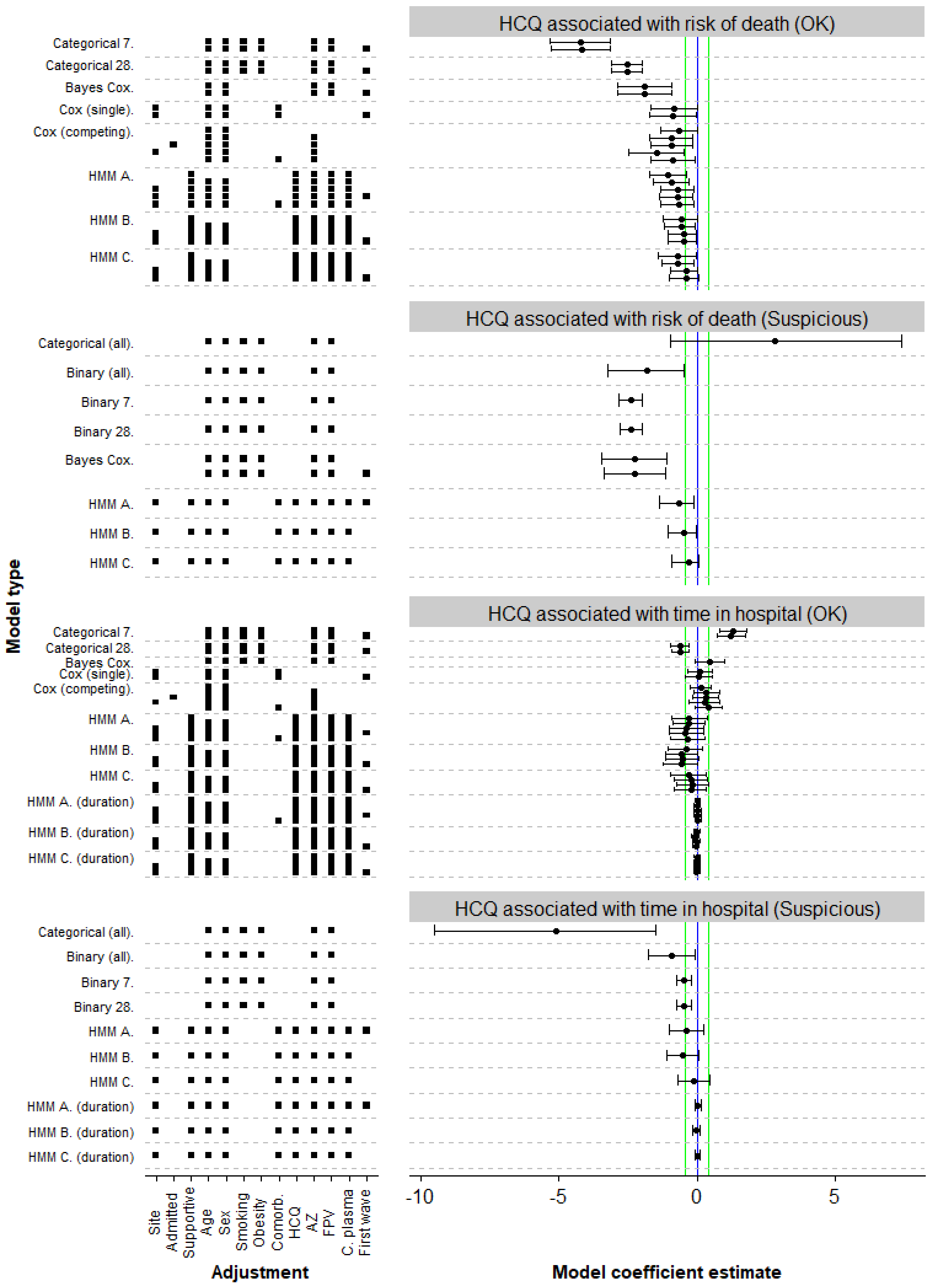


*Figure 6: Estimates of model coefficients for association between hydroxychloroquine and main outcomes. The "Suspicious" section shows models that were found to not fit the data well or have computational issues - see supplementary statistical analysis for*

*details. Each row represents a model - Categorical All/7/28 = Bayesian categorical regression for state at last observed day/day 7/day 28, Binary All/7/28 = Bayesian logistic regression for state at last observed day/day 7/day 28, Bayes Cox = Bayesian version of the Cox proportional hazards model with a binary outcome, Cox (single) = frequentist Cox model with a binary outcome, Cox (competing) = frequentist Cox model using competing risks (as in Figure 1a), HMM A = Bayesian hidden-Markov model as in Figure 1b with predictors for rate groups, HMM B = Bayesian hidden-markov model as in Figure 1b with predictors for individual rates, HMM C = Bayesian hidden-Markov model as in Figure 1c. For frequentist models, we show maximum likelihood estimate and 95% confidence interval, for Bayesian models we show posterior mean and 95% credible interval. The estimands are either log odds-ratio (Categorical, HMM) or log hazard ratio (Cox variants) or log ratio of mean duration of hospitalization (HMM duration). In all cases coefficient < 0 means better patient outcome in the treatment group. Vertical lines indicate zero (blue) and substantial increase or decrease with odds or hazard ratio of 3:2 or 2:3 (green). Additionally the factors the model adjusted for are listed - Site = the study site, admitted = Admitted for Covid-19, Supportive = best supportive care initiated, Comorb. = total number of comorbidities, AZ = took azithromycin, HCQ = took hydroxychloroquine, FPV = took favipiravir, C. plasma = received convalescent plasma, first wave = only patients admitted before September 1st were included.*

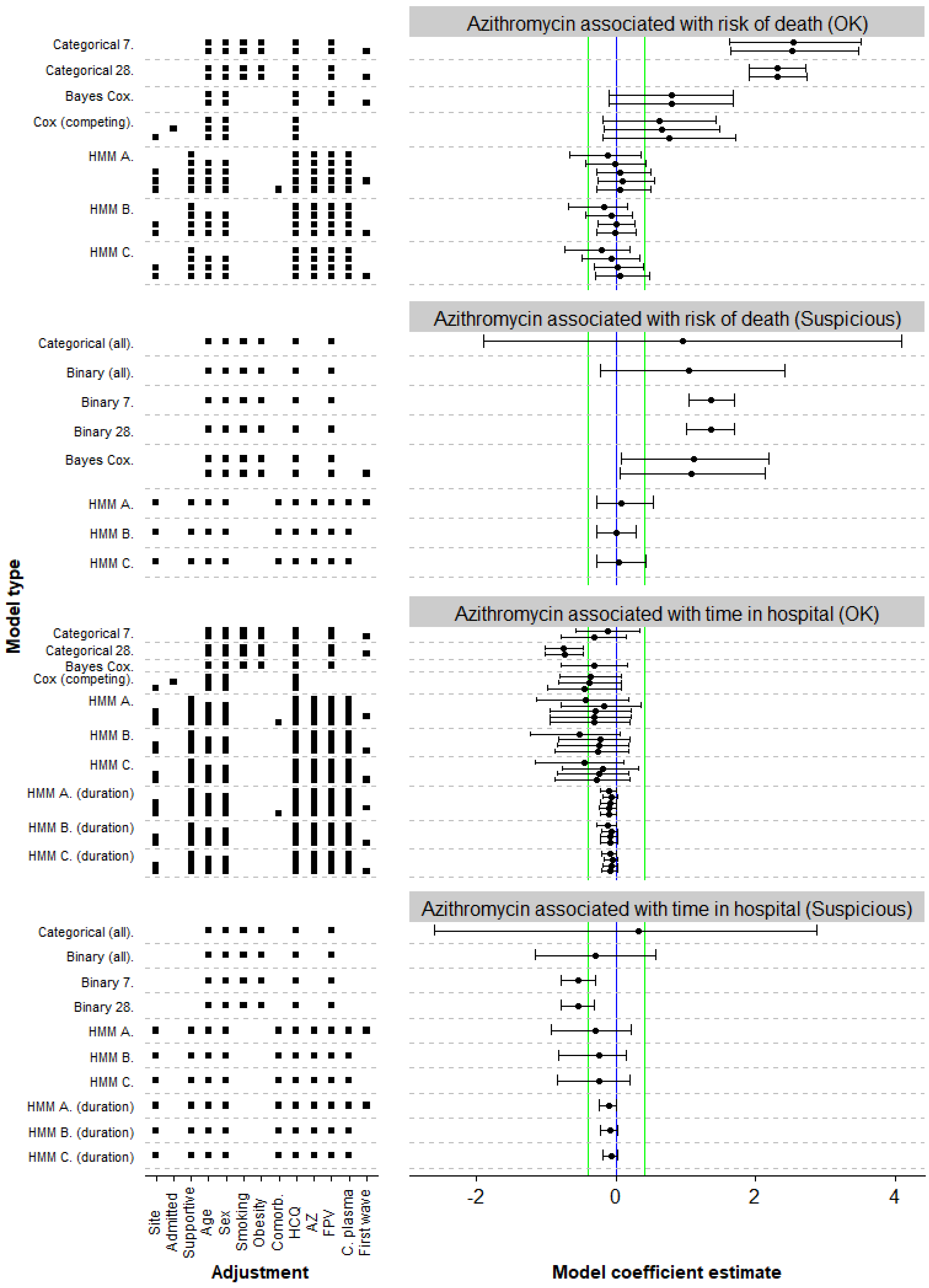


*Figure 7: Estimates of model coefficients for association between azithromycin and main outcomes. The "Suspicious" section shows models that were found to not fit the data well or have computational issues - see supplementary statistical analysis for*

*details. Each row represents a model - Categorical All/7/28 = Bayesian categorical regression for state at last observed day/day 7/day 28, Binary All/7/28 = Bayesian logistic regression for state at last observed day/day 7/day 28, Bayes Cox = Bayesian version of the Cox proportional hazards model with a binary outcome, Cox (single) = frequentist Cox model with a binary outcome, Cox (competing) = frequentist Cox model using competing risks (as in Figure 1a), HMM A = Bayesian hidden-Markov model as in Figure 1b with predictors for rate groups, HMM B = Bayesian hidden-markov model as in Figure 1b with predictors for individual rates, HMM C = Bayesian hidden-Markov model as in Figure 1c. For frequentist models, we show maximum likelihood estimate and 95% confidence interval, for Bayesian models we show posterior mean and 95% credible interval. The estimands are either log odds-ratio (Categorical, HMM) or log hazard ratio (Cox variants) or log ratio of mean duration of hospitalization (HMM duration). In all cases coefficient < 0 means better patient outcome in the treatment group. Vertical lines indicate zero (blue) and substantial increase or decrease with odds or hazard ratio of 3:2 or 2:3 (green). Additionally the factors the model adjusted for are listed - Site = the study site, admitted = Admitted for Covid-19, Supportive = best supportive care initiated, Comorb. = total number of comorbidities, AZ = took azithromycin, HCQ = took hydroxychloroquine, FPV = took favipiravir, C. plasma = received convalescent plasma, first wave = only patients admitted before September 1st were included.*

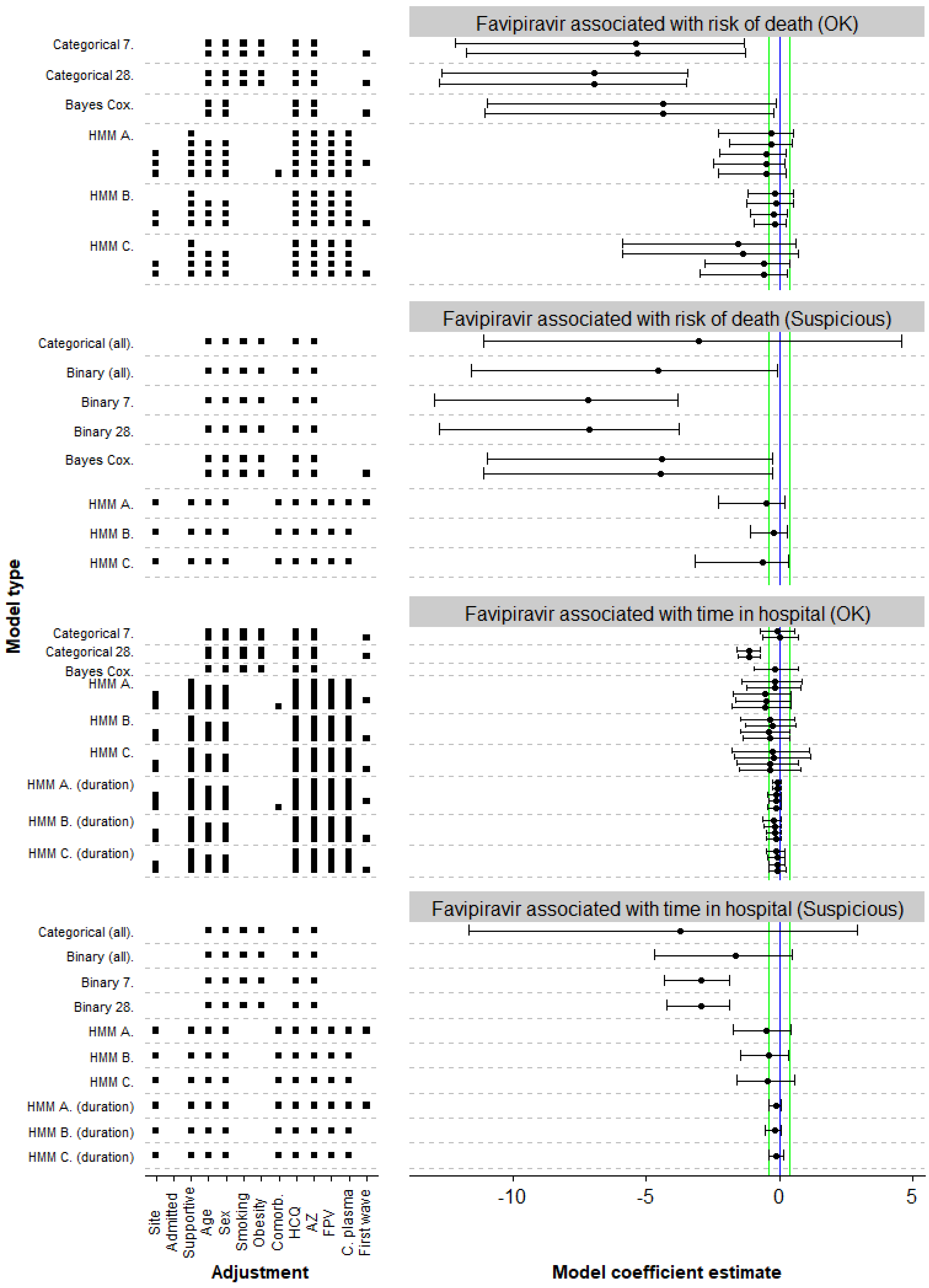


*Figure 8: Estimates of model coefficients for association between favipiravir and main outcomes. The "Suspicious" section shows models that were found to not fit the data well or have computational issues - see supplementary statistical analysis for details.*

*Each row represents a model - Categorical All/7/28 = Bayesian categorical regression for state at last observed day/day 7/day 28, Binary All/7/28 = Bayesian logistic regression for state at last observed day/day 7/day 28, Bayes Cox = Bayesian version of the Cox proportional hazards model with a binary outcome, Cox (single) = frequentist Cox model with a binary outcome, Cox (competing) = frequentist Cox model using competing risks (as in Figure 1a), HMM A = Bayesian hidden-Markov model as in Figure 1b with predictors for rate groups, HMM B = Bayesian hidden-markov model as in Figure 1b with predictors for individual rates, HMM C = Bayesian hidden-Markov model as in Figure 1c. For frequentist models, we show maximum likelihood estimate and 95% confidence interval, for Bayesian models we show posterior mean and 95% credible interval. The estimands are either log odds-ratio (Categorical, HMM) or log hazard ratio (Cox variants) or log ratio of mean duration of hospitalization (HMM duration). In all cases coefficient < 0 means better patient outcome in the treatment group. Vertical lines indicate zero (blue) and substantial increase or decrease with odds or hazard ratio of 3:2 or 2:3 (green). Additionally the factors the model adjusted for are listed - Site = the study site, admitted = Admitted for Covid-19, Supportive = best supportive care initiated, Comorb. = total number of comorbidities, AZ = took azithromycin, HCQ = took hydroxychloroquine, FPV = took favipiravir, C. plasma = received convalescent plasma, first wave = only patients admitted before September 1st were included.*

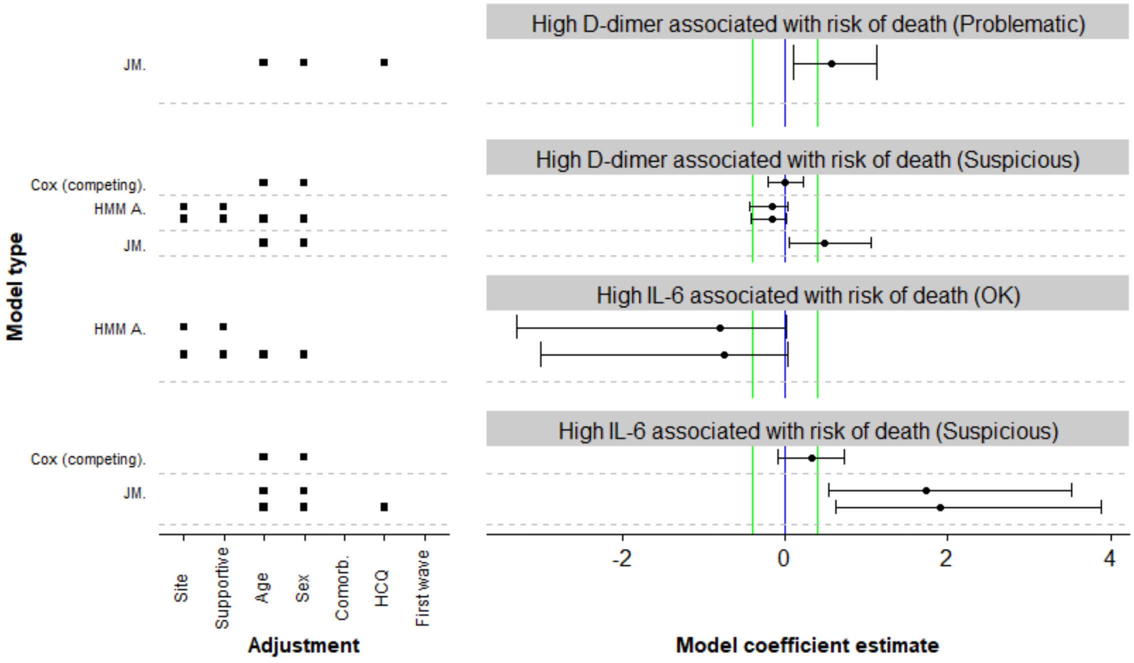


*Figure 9: Estimates of model coefficients (log hazard ratios) for association between markers and death. The "Suspicious" section shows models that were found to not fit the data well or have computational issues, "Problematic" section shows models that were completely broken - see supplementary statistical analysis for details. Each row represents a model - Cox (competing) = frequentist Cox model using competing risks (as in Figure 1a), HMM A = Bayesian hidden-markov model as in Figure 1b with*

*predictors for rate groups, JM = Bayesian joint longitudinal and time-to-event model. For frequentist models, we show maximum likelihood estimate and 95% confidence interval, for Bayesian models we show posterior mean and 95% credible interval. Additionally the factors the model adjusted for are listed - Site = the study site, Supportive = best supportive care initiated, HCQ = took Hydroxychloroquine. We show posterior mean and 95% credible interval.*
