## Supplementary Statistical Analysis for "Detailed disease progression of 213 patients hospitalized with Covid-19 in the Czech Republic: An exploratory analysis"

Version: 2020-12-03

**Abstract**

This documents describes additional details about the models and statistical analyses used in the paper, including fitted coefficient values. Additionally, details on the risk scores and how we evaluated them are provided. Chapters of the document correspond to individual model families we used, final chapter is devoted to risk scores evaluation. Overall description of the models and the outputs presented is given at the beginning of each chapter and is not repeated for each model. To avoid any ambiguities, complete code to compile this document and for all analyses is available at https://github.com/cas-bioinf/covid19retrospective/.

**8 Original computing environment** **84**

**References** **85**

### 1 General remarks on models

For all treatments under investigation, we try to quantify their association with both mortality and length of hospitalization. The exact ways those are represented differs between models.

We sort models into 3 groups:

- "Problematic" - model is almost certainly not useful, especially because of computational issues.
- "Suspicious" - model has some problems representing the data or minor computational issues. It is likely the results are not completely trustworthy, but they are also unlikely to be completely useless.
- "OK" - we found no problems with the model.

In the main text, we report only "OK" models, but supplementary figures show the multiverse analysis performed for all models we tried.

To make sure our inferences are not skewed by including patients from both first and second waves of the pandemic, we run most analyses twice: once with those patients included and once without them.

#### 1.1 Comorbidity score

Multiple models share the same comorbidity score that is computed as the count of the following 9 conditions that are met (missing data are treated as 0):

- Heart problems (NYHA > 1 OR ischemic heart disease OR heart failur)
- Hypertension (uses at least 1 hypertension drug)
- Lung problems (COPD OR asthma OR other lung disease)
- Diabetes
- Renal disease
- Liver disease
- Smoking
- Markers out of range (creatinine > 115 for males, creatinine > 97 for females OR PT INR > 1.2 OR albumin < 36)
- Obesity (as ascertained by a clinician OR BMI > 30)

All of those quantities were evaluated upon admission. See the protocol of the study for more details on the definitions.

### 2 Single outcome proprtional hazards model

Here we use a single outcome survival model via the `coxph` function of the `Survival` package. Hydroxychloroquine is treated as a time-varying covariate, other covariates are fixed.

#### 2.1 Risk of death, increased discharge

Here, we compare the model with hydroxychloroquine as a covariate (`m1`) with model not including hydroxychloroquine (`m0`), the formulas used are:

```
m1: Surv(t0,t1,outcome=='Death')~hcq+age+sex+strata(hospital_id)+comorb_sum)
m0: Surv(t0,t1,outcome=='Death')~    age+sex+strata(hospital_id)+comorb_sum)
```

where `comorb_sum` is a score representing the cumulative presence of ischemic heart disease, hypertension drugs, former heart failure, COPD, other lung diseases, renal disease, or high creatinine. A separate model is fitted for the "Discharged" outcome.

The point estimate of the effect and its confidence intervals are taken directly from the `m1` model, a p-value is computed via a call `anova` comparing the two models.

Here are the fitted models for the "Death" outcome:

```
## Call:
## coxph(formula = S <- Surv(t0, t1, outcome == "Death") ~ hcq +
##     age + sex + strata(hospital_id) + comorb_sum)
##
##                coef exp(coef) se(coef)      z       p
## hcq        -0.80853   0.44551  0.43076 -1.877 0.06052
## age         0.05295   1.05438  0.01651  3.207 0.00134
## sexM       -0.09150   0.91257  0.35731 -0.256 0.79790
## comorb_sum  0.20826   1.23153  0.13542  1.538 0.12408
##
## Likelihood ratio test=35.46  on 4 df, p=3.731e-07
## n= 231, number of events= 42
```

```
## Call:
## coxph(formula = S <- Surv(t0, t1, outcome == "Death") ~ age +
##     sex + strata(hospital_id) + comorb_sum)
##
##               coef exp(coef) se(coef)     z        p
## age        0.05703   1.05868  0.01653 3.449 0.000563
## sexM       0.06408   1.06617  0.34518 0.186 0.852733
## comorb_sum 0.27097   1.31123  0.12639 2.144 0.032038
##
## Likelihood ratio test=31.91  on 3 df, p=5.471e-07
## n= 231, number of events= 42
```

And the fitted models for the "Discharged" outcome:

```
## Call:
## coxph(formula = S <- Surv(t0, t1, outcome == "Discharged") ~
##     hcq + age + sex + strata(hospital_id) + comorb_sum)
```

```
##
##                 coef exp(coef)  se(coef)      z       p
## hcq        -0.111633  0.894372  0.235877 -0.473 0.63602
## age        -0.026860  0.973498  0.006946 -3.867 0.00011
## sexM       -0.381516  0.682826  0.212596 -1.795 0.07272
## comorb_sum -0.191992  0.825314  0.110851 -1.732 0.08328
##
## Likelihood ratio test=31.69  on 4 df, p=2.218e-06
## n= 231, number of events= 122
```

```
## Call:
## coxph(formula = S <- Surv(t0, t1, outcome == "Discharged") ~
##     age + sex + strata(hospital_id) + comorb_sum)
##
##                 coef exp(coef)  se(coef)      z        p
## age        -0.027171  0.973194  0.006958 -3.905 9.41e-05
## sexM       -0.373993  0.687982  0.212417 -1.761   0.0783
## comorb_sum -0.180325  0.834999  0.107669 -1.675   0.0940
##
## Likelihood ratio test=31.46  on 3 df, p=6.79e-07
## n= 231, number of events= 122
```

And here are the resulting evaluation of individual hypotheses - note that we change sign of the coefficient for the "length of hospitalization" hypothesis (modeled here as the risk of discharge) to conform to the overall rule that negative coefficient implies better outcomes for treated patients:

```
##        hypothesis point_estimate    p_value     ci_low    ci_high
## hcq     hcq_death     -0.8085289 0.05937969 -1.6527965 0.03573871
## hcq1 hcq_hospital      0.1116331 0.63722685 -0.3506773 0.57394344
```

#### 2.2 Risk of death, increased discharge - first wave only

Here, we perform the same analysis as above, but including only patients from the first wave, here are the fitted models for the "Death" outcome:

```
## Call:
## coxph(formula = S <- Surv(t0, t1, outcome == "Death") ~ hcq +
##     age + sex + strata(hospital_id) + comorb_sum)
##
##                coef exp(coef) se(coef)      z       p
## hcq        -0.87891   0.41524  0.43822 -2.006 0.04490
## age         0.05181   1.05317  0.01638  3.162 0.00156
## sexM       -0.06256   0.93935  0.36108 -0.173 0.86244
## comorb_sum  0.16177   1.17559  0.13977  1.157 0.24708
##
## Likelihood ratio test=33.05  on 4 df, p=1.167e-06
## n= 209, number of events= 41
```

```
## Call:
## coxph(formula = S <- Surv(t0, t1, outcome == "Death") ~ age +
##     sex + strata(hospital_id) + comorb_sum)
##
```

```
##                coef exp(coef) se(coef)     z        p
## age        0.05622   1.05783  0.01639 3.430 0.000603
## sexM       0.10528   1.11102  0.34833 0.302 0.762468
## comorb_sum 0.23554   1.26560  0.12931 1.822 0.068526
##
## Likelihood ratio test=28.96  on 3 df, p=2.279e-06
## n= 209, number of events= 41
```

And here are the fits for the "Discharged" outcome:

```
## Call:
## coxph(formula = S <- Surv(t0, t1, outcome == "Discharged") ~
##     hcq + age + sex + strata(hospital_id) + comorb_sum)
##
##                  coef exp(coef)  se(coef)      z       p
## hcq        -0.073062  0.929544  0.243409 -0.300 0.76406
## age        -0.024585  0.975715  0.007145 -3.441 0.00058
## sexM       -0.284091  0.752698  0.220526 -1.288 0.19766
## comorb_sum -0.222501  0.800515  0.115257 -1.930 0.05355
##
## Likelihood ratio test=28.68  on 4 df, p=9.074e-06
## n= 209, number of events= 115
```

```
## Call:
## coxph(formula = S <- Surv(t0, t1, outcome == "Discharged") ~
##     age + sex + strata(hospital_id) + comorb_sum)
##
##                 coef exp(coef) se(coef)      z        p
## age        -0.02472   0.97559  0.00716 -3.452 0.000557
## sexM       -0.27768   0.75754  0.21975 -1.264 0.206371
## comorb_sum -0.21484   0.80667  0.11216 -1.916 0.055427
##
## Likelihood ratio test=28.59  on 3 df, p=2.729e-06
## n= 209, number of events= 115
```

Which give us the following evaluation of the hypotheses:

```
##        hypothesis point_estimate    p_value     ci_low     ci_high
## hcq     hcq_death    -0.87890608 0.04322966 -1.7378092 -0.02000296
## hcq1 hcq_hospital     0.07306167 0.76457321 -0.4040118  0.55013509
```

### 3 Competing risks proportional hazards models

In all of those models we use the coxph function from the Survival package and treat death and discharged as competing risks. The status column in the data corresponds to the three-valued state "Hospitalized" (1) / "Discharged" (2) / "Death" (3). I.e. in all model outputs below, there are two transitions "1:2" is the transition to "Discharged" and "1:3" is the transition to "Death". We evaluate the hypotheses about treatment effects or marker associations directly via the estimated log hazard ratio of the treatment for the relevant transition. Notably, the sign of the effect on the transition to "Discharged" is flipped for the multiverse analysis because it assumes that positive effect means increased hospital stay.

We do not work with models with more than about 5 predictors, as those become impossible to estimate due to insufficient amount of events.

#### 3.1 Treatment effects

##### 3.1.1 Age + sex + hydroxychloroquine

The model uses the formula:

```
## Surv(daym1, day, status) ~ age + sex + took_hcq
```

Fitted coefficients below:

```
## Call:
## coxph(formula = f_competing_hcq, data = data_surv, id = patient_id)
##
##
## 1:2                 coef exp(coef)  se(coef)      z        p
##   age          -0.033093  0.967448  0.005576 -5.935 2.94e-09
##   sexM         -0.619574  0.538173  0.192141 -3.225  0.00126
##   took_hcqTRUE -0.134780  0.873908  0.192675 -0.700  0.48423
##
##
## 1:3                coef exp(coef) se(coef)      z        p
##   age           0.06784   1.07019  0.01536  4.416 1.01e-05
##   sexM          0.20432   1.22669  0.32281  0.633   0.5268
##   took_hcqTRUE -0.64734   0.52344  0.33379 -1.939   0.0525
##
##  States:  1= (s0), 2= Discharged, 3= Death
##
## Likelihood ratio test=72.12  on 6 df, p=1.504e-13
## n= 2856, number of events= 164
```

##### 3.1.2 Age + sex + hydroxychloroquine + azithromycin

The formula becomes:

```
## Surv(daym1, day, status) ~ age + sex + took_hcq + took_az
```

The fitted coefficients are:

```
## Call:
## coxph(formula = f_competing_hcq_az, data = data_surv, id = patient_id)
##
##
## 1:2                 coef exp(coef)  se(coef)      z        p
##   age          -0.032566  0.967959  0.005594 -5.821 5.84e-09
##   sexM         -0.630591  0.532277  0.192395 -3.278  0.00105
##   took_hcqTRUE -0.340387  0.711495  0.235689 -1.444  0.14868
##   took_azTRUE   0.362918  1.437518  0.227127  1.598  0.11007
##
##
## 1:3               coef exp(coef) se(coef)      z        p
##   age           0.07070   1.07326  0.01581  4.472 7.75e-06
##   sexM          0.20152   1.22326  0.32247  0.625   0.5320
##   took_hcqTRUE -0.92810   0.39530  0.39396 -2.356   0.0185
##   took_azTRUE   0.62289   1.86431  0.41440  1.503   0.1328
##
##  States:  1= (s0), 2= Discharged, 3= Death
##
## Likelihood ratio test=76.87  on 8 df, p=2.078e-13
## n= 2856, number of events= 164
```

###### 3.1.3 Age + sex + hydroxychloroquine + azithromycin + favipiravir

The formula is:

```
## Surv(daym1, day, status) ~ age + sex + took_hcq + took_az + took_favipiravir
```

And the fitted model is:

```
## Warning in fitter(X, Y, istrat, offset, init, control, weights = weights, :
## Loglik converged before variable 10 ; beta may be infinite.
```

```
## Call:
## coxph(formula = f_competing_hcq_az_fpv, data = data_surv, id = patient_id)
##
##
## 1:2                         coef exp(coef)  se(coef)      z        p
##   age                  -0.032595  0.967931  0.005601 -5.819 5.92e-09
##   sexM                 -0.633196  0.530892  0.193360 -3.275  0.00106
##   took_hcqTRUE         -0.341346  0.710813  0.235828 -1.447  0.14778
##   took_azTRUE           0.356086  1.427730  0.232984  1.528  0.12642
##   took_favipiravirTRUE  0.053290  1.054735  0.396045  0.135  0.89296
##
##
## 1:3                         coef  exp(coef)   se(coef)      z        p
##   age                  6.859e-02  1.071e+00  1.566e-02  4.379 1.19e-05
##   sexM                 2.223e-01  1.249e+00  3.216e-01  0.691    0.489
##   took_hcqTRUE        -9.002e-01  4.065e-01  3.899e-01 -2.309    0.021
##   took_azTRUE          6.712e-01  1.957e+00  4.099e-01  1.638    0.102
##   took_favipiravirTRUE -1.560e+01  1.681e-07  2.387e+03 -0.007    0.995
##
```

```
##  States:  1= (s0), 2= Discharged, 3= Death
##
## Likelihood ratio test=78.86  on 10 df, p=8.41e-13
## n= 2856, number of events= 164
```

###### 3.1.4 Age + sex + hydroxychloroquine + azithromycin + admitted

The formula is:

```
## Surv(daym1, day, status) ~ age + sex + took_hcq + took_az + admitted_for_covid
```

The fitted coefficients are:

```
## Call:
## coxph(formula = f_competing_hcq_az_admitted, data = data_surv,
##     id = patient_id)
##
##
## 1:2                            coef exp(coef)  se(coef)      z        p
##   age                     -0.032168  0.968344  0.005645 -5.698 1.21e-08
##   sexM                    -0.633182  0.530900  0.192108 -3.296 0.000981
##   took_hcqTRUE            -0.316517  0.728683  0.242984 -1.303 0.192703
##   took_azTRUE              0.374586  1.454389  0.229305  1.634 0.102349
##   admitted_for_covidTRUE -0.108702  0.896997  0.260350 -0.418 0.676295
##
##
## 1:3                           coef exp(coef) se(coef)      z        p
##   age                      0.07164   1.07427  0.01600  4.476 7.59e-06
##   sexM                     0.19173   1.21135  0.32272  0.594   0.5524
##   took_hcqTRUE            -0.91491   0.40055  0.39714 -2.304   0.0212
##   took_azTRUE              0.65309   1.92147  0.42041  1.553   0.1203
##   admitted_for_covidTRUE -0.27471   0.75979  0.38314 -0.717   0.4734
##
##  States:  1= (s0), 2= Discharged, 3= Death
##
## Likelihood ratio test=77.53  on 10 df, p=1.526e-12
## n= 2856, number of events= 164
```

###### 3.1.5 Age + sex + hydroxychloroquine + azithromycin + hospital

The new formula is:

```
## Surv(daym1, day, status) ~ age + sex + took_hcq + took_az + strata(hospital_id)
```

The resulting coefficients are:

```
## Call:
## coxph(formula = f_competing_hcq_az_hospital, data = data_surv_hospital,
##     id = patient_id)
##
##
```

```
## 1:2               coef exp(coef) se(coef)      z        p
##   age          -0.03107   0.96940  0.00622 -4.996 5.86e-07
##   sexM         -0.51804   0.59569  0.20993 -2.468   0.0136
##   took_hcqTRUE -0.28205   0.75424  0.28446 -0.992   0.3214
##   took_azTRUE   0.44901   1.56676  0.26655  1.685   0.0921
##
##
## 1:3               coef exp(coef) se(coef)      z        p
##   age           0.06824   1.07062  0.01672  4.081 4.49e-05
##   sexM         -0.01159   0.98848  0.35920 -0.032  0.97426
##   took_hcqTRUE -1.47076   0.22975  0.51683 -2.846  0.00443
##   took_azTRUE   0.75845   2.13496  0.48737  1.556  0.11966
##
##  States:  1= (s0), 2= Discharged, 3= Death
##
## Likelihood ratio test=69.91  on 8 df, p=5.123e-12
## n= 2856, number of events= 164
```

##### 3.1.6 Age + sex + hydroxychloroquine + azithromycin + comorbidities

The formula becomes:

```
## Surv(daym1, day, status) ~ age + sex + took_hcq + took_az + comorbidities_sum
```

Model fit below:

```
## Call:
## coxph(formula = f_competing_hcq_az_comorbidities_sum, data = data_surv,
##     id = patient_id)
##
##
## 1:2                     coef exp(coef)  se(coef)      z        p
##   age               -0.026956  0.973404  0.005915 -4.557 5.18e-06
##   sexM              -0.578760  0.560593  0.192666 -3.004  0.00266
##   took_hcqTRUE      -0.419492  0.657381  0.240529 -1.744  0.08115
##   took_azTRUE        0.310106  1.363570  0.231478  1.340  0.18035
##   comorbidities_sum -0.156459  0.855166  0.060952 -2.567  0.01026
##
##
## 1:3                    coef exp(coef) se(coef)      z       p
##   age                0.07084   1.07341  0.01599  4.431 9.4e-06
##   sexM               0.18351   1.20143  0.32608  0.563  0.5736
##   took_hcqTRUE      -0.88587   0.41236  0.40874 -2.167  0.0302
##   took_azTRUE        0.60279   1.82722  0.41527  1.452  0.1466
##   comorbidities_sum  0.03840   1.03914  0.10676  0.360  0.7191
##
##  States:  1= (s0), 2= Discharged, 3= Death
##
## Likelihood ratio test=83.67  on 10 df, p=9.547e-14
## n= 2856, number of events= 164
```

#### 3.2 Marker effects

##### 3.2.1 Age + sex + IL-6

The formula is:

```
## Surv(daym1, day, status) ~ age + sex + log_peak_IL_6
```

where `log_peak_IL_6` is the logarithm of the peak IL-6 value seen up to the given time. The fitted model is:

```
## Warning in fitter(X, Y, istrat, offset, init, control, weights = weights, :
## Loglik converged before variable 3 ; beta may be infinite.
```

```
## Call:
## coxph(formula = f_competing_il_6, data = data_IL_6, control = coxph.control(iter.max = 200),
##     id = patient_id)
##
##
## 1:2                   coef  exp(coef)   se(coef)      z        p
##   age           -4.948e-02  9.517e-01  1.267e-02 -3.906 9.38e-05
##   sexM          -5.280e-01  5.898e-01  3.917e-01 -1.348    0.178
##   log_peak_IL_6 -1.955e+01  3.243e-09  6.621e+03 -0.003    0.998
##
##
## 1:3                coef exp(coef) se(coef)     z      p
##   age           0.06808   1.07045  0.03417 1.992 0.0463
##   sexM          0.78070   2.18301  0.79933 0.977 0.3287
##   log_peak_IL_6 0.31954   1.37649  0.20799 1.536 0.1245
##
##  States:  1= (s0), 2= Discharged, 3= Death
##
## Likelihood ratio test=34.56  on 6 df, p=5.251e-06
## n= 960, number of events= 38
```

Note the warning on convergence and the huge `se` for the `log_peak_IL_6` coefficient for the 1:2 transition. We mark this model as "Suspicious".

##### 3.2.2 Age + sex + D-dimer

The formula is:

```
## Surv(daym1, day, status) ~ age + sex + log_peak_d_dimer
```

where `log_peak_d_dimer` is the logarithm of the peak D-dimer value seen up to the given time. The fitted model is:

```
## Warning in fitter(X, Y, istrat, offset, init, control, weights = weights, :
## Loglik converged before variable 3 ; beta may be infinite.
```

```
## Call:
## coxph(formula = f_competing_d_dimer, data = data_d_dimer, control = coxph.control(iter.max = 2000),
##     id = patient_id)
##
##
## 1:2                       coef  exp(coef)   se(coef)      z        p
##   age              -3.068e-02  9.698e-01  8.196e-03 -3.743 0.000182
##   sexM             -5.196e-01  5.948e-01  2.678e-01 -1.940 0.052369
##   log_peak_d_dimer -7.246e+01  3.391e-32  2.110e+04 -0.003 0.997260
##
##
## 1:3                     coef exp(coef) se(coef)     z      p
##   age              0.059677  1.061494 0.024095 2.477 0.0133
##   sexM             0.729760  2.074582 0.582730 1.252 0.2105
##   log_peak_d_dimer 0.003706  1.003713 0.109288 0.034 0.9729
##
##  States:  1= (s0), 2= Discharged, 3= Death
##
## Likelihood ratio test=43.26  on 6 df, p=1.035e-07
## n= 1595, number of events= 76
```

We see similar, but even more extreme convergence issues as before, we mark the model “Suspicious”.

### 4 Bayesian models with brms

Here we use a set of Bayesian linear models using the `brms` package. There are several classes of model:

- Binary survival models - logistic regression with survival as the outcome.
- Survival time models - Bayesian version of the single-outcome Cox proportional hazards model for the "Death" event.
- Binary discharged models - logistic regression with discharged as the outcome (differs from binary survival in the handling of censored patients)
- Discharged time models - Bayesian version of the single-outcome Cox proportional hazards model for the "Discharged" event
- Categorical models - categorical (multinomial) regression of the "Discharged"/"Hospitalized"/"Death" outcome.

For the final outcome models (binary and categorical) we use three variants of dealing with censoring:

- All: Remove censored patients and keep only those that have data until final outcome
- 7 days: Remove patients with less than 7 days of data and treat the state at day 7 post admission as the outcome
- 28 days: Same as 7 days, but with a longer time frame

The 7 and 28 days models should be somewhat less biased as censored patients are more likely those that spent longer time in hospital and thus have likely more severe disease. We marked the "All" models as "Suspicious" for the purpose of the multiverse analysis.

For each model we show the `brms` formula used, summary of fitted coefficients. See the `brms` manual for detailed explanation of each model. Additionally we show some *posterior predictive checks* (Gabry et al. 2019; Gelman et al. 2020) to verify that models represent the data well. Consult the `bayesplot` package manual for details on the checks used. As another way to check for potential overfitting we use the `loo` package (Vehtari, Gelman, and Gabry 2017) and its Pareto-k diagnostic.

All hypotheses are evaluated via model coefficients of predictors representing treatments, except for sign corrections (e.g. in binary survival models positive effects represent increased survival while for the main analysis we take positive effect as increased mortality, so we flip the sign).

Some derived predictors are used `age_norm` (age, normalized to have roughly mean 0 and standard deviation of 1), `ever_az`, `ever_hcq`, `ever_favipiravir` (whether the patient took the respective medications at any time) and `comorbidities_sum` - the sum of comorbidities as described in Section 1.1.

#### 4.1 Binary survival models

All variants of the model use the same `brms` formula, with family `bernoulli("logit")`.

```
## survival ~ sex + age_norm + smoking + mo(obesity) + ever_hcq + ever_az +
##  ever_favipiravir + (1 | hospital_id)
```

###### 4.1.1 Basic binary model

The fitted coefficients:

```
##  Family: bernoulli
##   Links: mu = logit
## Formula: survival ~ sex + age_norm + smoking + mo(obesity) + ever_hcq + ever_az + ever_favipiravir +
##    Data: data$patient_data (Number of observations: 195)
## Samples: 4 chains, each with iter = 2000; warmup = 1000; thin = 1;
##          total post-warmup samples = 4000
##
## Group-Level Effects:
## ~hospital_id (Number of levels: 10)
##               Estimate Est.Error l-95% CI u-95% CI Rhat Bulk_ESS Tail_ESS
## sd(Intercept)     1.81      0.87     0.68     4.02 1.00     1228     1821
##
## Population-Level Effects:
##                      Estimate Est.Error l-95% CI u-95% CI Rhat Bulk_ESS Tail_ESS
## Intercept                2.65      1.00     0.89     4.79 1.00     1870     2129
## sexM                    -0.70      0.51    -1.72     0.29 1.00     4496     2859
## age_norm                -2.10      0.47    -3.07    -1.23 1.00     3747     2517
## smokingTRUE              1.85      1.26    -0.28     4.66 1.00     4787     2388
## ever_hcqTRUE             1.80      0.72     0.45     3.26 1.00     2940     3006
## ever_azTRUE             -1.04      0.66    -2.41     0.22 1.00     3071     2757
## ever_favipiravirTRUE     4.55      2.95     0.06    11.57 1.00     4170     2666
## moobesity                0.22      0.37    -0.51     0.94 1.00     3343     3038
##
## Simplex Parameters:
##               Estimate Est.Error l-95% CI u-95% CI Rhat Bulk_ESS Tail_ESS
## moobesity1[1]     0.54      0.28     0.04     0.98 1.00     4684     2612
## moobesity1[2]     0.46      0.28     0.02     0.96 1.00     4684     2612
##
## Samples were drawn using sample(hmc). For each parameter, Bulk_ESS
## and Tail_ESS are effective sample size measures, and Rhat is the potential
## scale reduction factor on split chains (at convergence, Rhat = 1).
```

The `error_binned` posterior predictive check - showing the distribution of errors per predicted probability bin.

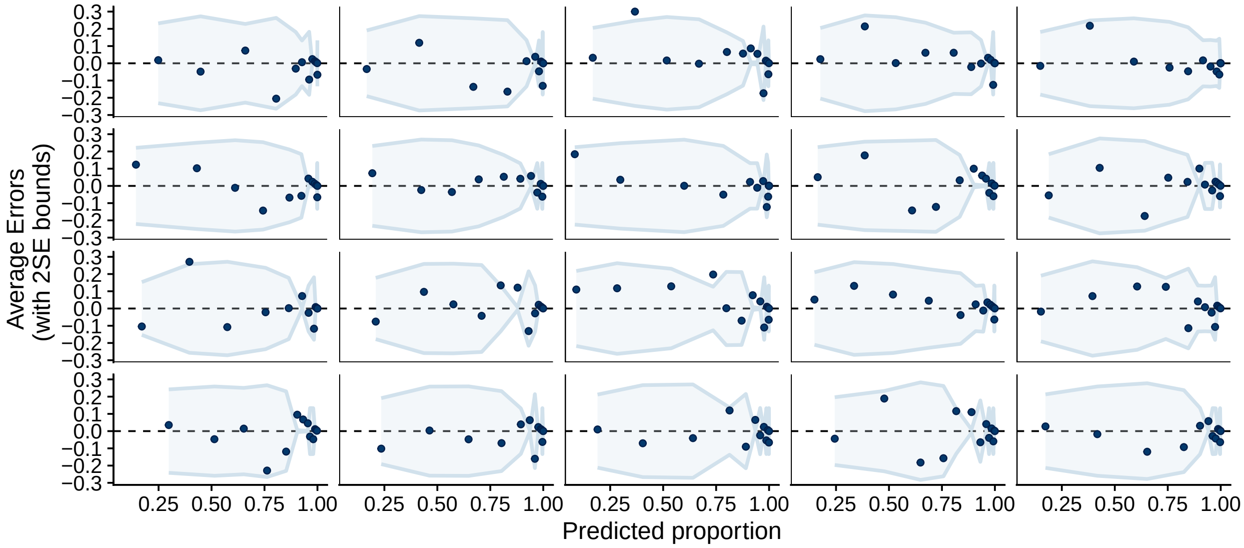

##### 4.1.2 Basic binary model (7 days)

The fitted coefficients:

```
##  Family: bernoulli
##   Links: mu = logit
## Formula: survival ~ sex + age_norm + smoking + mo(obesity) + ever_hcq + ever_az + ever_favipiravir +
##    Data: data$surv_data_7 (Number of observations: 2645)
## Samples: 4 chains, each with iter = 2000; warmup = 1000; thin = 1;
##          total post-warmup samples = 4000
##
## Group-Level Effects:
## ~hospital_id (Number of levels: 10)
##               Estimate Est.Error l-95% CI u-95% CI Rhat Bulk_ESS Tail_ESS
## sd(Intercept)     2.57      1.08     1.21     5.42 1.00     1105     1660
##
## Population-Level Effects:
##                      Estimate Est.Error l-95% CI u-95% CI Rhat Bulk_ESS Tail_ESS
## Intercept                2.74      0.94     0.91     4.77 1.01      900     1395
## sexM                    -0.95      0.14    -1.21    -0.69 1.00     3526     3131
## age_norm                -1.21      0.12    -1.46    -0.99 1.00     2303     2396
## smokingTRUE              2.07      0.29     1.51     2.64 1.00     2619     2501
## ever_hcqTRUE             2.40      0.22     1.97     2.84 1.00     2287     2745
## ever_azTRUE             -1.36      0.17    -1.70    -1.03 1.00     2489     2761
## ever_favipiravirTRUE     7.20      2.43     3.81    13.00 1.00     2775     1567
## moobesity                0.21      0.11    -0.08     0.39 1.00     1547      618
##
## Simplex Parameters:
##               Estimate Est.Error l-95% CI u-95% CI Rhat Bulk_ESS Tail_ESS
## moobesity1[1]     0.84      0.20     0.14     1.00 1.00     1542      577
## moobesity1[2]     0.16      0.20     0.00     0.86 1.00     1542      577
##
## Samples were drawn using sample(hmc). For each parameter, Bulk_ESS
## and Tail_ESS are effective sample size measures, and Rhat is the potential
## scale reduction factor on split chains (at convergence, Rhat = 1).
```

The `error_binned` check:

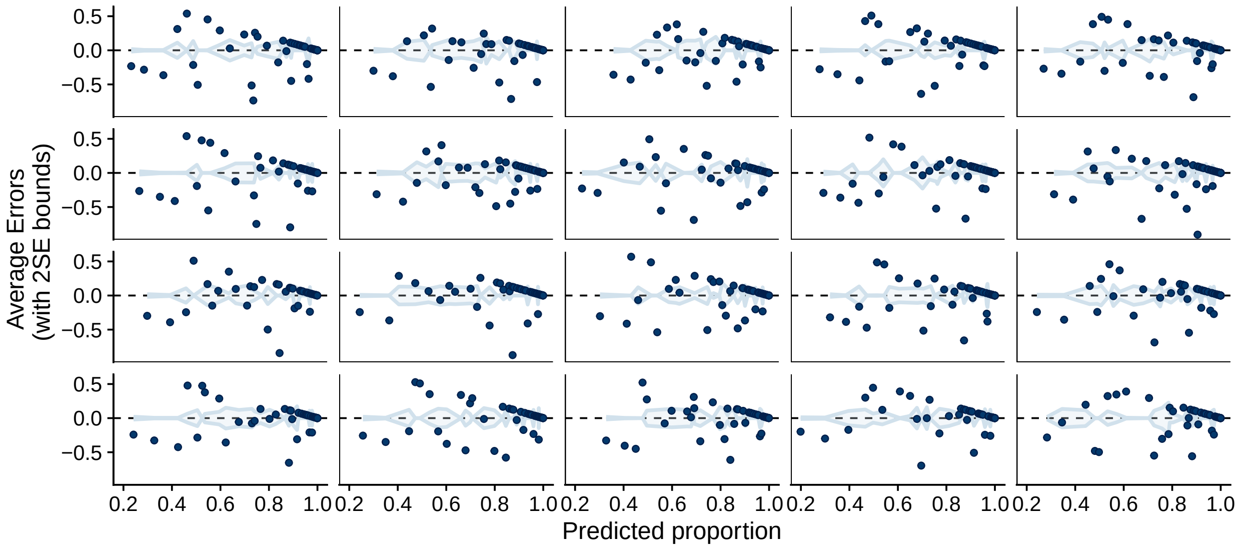

##### 4.1.3 Basic binary model (28 days)

The fitted coefficients:

```
## Warning: There were 1 divergent transitions after warmup. Increasing adapt_delta above may help. See
## transitions-after-warmup

##  Family: bernoulli
##   Links: mu = logit
## Formula: survival ~ sex + age_norm + smoking + mo(obesity) + ever_hcq + ever_az + ever_favipiravir +
##    Data: data$surv_data_28 (Number of observations: 2645)
## Samples: 4 chains, each with iter = 2000; warmup = 1000; thin = 1;
##          total post-warmup samples = 4000
##
## Group-Level Effects:
## ~hospital_id (Number of levels: 10)
##               Estimate Est.Error l-95% CI u-95% CI Rhat Bulk_ESS Tail_ESS
## sd(Intercept)     2.53      1.05     1.20     5.22 1.00     1098     2062
##
## Population-Level Effects:
##                      Estimate Est.Error l-95% CI u-95% CI Rhat Bulk_ESS Tail_ESS
## Intercept                2.85      0.96     0.97     4.93 1.01      883     1204
## sexM                    -0.94      0.14    -1.23    -0.67 1.00     4414     2447
## age_norm                -1.21      0.12    -1.44    -0.99 1.00     3735     2773
## smokingTRUE              2.07      0.30     1.49     2.65 1.00     3742     2965
## ever_hcqTRUE             2.39      0.22     1.97     2.81 1.00     2962     2792
## ever_azTRUE             -1.35      0.17    -1.70    -1.01 1.00     3196     3191
## ever_favipiravirTRUE     7.15      2.33     3.77    12.80 1.00     3619     2303
## moobesity                0.20      0.11    -0.10     0.38 1.00     1962      880
##
## Simplex Parameters:
##               Estimate Est.Error l-95% CI u-95% CI Rhat Bulk_ESS Tail_ESS
## moobesity1[1]     0.84      0.21     0.11     1.00 1.00     1599      797
```

```
## moobesity1[2]     0.16      0.21     0.00     0.89 1.00     1599      797
##
## Samples were drawn using sample(hmc). For each parameter, Bulk_ESS
## and Tail_ESS are effective sample size measures, and Rhat is the potential
## scale reduction factor on split chains (at convergence, Rhat = 1).
```

Note that there is a divergent transition indicating potential computational problems, we will mark the model as "Suspicious".

The `error_binned` check:

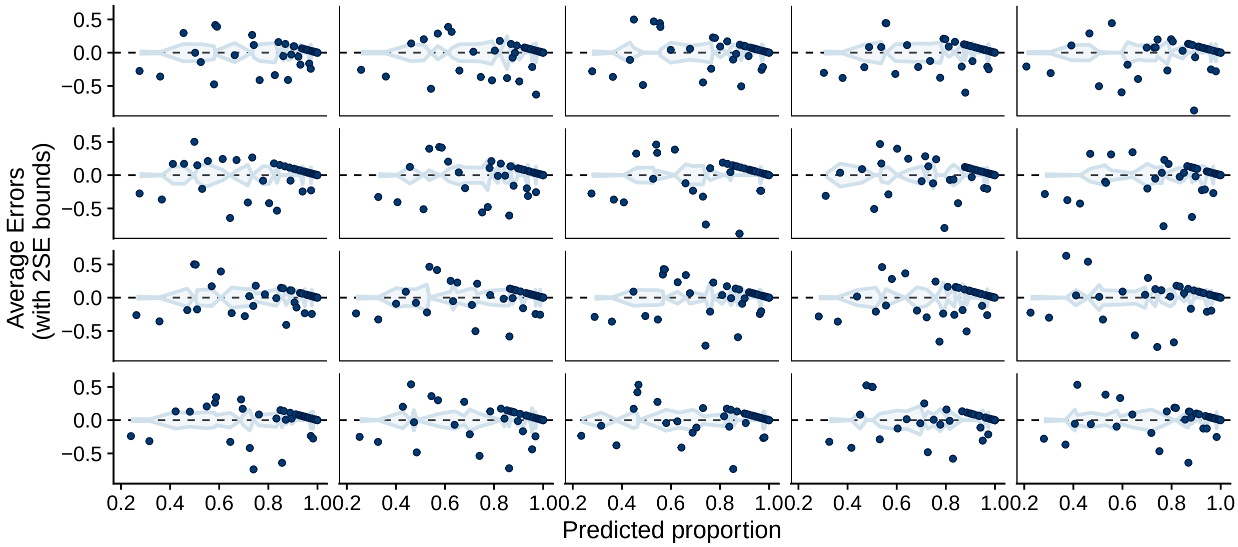

#### 4.2 Survival time models

##### 4.2.1 Fewer adjustments

Those models are described by the `brms` formula with family `brmsfamily("cox", "log")`:

```
## day | cens(censored) ~ sex + age_norm + ever_hcq + ever_az + ever_favipiravir +
##  (1 | hospital_id)
```

Here are the fitted coefficients:

```
##  Family: cox
##   Links: mu = log
## Formula: day | cens(censored) ~ sex + age_norm + ever_hcq + ever_az + ever_favipiravir + (1 | hospita
##    Data: data$surv_data_last_day (Number of observations: 208)
## Samples: 4 chains, each with iter = 2000; warmup = 1000; thin = 1;
##          total post-warmup samples = 4000
##
## Group-Level Effects:
## ~hospital_id (Number of levels: 10)
##               Estimate Est.Error l-95% CI u-95% CI Rhat Bulk_ESS Tail_ESS
## sd(Intercept)     1.17      0.52     0.49     2.49 1.00     1311     2411
##
```

```
## Population-Level Effects:
##                      Estimate Est.Error l-95% CI u-95% CI Rhat Bulk_ESS Tail_ESS
## Intercept               -0.83      0.66    -2.16     0.47 1.00     1910     2279
## sexM                     0.15      0.34    -0.52     0.81 1.00     4572     3116
## age_norm                 1.44      0.32     0.85     2.09 1.00     4303     2881
## ever_hcqTRUE            -1.88      0.51    -2.90    -0.90 1.00     2925     2765
## ever_azTRUE              0.79      0.44    -0.10     1.67 1.00     3299     3029
## ever_favipiravirTRUE    -4.38      2.89   -10.99    -0.10 1.00     3647     2094
##
## Samples were drawn using sample(hmc). For each parameter, Bulk_ESS
## and Tail_ESS are effective sample size measures, and Rhat is the potential
## scale reduction factor on split chains (at convergence, Rhat = 1).
```

###### 4.2.2 Fewer adjustments - First wave only

The same model as above, but using data from the first wave only. Here is the summary of the fit:

```
##  Family: cox
##   Links: mu = log
## Formula: day | cens(censored) ~ sex + age_norm + ever_hcq + ever_az + ever_favipiravir + (1 | hospita
##    Data: data$surv_data_last_day %>% filter(first_wave) (Number of observations: 187)
## Samples: 4 chains, each with iter = 2000; warmup = 1000; thin = 1;
##          total post-warmup samples = 4000
##
## Group-Level Effects:
## ~hospital_id (Number of levels: 10)
##               Estimate Est.Error l-95% CI u-95% CI Rhat Bulk_ESS Tail_ESS
## sd(Intercept)     1.28      0.60     0.52     2.80 1.00     1289     2073
##
## Population-Level Effects:
##                      Estimate Est.Error l-95% CI u-95% CI Rhat Bulk_ESS Tail_ESS
## Intercept               -0.86      0.71    -2.35     0.50 1.00     1939     2356
## sexM                     0.19      0.34    -0.48     0.86 1.00     4462     3169
## age_norm                 1.41      0.34     0.79     2.13 1.00     3679     2808
## ever_hcqTRUE            -1.88      0.51    -2.89    -0.91 1.00     2820     2750
## ever_azTRUE              0.80      0.46    -0.11     1.68 1.00     3451     2908
## ever_favipiravirTRUE    -4.36      2.91   -11.06    -0.20 1.00     3561     2542
##
## Samples were drawn using sample(hmc). For each parameter, Bulk_ESS
## and Tail_ESS are effective sample size measures, and Rhat is the potential
## scale reduction factor on split chains (at convergence, Rhat = 1).
```

```
##
## Computed from 4000 by 187 log-likelihood matrix
##
##          Estimate   SE
## elpd_loo   -187.9 23.0
## p_loo        12.8  2.0
## looic       375.8 46.0
## ------
## Monte Carlo SE of elpd_loo is NA.
##
## Pareto k diagnostic values:
```

```
##                          Count Pct.    Min. n_eff
## (-Inf, 0.5]   (good)     179   95.7%   1325
##  (0.5, 0.7]   (ok)         6    3.2%   663
##    (0.7, 1]   (bad)        1    0.5%   339
##    (1, Inf)   (very bad)   1    0.5%   2000
## See help('pareto-k-diagnostic') for details.
```

###### 4.2.3 More adjustments

For those models the formula changes to:

```
## day | cens(censored) ~ sex + age_norm + smoking + mo(obesity) + ever_hcq +
##  ever_az + ever_favipiravir + (1 | hospital_id)
```

Here are the fitted coefficients - there is a divergent transition so we are possibly fitting an overparemtrized model and we will mark it as "Suspicious":

```
## Warning: There were 1 divergent transitions after warmup. Increasing adapt_delta above may help. See
## transitions-after-warmup
```

```
##  Family: cox
##   Links: mu = log
## Formula: day | cens(censored) ~ sex + age_norm + smoking + mo(obesity) + ever_hcq + ever_az + ever_fa
##    Data: data$surv_data_last_day (Number of observations: 191)
## Samples: 4 chains, each with iter = 2000; warmup = 1000; thin = 1;
##          total post-warmup samples = 4000
##
## Group-Level Effects:
## ~hospital_id (Number of levels: 10)
##               Estimate Est.Error l-95% CI u-95% CI Rhat Bulk_ESS Tail_ESS
## sd(Intercept)     1.43      0.71     0.55     3.45 1.00     1491     2043
##
## Population-Level Effects:
##                      Estimate Est.Error l-95% CI u-95% CI Rhat Bulk_ESS Tail_ESS
## Intercept               -0.35      0.85    -2.16     1.27 1.00     1818     1707
## sexM                     0.23      0.37    -0.50     0.95 1.00     4486     2686
## age_norm                 1.27      0.35     0.62     1.95 1.00     4249     2943
## smokingTRUE             -2.63      1.19    -5.31    -0.64 1.00     3342     2151
## ever_hcqTRUE            -2.25      0.59    -3.44    -1.12 1.00     2423     2829
## ever_azTRUE              1.11      0.53     0.08     2.18 1.00     2900     3035
## ever_favipiravirTRUE    -4.44      2.85   -10.98    -0.28 1.00     3383     2443
## moobesity               -0.19      0.28    -0.74     0.36 1.00     2996     2904
##
## Simplex Parameters:
##               Estimate Est.Error l-95% CI u-95% CI Rhat Bulk_ESS Tail_ESS
## moobesity1[1]     0.55      0.29     0.04     0.98 1.00     3758     2225
## moobesity1[2]     0.45      0.29     0.02     0.96 1.00     3758     2225
##
## Samples were drawn using sample(hmc). For each parameter, Bulk_ESS
## and Tail_ESS are effective sample size measures, and Rhat is the potential
## scale reduction factor on split chains (at convergence, Rhat = 1).
```

In addition, the LOO diagnostics warn us that there were some very influential observations, indicating possible overfitting.

```
##
## Computed from 4000 by 191 log-likelihood matrix
##
##          Estimate   SE
## elpd_loo   -168.5 22.5
## p_loo        14.6  2.4
## looic       337.0 45.0
## ------
## Monte Carlo SE of elpd_loo is NA.
##
## Pareto k diagnostic values:
##                          Count Pct.    Min. n_eff
## (-Inf, 0.5]   (good)     172   90.1%   1198
##  (0.5, 0.7]   (ok)        11    5.8%   663
##    (0.7, 1]   (bad)        3    1.6%   63
##    (1, Inf)   (very bad)   5    2.6%   2000
## See help('pareto-k-diagnostic') for details.
```

###### 4.2.4 More adjustments - First wave only

The same model as above, but fitting only to patients from first wave. Here are the fitted model coefficients:

```
##  Family: cox
##   Links: mu = log
## Formula: day | cens(censored) ~ sex + age_norm + smoking + mo(obesity) + ever_hcq + ever_az + ever_fa
##    Data: data$surv_data_last_day %>% filter(first_wave) (Number of observations: 173)
## Samples: 4 chains, each with iter = 2000; warmup = 1000; thin = 1;
##          total post-warmup samples = 4000
##
## Group-Level Effects:
## ~hospital_id (Number of levels: 10)
##               Estimate Est.Error l-95% CI u-95% CI Rhat Bulk_ESS Tail_ESS
## sd(Intercept)     1.28      0.61     0.48     2.86 1.00     1403     2387
##
## Population-Level Effects:
##                      Estimate Est.Error l-95% CI u-95% CI Rhat Bulk_ESS Tail_ESS
## Intercept               -0.18      0.80    -1.81     1.32 1.00     1895     2194
## sexM                     0.22      0.37    -0.52     0.95 1.00     4364     2932
## age_norm                 1.24      0.35     0.57     1.98 1.00     3933     2708
## smokingTRUE             -2.64      1.19    -5.30    -0.71 1.00     4218     2454
## ever_hcqTRUE            -2.24      0.59    -3.39    -1.14 1.00     2839     2901
## ever_azTRUE              1.08      0.53     0.07     2.13 1.00     3264     2710
## ever_favipiravirTRUE    -4.47      2.85   -11.11    -0.28 1.00     4104     2709
## moobesity               -0.17      0.28    -0.73     0.39 1.00     3431     2562
##
## Simplex Parameters:
##               Estimate Est.Error l-95% CI u-95% CI Rhat Bulk_ESS Tail_ESS
## moobesity1[1]     0.55      0.28     0.03     0.98 1.00     4929     2750
## moobesity1[2]     0.45      0.28     0.02     0.97 1.00     4929     2750
##
```

```
## Samples were drawn using sample(hmc). For each parameter, Bulk_ESS
## and Tail_ESS are effective sample size measures, and Rhat is the potential
## scale reduction factor on split chains (at convergence, Rhat = 1).
```

We do not see divergent transitions, but some overly influential observations (according to `loo`) remain, the model will be marked as "Suspicious".

```
##
## Computed from 4000 by 173 log-likelihood matrix
##
##          Estimate   SE
## elpd_loo   -168.1 22.1
## p_loo        14.3  2.3
## looic       336.2 44.2
## ------
## Monte Carlo SE of elpd_loo is NA.
##
## Pareto k diagnostic values:
##                          Count Pct.    Min. n_eff
## (-Inf, 0.5]   (good)     166   96.0%   801
##  (0.5, 0.7]   (ok)         5    2.9%   2104
##    (0.7, 1]   (bad)        1    0.6%   167
##    (1, Inf)   (very bad)   1    0.6%   2000
## See help('pareto-k-diagnostic') for details.
```

##### 4.3 Binary discharged models

All the models in this section use the `bernoulli("logit")` family (i.e. logistic regression) and follow the `brms` formula

```
## discharged ~ sex + age_norm + smoking + mo(obesity) + ever_hcq + ever_az +
##  ever_favipiravir + (1 | hospital_id)
```

###### 4.3.1 Basic binary model

Here is the summary of the fitted coefficients

```
##  Family: bernoulli
##   Links: mu = logit
## Formula: discharged ~ sex + age_norm + smoking + mo(obesity) + ever_hcq + ever_az + ever_favipiravir
##    Data: data$patient_data (Number of observations: 195)
## Samples: 4 chains, each with iter = 2000; warmup = 1000; thin = 1;
##          total post-warmup samples = 4000
##
## Group-Level Effects:
## ~hospital_id (Number of levels: 10)
##               Estimate Est.Error l-95% CI u-95% CI Rhat Bulk_ESS Tail_ESS
## sd(Intercept)     0.67      0.36     0.09     1.54 1.00     1111     1320
##
## Population-Level Effects:
##                      Estimate Est.Error l-95% CI u-95% CI Rhat Bulk_ESS Tail_ESS
```

```
## Intercept                  0.58      0.45     -0.28     1.52 1.00      2233      1907
## sexM                      -0.55      0.34     -1.23     0.11 1.00      3844      2958
## age_norm                  -1.04      0.24     -1.53    -0.57 1.00      3725      2405
## smokingTRUE                0.06      0.50     -0.93     1.07 1.00      4419      2967
## ever_hcqTRUE               0.91      0.44      0.07     1.78 1.00      2950      2472
## ever_azTRUE                0.30      0.44     -0.57     1.15 1.00      3334      2653
## ever_favipiravirTRUE       1.65      1.31     -0.49     4.70 1.00      3620      2407
## moobesity                 -0.31      0.22     -0.76     0.13 1.00      2901      3015
##
## Simplex Parameters:
##              Estimate Est.Error l-95% CI u-95% CI Rhat Bulk_ESS Tail_ESS
## moobesity1[1]     0.40      0.27     0.02     0.93 1.00     3991     2369
## moobesity1[2]     0.60      0.27     0.07     0.98 1.00     3991     2369
##
## Samples were drawn using sample(hmc). For each parameter, Bulk_ESS
## and Tail_ESS are effective sample size measures, and Rhat is the potential
## scale reduction factor on split chains (at convergence, Rhat = 1).
```

And the error_binned check:

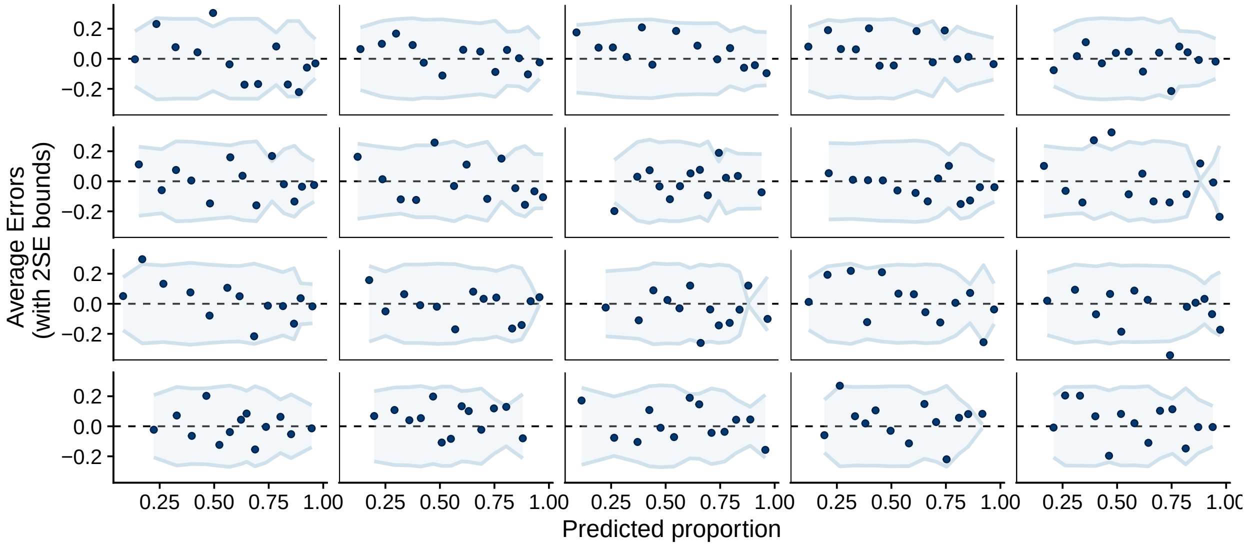

##### 4.3.2 Basic binary model (7 Days)

Here is the summary of the fitted coefficients:

```
##  Family: bernoulli
##   Links: mu = logit
## Formula: discharged ~ sex + age_norm + smoking + mo(obesity) + ever_hcq + ever_az + ever_favipiravir
##    Data: data$surv_data_7 (Number of observations: 2645)
## Samples: 4 chains, each with iter = 2000; warmup = 1000; thin = 1;
##          total post-warmup samples = 4000
##
## Group-Level Effects:
## ~hospital_id (Number of levels: 10)
##              Estimate Est.Error l-95% CI u-95% CI Rhat Bulk_ESS Tail_ESS
## sd(Intercept)     1.70      0.60     0.89     3.20 1.00      884      997
```

```
##
## Population-Level Effects:
##                       Estimate Est.Error l-95% CI u-95% CI Rhat Bulk_ESS Tail_ESS
## Intercept                 0.84      0.57    -0.34     1.98 1.00      818     1275
## sexM                     -0.21      0.10    -0.41    -0.02 1.00     4319     3033
## age_norm                 -0.69      0.07    -0.83    -0.55 1.00     3728     2948
## smokingTRUE              -0.06      0.14    -0.34     0.21 1.00     3796     2863
## ever_hcqTRUE              0.46      0.13     0.20     0.72 1.00     3391     3234
## ever_azTRUE               0.55      0.12     0.30     0.79 1.00     3750     2889
## ever_favipiravirTRUE      2.95      0.62     1.89     4.33 1.00     4144     2188
## moobesity                -0.48      0.06    -0.59    -0.36 1.00     4223     3003
##
## Simplex Parameters:
##                Estimate Est.Error l-95% CI u-95% CI Rhat Bulk_ESS Tail_ESS
## moobesity1[1]      0.05      0.04     0.00     0.16 1.00     2908     1773
## moobesity1[2]      0.95      0.04     0.84     1.00 1.00     2908     1773
##
## Samples were drawn using sample(hmc). For each parameter, Bulk_ESS
## and Tail_ESS are effective sample size measures, and Rhat is the potential
## scale reduction factor on split chains (at convergence, Rhat = 1).
```

And the `error_binned` check.

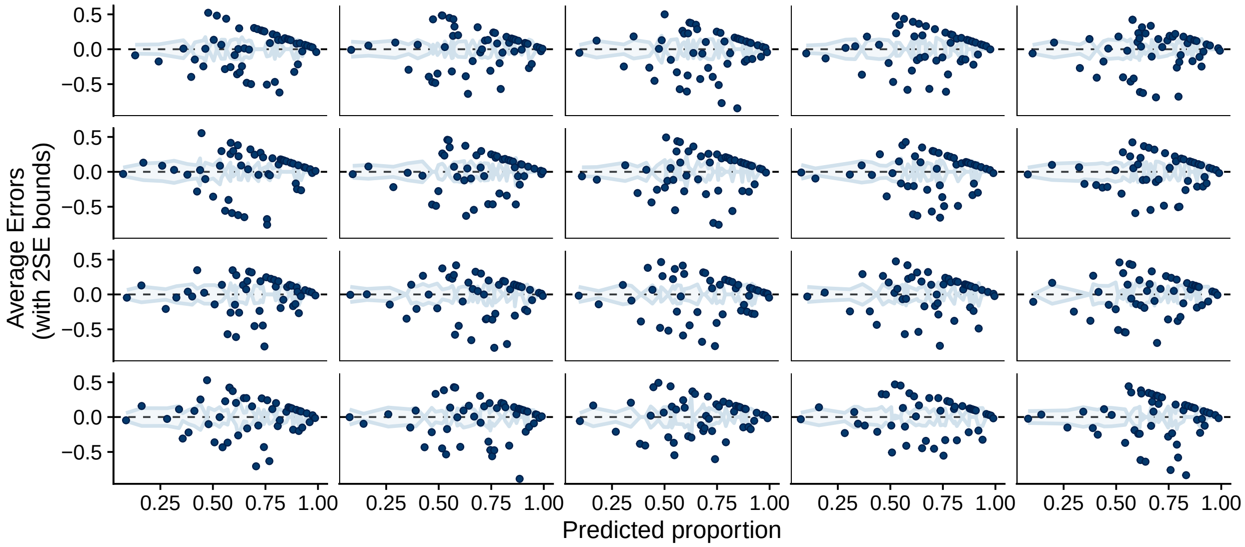

###### 4.3.3 Basic binary model (28 Days)

Here are the fitted coefficients:

```
##  Family: bernoulli
##   Links: mu = logit
## Formula: discharged ~ sex + age_norm + smoking + mo(obesity) + ever_hcq + ever_az + ever_favipiravir
##    Data: data$surv_data_28 (Number of observations: 2645)
## Samples: 4 chains, each with iter = 2000; warmup = 1000; thin = 1;
##          total post-warmup samples = 4000
##
## Group-Level Effects:
```

```
## ~hospital_id (Number of levels: 10)
##               Estimate Est.Error l-95% CI u-95% CI Rhat Bulk_ESS Tail_ESS
## sd(Intercept)     1.68      0.57     0.91     3.06 1.00      989     1816
##
## Population-Level Effects:
##                      Estimate Est.Error l-95% CI u-95% CI Rhat Bulk_ESS Tail_ESS
## Intercept                0.89      0.56    -0.22     2.02 1.00      737     1070
## sexM                    -0.21      0.10    -0.40    -0.02 1.00     4377     2895
## age_norm                -0.69      0.08    -0.84    -0.54 1.00     3436     2570
## smokingTRUE             -0.06      0.14    -0.34     0.21 1.00     4080     3049
## ever_hcqTRUE             0.46      0.13     0.20     0.72 1.00     3125     2885
## ever_azTRUE              0.55      0.12     0.30     0.78 1.00     3214     2876
## ever_favipiravirTRUE     2.94      0.61     1.88     4.25 1.00     4099     2410
## moobesity               -0.48      0.06    -0.60    -0.36 1.00     4392     2861
##
## Simplex Parameters:
##               Estimate Est.Error l-95% CI u-95% CI Rhat Bulk_ESS Tail_ESS
## moobesity1[1]     0.05      0.04     0.00     0.15 1.00     3284     1827
## moobesity1[2]     0.95      0.04     0.85     1.00 1.00     3284     1827
##
## Samples were drawn using sample(hmc). For each parameter, Bulk_ESS
## and Tail_ESS are effective sample size measures, and Rhat is the potential
## scale reduction factor on split chains (at convergence, Rhat = 1).
```

And the `error_binned` check:

```
## Using 10 posterior samples for ppc type 'error_binned' by default.
```

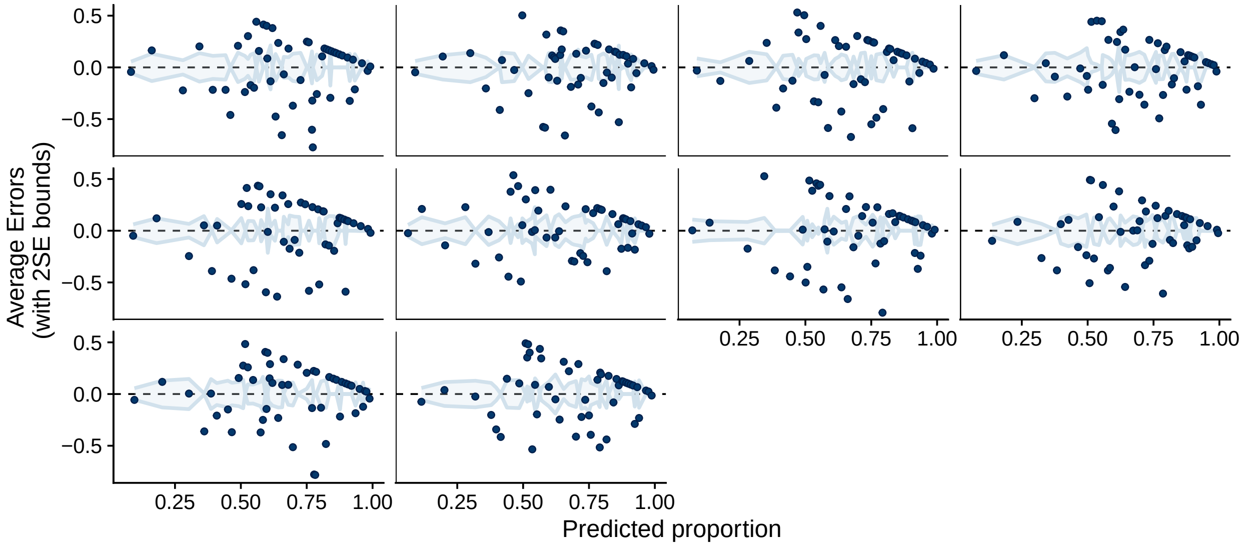

#### 4.4 Discharged time model

The model uses the family `brmsfamily("cox", "log")` and the formula:

```
## day | cens(censored_discharged) ~ sex + age_norm + smoking + mo(obesity) +
##  ever_hcq + ever_az + ever_favipiravir + (1 | hospital_id)
```

The fitted coefficients are:

```
##  Family: cox
##   Links: mu = log
## Formula: day | cens(censored_discharged) ~ sex + age_norm + smoking + mo(obesity) + ever_hcq + ever_a
##    Data: data$surv_data_last_day (Number of observations: 191)
## Samples: 4 chains, each with iter = 2000; warmup = 1000; thin = 1;
##          total post-warmup samples = 4000
##
## Group-Level Effects:
## ~hospital_id (Number of levels: 10)
##               Estimate Est.Error l-95% CI u-95% CI Rhat Bulk_ESS Tail_ESS
## sd(Intercept)     0.41      0.24     0.04     0.95 1.00     1124     1321
##
## Population-Level Effects:
##                      Estimate Est.Error l-95% CI u-95% CI Rhat Bulk_ESS Tail_ESS
## Intercept                2.32      0.37     1.62     3.09 1.00     1995     2537
## sexM                    -0.45      0.20    -0.85    -0.04 1.00     3694     2746
## age_norm                -0.68      0.12    -0.91    -0.45 1.00     4106     3047
## smokingTRUE             -0.46      0.29    -1.06     0.08 1.00     4086     2904
## ever_hcqTRUE            -0.47      0.27    -0.99     0.05 1.00     3315     3061
## ever_azTRUE              0.31      0.25    -0.17     0.79 1.00     3224     3275
## ever_favipiravirTRUE     0.17      0.42    -0.70     0.94 1.00     3488     3090
## moobesity               -0.10      0.13    -0.35     0.14 1.00     3237     2583
##
## Simplex Parameters:
##               Estimate Est.Error l-95% CI u-95% CI Rhat Bulk_ESS Tail_ESS
## moobesity1[1]     0.48      0.28     0.02     0.97 1.00     4112     2185
## moobesity1[2]     0.52      0.28     0.03     0.98 1.00     4112     2185
##
## Samples were drawn using sample(hmc). For each parameter, Bulk_ESS
## and Tail_ESS are effective sample size measures, and Rhat is the potential
## scale reduction factor on split chains (at convergence, Rhat = 1).
```

The `loo` diagnostics warn us that there were some very influential observations.

```
##
## Computed from 4000 by 191 log-likelihood matrix
##
##          Estimate   SE
## elpd_loo   -450.1 23.8
## p_loo        16.4  2.6
## looic       900.1 47.7
## ------
## Monte Carlo SE of elpd_loo is NA.
##
## Pareto k diagnostic values:
##                          Count Pct.    Min. n_eff
## (-Inf, 0.5]   (good)     176   92.1%   1332
##  (0.5, 0.7]   (ok)         5    2.6%   192
##    (0.7, 1]   (bad)        4    2.1%   167
##    (1, Inf)   (very bad)   6    3.1%   2000
## See help('pareto-k-diagnostic') for details.
```

##### 4.5 Categorical models

All the categorical models use the `categorical("logit")` family and the following formula:

```
## outcome3 ~ sex + age_norm + smoking + mo(obesity) + ever_hcq + ever_az +
##  ever_favipiravir + (1 | hospital_id)
```

###### 4.5.1 Basic Categorical model

Fitted coefficients below:

```
##  Family: categorical
##   Links: muDischarged = logit; muDeath = logit
## Formula: outcome3 ~ sex + age_norm + smoking + mo(obesity) + ever_hcq + ever_az + ever_favipiravir +
##    Data: data$patient_data (Number of observations: 195)
## Samples: 4 chains, each with iter = 2000; warmup = 1000; thin = 1;
##          total post-warmup samples = 4000
##
## Group-Level Effects:
## ~hospital_id (Number of levels: 10)
##                              Estimate Est.Error l-95% CI u-95% CI Rhat Bulk_ESS Tail_ESS
## sd(muDischarged_Intercept)     2.40      1.36     0.58     5.90 1.00     1397     1999
## sd(muDeath_Intercept)          3.56      1.68     1.26     7.71 1.00     1618     2415
##
## Population-Level Effects:
##                                  Estimate Est.Error l-95% CI u-95% CI Rhat Bulk_ESS Tail_ESS
## muDischarged_Intercept               5.33      1.89     2.50     9.91 1.00     1716     2075
## muDeath_Intercept                    1.47      2.15    -2.61     6.21 1.00     1927     2247
## muDischarged_sexM                   -2.32      0.99    -4.39    -0.60 1.00     2777     2396
## muDischarged_age_norm               -0.28      0.54    -1.33     0.75 1.00     3989     2963
## muDischarged_smokingTRUE             1.94      1.41    -0.59     5.05 1.00     3313     2752
## muDischarged_ever_hcqTRUE            5.11      2.07     1.49     9.55 1.00     2536     2214
## muDischarged_ever_azTRUE            -0.32      1.42    -2.87     2.61 1.00     3048     2723
## muDischarged_ever_favipiravirTRUE    3.75      3.76    -2.93    11.66 1.00     4116     2845
## muDeath_sexM                        -1.48      1.08    -3.77     0.48 1.00     2772     2621
## muDeath_age_norm                     2.00      0.71     0.62     3.43 1.00     4396     2425
## muDeath_smokingTRUE                 -0.33      1.89    -4.11     3.34 1.00     3777     2940
## muDeath_ever_hcqTRUE                 2.83      2.16    -0.96     7.43 1.00     2527     2123
## muDeath_ever_azTRUE                  0.95      1.53    -1.89     4.09 1.00     3158     2632
## muDeath_ever_favipiravirTRUE        -3.03      3.96   -11.12     4.61 1.00     4720     3129
## muDischarged_moobesity              -0.29      0.48    -1.26     0.65 1.00     2848     3333
## muDeath_moobesity                   -0.30      0.57    -1.46     0.83 1.00     2592     2781
##
## Simplex Parameters:
##                              Estimate Est.Error l-95% CI u-95% CI Rhat Bulk_ESS Tail_ESS
## muDischarged_moobesity1[1]     0.46      0.28     0.02     0.96 1.00     5277     2487
## muDischarged_moobesity1[2]     0.54      0.28     0.04     0.98 1.00     5277     2487
## muDeath_moobesity1[1]          0.54      0.29     0.03     0.98 1.00     5213     2358
## muDeath_moobesity1[2]          0.46      0.29     0.02     0.97 1.00     5213     2369
##
## Samples were drawn using sample(hmc). For each parameter, Bulk_ESS
## and Tail_ESS are effective sample size measures, and Rhat is the potential
## scale reduction factor on split chains (at convergence, Rhat = 1).
```

###### 4.5.2 Basic Categorical model (7 Days)

Summary of model fit:

```
##  Family: categorical
##   Links: muDischarged = logit; muDeath = logit
## Formula: outcome3 ~ sex + age_norm + smoking + mo(obesity) + ever_hcq + ever_az + ever_favipiravir +
##    Data: data$surv_data_7 (Number of observations: 2645)
## Samples: 4 chains, each with iter = 2000; warmup = 1000; thin = 1;
##          total post-warmup samples = 4000
##
## Group-Level Effects:
## ~hospital_id (Number of levels: 10)
##                             Estimate Est.Error l-95% CI u-95% CI Rhat Bulk_ESS Tail_ESS
## sd(muDischarged_Intercept)     1.28      0.39     0.74     2.22 1.00     1257     1885
## sd(muDeath_Intercept)          2.72      1.13     1.29     5.51 1.00     1422     2309
##
## Population-Level Effects:
##                                    Estimate Est.Error l-95% CI u-95% CI Rhat Bulk_ESS Tail_ESS
## muDischarged_Intercept                -1.50      0.47    -2.41    -0.56 1.00     1296     1814
## muDeath_Intercept                     -5.56      1.24    -8.39    -3.42 1.00     1291     2142
## muDischarged_sexM                     -0.49      0.17    -0.83    -0.15 1.00     5194     3044
## muDischarged_age_norm                 -1.44      0.11    -1.67    -1.22 1.00     4624     2723
## muDischarged_smokingTRUE              -0.83      0.23    -1.28    -0.37 1.00     5019     2834
## muDischarged_ever_hcqTRUE             -1.31      0.25    -1.81    -0.82 1.00     3653     2465
## muDischarged_ever_azTRUE               0.12      0.23    -0.34     0.57 1.00     3979     2883
## muDischarged_ever_favipiravirTRUE      0.08      0.33    -0.59     0.72 1.00     5439     3209
## muDeath_sexM                          -0.52      0.29    -1.12     0.05 1.00     4398     2937
## muDeath_age_norm                       2.34      0.33     1.72     3.01 1.00     4470     2910
## muDeath_smokingTRUE                   -8.20      2.35   -13.76    -4.67 1.00     3188     2165
## muDeath_ever_hcqTRUE                  -4.21      0.55    -5.35    -3.17 1.00     3264     2726
## muDeath_ever_azTRUE                    2.53      0.48     1.62     3.51 1.00     3258     2547
## muDeath_ever_favipiravirTRUE          -5.37      2.85   -12.19    -1.30 1.00     3961     2098
## muDischarged_moobesity                 0.03      0.13    -0.23     0.27 1.00     3304     2980
## muDeath_moobesity                      1.01      0.20     0.61     1.40 1.00     4056     2666
##
## Simplex Parameters:
##                             Estimate Est.Error l-95% CI u-95% CI Rhat Bulk_ESS Tail_ESS
## muDischarged_moobesity1[1]     0.55      0.31     0.03     0.99 1.00     3550     2449
## muDischarged_moobesity1[2]     0.45      0.31     0.01     0.97 1.00     3550     2449
## muDeath_moobesity1[1]          0.06      0.06     0.00     0.21 1.00     4668     2100
## muDeath_moobesity1[2]          0.94      0.06     0.79     1.00 1.00     4668     2100
##
## Samples were drawn using sample(hmc). For each parameter, Bulk_ESS
## and Tail_ESS are effective sample size measures, and Rhat is the potential
## scale reduction factor on split chains (at convergence, Rhat = 1).
```

###### 4.5.3 Basic Categorical model (7 Days), first wave only

The same model as above, using only data from the first wave. Summary below:

```
##  Family: categorical
##   Links: muDischarged = logit; muDeath = logit
```

```
## Formula: outcome3 ~ sex + age_norm + smoking + mo(obesity) + ever_hcq + ever_az + ever_favipiravir +
##    Data: data$surv_data_7 %>% filter(first_wave) (Number of observations: 2561)
## Samples: 4 chains, each with iter = 2000; warmup = 1000; thin = 1;
##          total post-warmup samples = 4000
##
## Group-Level Effects:
## ~hospital_id (Number of levels: 10)
##                             Estimate Est.Error l-95% CI u-95% CI Rhat Bulk_ESS Tail_ESS
## sd(muDischarged_Intercept)      1.00      0.33     0.53     1.82 1.00     1124     1961
## sd(muDeath_Intercept)           2.54      1.06     1.18     5.20 1.00     1312     1983
##
## Population-Level Effects:
##                                    Estimate Est.Error l-95% CI u-95% CI Rhat Bulk_ESS Tail_ESS
## muDischarged_Intercept                -1.96      0.41    -2.80    -1.16 1.01     1353     2093
## muDeath_Intercept                     -5.41      1.13    -8.02    -3.48 1.00     1006     1716
## muDischarged_sexM                     -0.33      0.18    -0.69     0.02 1.00     4648     3146
## muDischarged_age_norm                 -1.37      0.11    -1.59    -1.15 1.00     4318     3195
## muDischarged_smokingTRUE              -0.73      0.26    -1.23    -0.22 1.00     4478     2901
## muDischarged_ever_hcqTRUE             -1.21      0.27    -1.74    -0.71 1.00     2338     3008
## muDischarged_ever_azTRUE               0.32      0.24    -0.15     0.79 1.00     3050     3149
## muDischarged_ever_favipiravirTRUE     -0.02      0.34    -0.71     0.65 1.00     4415     2764
## muDeath_sexM                          -0.50      0.30    -1.09     0.10 1.00     4452     3105
## muDeath_age_norm                       2.33      0.33     1.69     2.99 1.00     3395     3018
## muDeath_smokingTRUE                   -8.24      2.39   -14.02    -4.67 1.00     3360     2037
## muDeath_ever_hcqTRUE                  -4.18      0.56    -5.29    -3.12 1.00     2744     2733
## muDeath_ever_azTRUE                    2.52      0.47     1.65     3.48 1.00     3020     2797
## muDeath_ever_favipiravirTRUE          -5.36      2.81   -11.76    -1.29 1.00     3761     2471
## muDischarged_moobesity                 0.12      0.15    -0.20     0.38 1.00     2381     2700
## muDeath_moobesity                      1.00      0.21     0.60     1.40 1.00     3414     3098
##
## Simplex Parameters:
##                             Estimate Est.Error l-95% CI u-95% CI Rhat Bulk_ESS Tail_ESS
## muDischarged_moobesity1[1]      0.65      0.29     0.04     0.99 1.00     2662     2383
## muDischarged_moobesity1[2]      0.35      0.29     0.01     0.96 1.00     2662     2383
## muDeath_moobesity1[1]           0.06      0.05     0.00     0.21 1.00     3855     1783
## muDeath_moobesity1[2]           0.94      0.05     0.79     1.00 1.00     3855     1783
##
## Samples were drawn using sample(hmc). For each parameter, Bulk_ESS
## and Tail_ESS are effective sample size measures, and Rhat is the potential
## scale reduction factor on split chains (at convergence, Rhat = 1).
```

##### 4.5.4 Basic Categorical model (28 Days)

Summary of the model is:

```
##  Family: categorical
##   Links: muDischarged = logit; muDeath = logit
## Formula: outcome3 ~ sex + age_norm + smoking + mo(obesity) + ever_hcq + ever_az + ever_favipiravir +
##    Data: data$surv_data_28 (Number of observations: 2645)
## Samples: 4 chains, each with iter = 2000; warmup = 1000; thin = 1;
##          total post-warmup samples = 4000
##
## Group-Level Effects:
```

```
## ~hospital_id (Number of levels: 10)
##                             Estimate Est.Error l-95% CI u-95% CI Rhat Bulk_ESS Tail_ESS
## sd(muDischarged_Intercept)      4.30      1.54     2.21     8.13 1.00     1361     2208
## sd(muDeath_Intercept)           4.55      1.55     2.31     8.16 1.00     1799     2167
##
## Population-Level Effects:
##                                    Estimate Est.Error l-95% CI u-95% CI Rhat Bulk_ESS Tail_ESS
## muDischarged_Intercept                 4.99      1.77     1.94     9.17 1.00      982     1555
## muDeath_Intercept                     -1.03      1.82    -4.94     2.44 1.00      932     1429
## muDischarged_sexM                     -1.04      0.12    -1.27    -0.82 1.00     4579     3224
## muDischarged_age_norm                 -1.16      0.09    -1.34    -0.98 1.00     3967     3262
## muDischarged_smokingTRUE              -0.50      0.16    -0.82    -0.19 1.00     5106     3110
## muDischarged_ever_hcqTRUE              0.62      0.17     0.29     0.95 1.00     3800     2913
## muDischarged_ever_azTRUE               0.74      0.14     0.47     1.02 1.00     4043     3181
## muDischarged_ever_favipiravirTRUE      1.14      0.22     0.70     1.58 1.00     4644     3273
## muDeath_sexM                          -0.18      0.17    -0.51     0.15 1.00     5021     2847
## muDeath_age_norm                       1.27      0.17     0.92     1.60 1.00     2773     2749
## muDeath_smokingTRUE                   -2.21      0.37    -2.96    -1.51 1.00     5454     2877
## muDeath_ever_hcqTRUE                  -2.54      0.28    -3.09    -2.01 1.00     3448     2907
## muDeath_ever_azTRUE                    2.31      0.20     1.90     2.71 1.00     3476     2781
## muDeath_ever_favipiravirTRUE          -6.97      2.41   -12.69    -3.46 1.00     3605     1893
## muDischarged_moobesity                -0.67      0.09    -0.84    -0.50 1.00     3562     2884
## muDeath_moobesity                      0.24      0.16    -0.13     0.51 1.00     2079     1823
##
## Simplex Parameters:
##                             Estimate Est.Error l-95% CI u-95% CI Rhat Bulk_ESS Tail_ESS
## muDischarged_moobesity1[1]      0.41      0.09     0.23     0.59 1.00     4075     2466
## muDischarged_moobesity1[2]      0.59      0.09     0.41     0.77 1.00     4075     2466
## muDeath_moobesity1[1]           0.19      0.22     0.00     0.88 1.00     2131     1581
## muDeath_moobesity1[2]           0.81      0.22     0.12     1.00 1.00     2131     1581
##
## Samples were drawn using sample(hmc). For each parameter, Bulk_ESS
## and Tail_ESS are effective sample size measures, and Rhat is the potential
## scale reduction factor on split chains (at convergence, Rhat = 1).
```

###### 4.5.5 Basic Categorical model (28 Days), first wave only

The same model, but only for patients from the first wave, summary of the fit:

```
##  Family: categorical
##   Links: muDischarged = logit; muDeath = logit
## Formula: outcome3 ~ sex + age_norm + smoking + mo(obesity) + ever_hcq + ever_az + ever_favipiravir +
##    Data: data$surv_data_28 (Number of observations: 2645)
## Samples: 4 chains, each with iter = 2000; warmup = 1000; thin = 1;
##          total post-warmup samples = 4000
##
## Group-Level Effects:
## ~hospital_id (Number of levels: 10)
##                             Estimate Est.Error l-95% CI u-95% CI Rhat Bulk_ESS Tail_ESS
## sd(muDischarged_Intercept)      4.37      1.58     2.25     8.42 1.00     1207     1989
## sd(muDeath_Intercept)           4.64      1.67     2.33     8.83 1.00     1774     2478
##
## Population-Level Effects:
```

```
##                                    Estimate Est.Error l-95% CI u-95% CI Rhat Bulk_ESS Tail_ESS
## muDischarged_Intercept                 4.91      1.81     1.82     8.91 1.01      878     1239
## muDeath_Intercept                     -1.02      1.90    -5.05     2.76 1.01      983     1478
## muDischarged_sexM                     -1.05      0.12    -1.29    -0.81 1.00     4367     2822
## muDischarged_age_norm                 -1.16      0.09    -1.35    -0.99 1.00     3730     3065
## muDischarged_smokingTRUE              -0.50      0.16    -0.80    -0.19 1.00     4382     3255
## muDischarged_ever_hcqTRUE              0.62      0.16     0.30     0.94 1.00     3547     3263
## muDischarged_ever_azTRUE               0.74      0.14     0.46     1.01 1.00     3556     3182
## muDischarged_ever_favipiravirTRUE      1.15      0.22     0.71     1.58 1.00     4081     2895
## muDeath_sexM                          -0.19      0.17    -0.52     0.14 1.00     4902     3191
## muDeath_age_norm                       1.26      0.18     0.90     1.61 1.00     2872     2886
## muDeath_smokingTRUE                   -2.21      0.36    -2.91    -1.53 1.00     5427     3476
## muDeath_ever_hcqTRUE                  -2.53      0.28    -3.08    -2.01 1.00     3347     2629
## muDeath_ever_azTRUE                    2.30      0.21     1.91     2.73 1.00     3145     2810
## muDeath_ever_favipiravirTRUE          -6.97      2.41   -12.78    -3.51 1.00     3749     2478
## muDischarged_moobesity                -0.67      0.09    -0.84    -0.51 1.00     2576     2714
## muDeath_moobesity                      0.23      0.17    -0.15     0.51 1.00     1959     1898
##
## Simplex Parameters:
##                             Estimate Est.Error l-95% CI u-95% CI Rhat Bulk_ESS Tail_ESS
## muDischarged_moobesity1[1]      0.41      0.09     0.22     0.58 1.00     4167     2197
## muDischarged_moobesity1[2]      0.59      0.09     0.42     0.78 1.00     4167     2197
## muDeath_moobesity1[1]           0.21      0.25     0.00     0.94 1.00     2135     2061
## muDeath_moobesity1[2]           0.79      0.25     0.06     1.00 1.00     2135     2061
##
## Samples were drawn using sample(hmc). For each parameter, Bulk_ESS
## and Tail_ESS are effective sample size measures, and Rhat is the potential
## scale reduction factor on split chains (at convergence, Rhat = 1).
```

##### 4.6 Worst Breathing

Not reported in the main manuscript but we also experimented with ordinal (stopping ratio) models for the worst breathing level experienced by a patient.

```
##  Family: sratio
##   Links: mu = logit; disc = identity
## Formula: worst_breathing ~ sex + age_norm + smoking + mo(obesity) + ever_hcq + ever_az + ever_favipi
##    Data: data$patient_data (Number of observations: 143)
## Samples: 4 chains, each with iter = 2000; warmup = 1000; thin = 1;
##          total post-warmup samples = 4000
##
## Group-Level Effects:
## ~hospital_id (Number of levels: 10)
##               Estimate Est.Error l-95% CI u-95% CI Rhat Bulk_ESS Tail_ESS
## sd(Intercept)     1.16      0.39     0.60     2.08 1.00     1358     2359
##
## Population-Level Effects:
##                      Estimate Est.Error l-95% CI u-95% CI Rhat Bulk_ESS Tail_ESS
## Intercept[1]            -0.38      0.50    -1.35     0.63 1.00     1433     2026
## Intercept[2]             1.40      0.56     0.35     2.53 1.00     1483     2051
## Intercept[3]            -1.49      0.78    -3.03    -0.01 1.00     2359     2354
## Intercept[4]             0.80      0.63    -0.41     2.09 1.00     1755     2132
## Intercept[5]            -1.70      1.13    -4.29     0.24 1.00     2455     2179
```

```
## sexM                      0.20      0.29    -0.37     0.77 1.00     4409     3008
## age_norm                  1.13      0.21     0.73     1.55 1.00     3995     2892
## smokingTRUE              -0.71      0.41    -1.48     0.11 1.00     4117     3295
## ever_hcqTRUE              0.17      0.39    -0.60     0.95 1.00     3307     3171
## ever_azTRUE              -0.02      0.38    -0.74     0.70 1.00     3121     2807
## ever_favipiravirTRUE     -0.86      0.73    -2.31     0.55 1.00     3675     2586
## moobesity                 0.35      0.20    -0.03     0.74 1.00     2516     2701
##
## Simplex Parameters:
##               Estimate Est.Error l-95% CI u-95% CI Rhat Bulk_ESS Tail_ESS
## moobesity1[1]     0.31      0.24     0.01     0.89 1.00     3747     2653
## moobesity1[2]     0.69      0.24     0.11     0.99 1.00     3747     2653
##
## Family Specific Parameters:
##      Estimate Est.Error l-95% CI u-95% CI Rhat Bulk_ESS Tail_ESS
## disc     1.00      0.00     1.00     1.00 1.00     4000     4000
##
## Samples were drawn using sample(hmc). For each parameter, Bulk_ESS
## and Tail_ESS are effective sample size measures, and Rhat is the potential
## scale reduction factor on split chains (at convergence, Rhat = 1).
```

### 5 Hidden Markov Models

#### 5.1 The overall approach for hidden Markov models

To complement the more traditionally used proportional hazards models we also use several variants of hidden Markov models (HMMs). A discrete random process $X_1, X_2, X_3, ...$ satisfies the Markov property whenever for all states $x$ and times $t$ we have $P(X_{t+1} = x|X_1 = x_1, X_2 = x_2, ..., X_t = x_t) = P(X_{t+1} = x|X_t = x_t)$, i.e. when the process is "memoryless". If this process is unobservable, but we can observe another process $Y$ such that $P(Y_t = y|X_1 = x_1, X_2 = x_2, ..., X_n = x_n) = P(Y_t = y|X_t = x_t)$ we call the pair $(X, Y)$ a hidden Markov process.

We treat the breathing support required as a Markov process, either with states directly corresponding to the breathing support and directly observable (Fig. 1a), or with the states carrying a binary improving/worsening component which is unobservable (Fig. 1b) while the breathing support dimension still being completely observable. The former completely observable model is called *simple* in the following text while the latter is called *complex*. This results in a very restricted observation matrix. The reason is that the breathing support actually used is very likely to correspond to the breathing support needed by the patient. In initial versions of our models we also tested models that treated the breathing support observed as a noisy realisation of the actual state, but the fitted probabilities of such imprecise observations were always very low, so we ended up not using them.

Directly modeling the full transition matrix of the Markov process would not make best use of the data as we know there is additional structure expressed in the ordering of states, e.g. that the probability of being discharged from the "AA" state is higher than from the "Oxygen" state, or that transitioning to "Ventilated" is more likely from the "Oxygen" state than from the "AA" state. To incorporate this structure, we follow the approach outlined in (Williams et al. 2020).

We setup a *rate matrix* $R$ so that for any two states $i \neq j$ $R_{i,j}$ is the rate of transition from $i$ to $j$. $R$ is intended to be sparse, i.e. $R_{i,j} \neq 0$ only for transitions explicitly intended to be modeled directly. Additionally the diagonal elements are set as $R_{i,i} = -\sum_{j \neq i} R_{i,j}$.

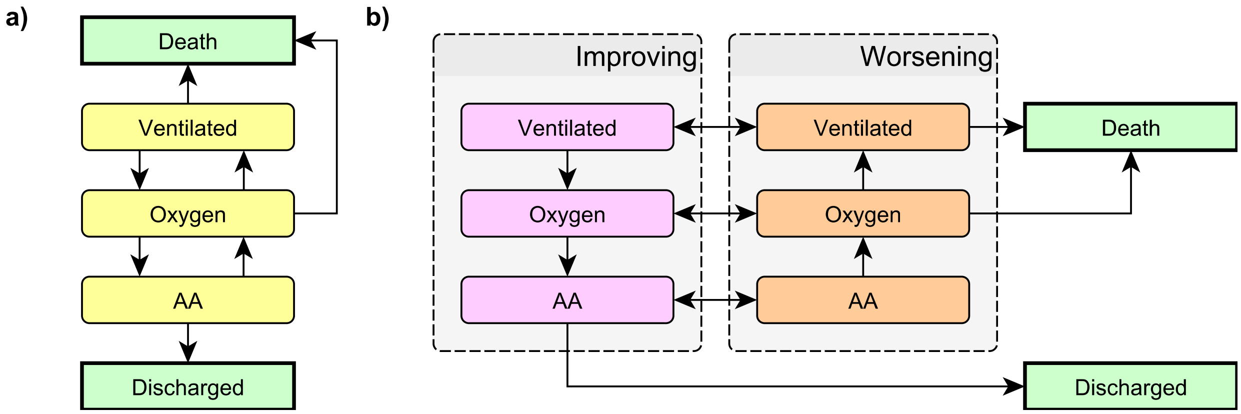

Figure 1: The Markov models used a) simple, b) complex. AA = Ambient air, Oxygen = Nasal oxygen, Ventilated = any form of ventilation, including non-invasive positive-pressure ventilation, mechanical ventilation and extra-corporeal membrane oxygenation. In all models the 'Death' and 'Discharged' states are terminal. In the complex model (c), the 'Improving' and 'Worsening' variants of each non-terminal state are not observable - only the breathing support is observed.

The evolution of the vector $p(t)$ of the state probabilities in continuous time $t$ is then given by the differential equation

Table 1: Rates used in the simple model.

| .from | .to | rate_group | .rate_id |
|---|---|---|---|
| Oxygen | Death | death | Oxygen_Death |
| Ventilated | Death | death | Ventilated_Death |
| AA | Discharged | improve_one | AA_Discharged |
| Oxygen | AA | improve_one | Oxygen_AA |
| Ventilated | Oxygen | improve_one | Ventilated_Oxygen |
| AA | Oxygen | worsen_one | AA_Oxygen |
| Oxygen | Ventilated | worsen_one | Oxygen_Ventilated |

$$\frac{dp(t)}{dt} = Rp$$

Given the initial state probabilities $p(0)$, the solution to this equation is:

$$p(t) = \exp(tR)p(0)$$

where exp is the matrix exponential. We can thus compute a discrete-time transition matrix $S = \exp(R)$ so that $p(t+1) = Sp(t)$. The appeal of this approach is that we can enforce a lot of structure on the transition matrix $S$ while allowing positive probability for almost all transitions. In the case of the complex model, the full transition matrix between the 8 states would have 42 free parameters (no transitions from the "Death" and "Discharged states) while the rate matrix has only 13 free parameters. We could obviously have a sparse transition matrix, but that would make the model overly rigid as transitions that are not modeled directly but actually occur in the data (e.g. from"Oxygen" to "Discharged") would have zero probability.

We also find the rate formulation to be theoretically appealing - the disease progression takes place in continuous time, the discretization into individual days is an artifact of the way we collected data, not the reality.

Finally we put a mixed-effects linear predictor on each of the rates we model so that they are allowed to differ between patients (indexed by $k$) and over (discrete) time (indexed by $t$) $R_{k,t;i,j} = \exp(\mu_{k,t;i,j})$ and $S_{k,t} = \exp(R_{k,t})$. We build the model in the Stan language, using the `brms` package to express the linear predictors $\mu_{k,t;i,j}$ - we use baseline patient characteristics as time-constant predictors and the treatments administered, initiation of best supportive care and markers as time-varying predictors.

As an additional structure, we consider *rate groups* which are: "Improving" (from a breathing level to a better one, including "Discharged"), "Worsening" (from a breathing level to a worse one, not including "Death"), and "Death" (any transition to the "Death" state). In addition, the complex model has "To improving" and "To worsening" which correspond to the switches between the improving and worsening variants of each breathing level.

Below we use three types of models: simple with predictors acting on rate groups (i.e. assuming that any predictor has the same multiplicative effect on all rates in a group), simple with predictors acting on rates directly (i.e. a predictor can have different effect on each rate) and complex with predictors acting on rate groups.

Models will be presented in a unified structure that we explain with the first model and will not be repeated

#### 5.2 Treatments - simple, effects on rate groups

The rates included in the simple model are shown in Table 1.

##### 5.2.1 Only treatments

For this model, the $\log(R_{i,j})$ elements are modeled via the `brms` formula

```
## ~0 + .rate_id + (0 + best_supportive_care + took_hcq + took_az +
##     took_favipiravir + took_convalescent_plasma || rate_group)
```

Our code is setup so that the formula can combine predictors drawn from the description of rates (see Table 1) and those taken from patient's data (both the fixed baseline characteristics and time-varying values). There is a separate coefficient for each rate and then the other predictors act uniformly on each rate group. The `took_XXX` variables are boolean predictors that are set to true for all days since the first dose of the treatment was given - i.e. we assume the treatment alters the overall disease progression irreversibly.

This is the summary of the fitted coefficients. `Q2.5` and `Q97.5` are the boundaries of the central 95% credible interval.

```
##
## Rate coefficients:
##                            Estimate Est.Error      Q2.5     Q97.5
## .rate_idOxygen_Death      -4.215832 0.3322499 -4.887730 -3.605872
## .rate_idVentilated_Death  -3.703096 0.3942088 -4.517820 -2.984310
## .rate_idAA_Discharged     -2.493480 0.1223095 -2.734316 -2.260126
## .rate_idOxygen_AA         -2.379096 0.1371918 -2.651860 -2.110353
## .rate_idVentilated_Oxygen -3.786142 0.3278533 -4.470334 -3.174553
## .rate_idAA_Oxygen         -2.510420 0.1522897 -2.809409 -2.219550
## .rate_idOxygen_Ventilated -4.069646 0.2995599 -4.681351 -3.509412
##
## Per rate group effects:
## , , best_supportive_care
##
##               Estimate Est.Error       Q2.5     Q97.5
## improve_one -0.1894345 0.1823932 -0.5495993 0.1515377
## death        2.5900598 0.3675751  1.8775815 3.3276210
## worsen_one  -0.5631096 0.3426176 -1.2907475 0.0633739
##
## , , took_hcq
##
##               Estimate Est.Error       Q2.5       Q97.5
## improve_one  0.1405928 0.1588004 -0.1637444  0.46431390
## death       -1.1161687 0.4016803 -1.8866447 -0.32596612
## worsen_one  -0.5032025 0.2507977 -1.0291112 -0.02631915
##
## , , took_az
##
##                Estimate Est.Error         Q2.5     Q97.5
## improve_one  0.28835660 0.1675011 -0.007057855 0.6173740
## death       -0.04360711 0.3121402 -0.730328450 0.5576086
## worsen_one   0.24773804 0.2519616 -0.194195200 0.7828337
##
## , , took_favipiravir
##
##               Estimate Est.Error       Q2.5     Q97.5
## improve_one  0.1435700 0.2465105 -0.3051075 0.6663837
## death       -0.3622794 0.9163666 -2.8271655 0.7764941
```

```
## worsen_one   0.1004656 0.4316698 -0.7360294 1.0853618
##
## , , took_convalescent_plasma
##
##                Estimate Est.Error      Q2.5     Q97.5
## improve_one -0.28947980 0.3198814 -1.019491 0.1931317
## death       -0.03488977 0.5293895 -1.245760 1.0644583
## worsen_one   0.03460306 0.4849956 -0.998354 1.0600793
```

Note that the coefficients are hard to interpret directly. E.g. initiated best supportive care (`best_supportive_care`) increases the rate of "Death" but it also might decrease the rate of "worsening" transitions. How should those be combined? To make the results easy to interpret we use the model to build counterfactual predictions for each patient and treatment: what is (according to the model) the probability of the patient being alive at 28 days if they did not receive the treatment at all and what if they received the treatment immediately upon admission. Those two probabilities can then be used to compute log odds-ratio, which is the value we report. Similarly, we compute the log odds-ratio for being still hospitalized against being discharged. In both cases log odds ratio $> 0$ is worse for the patient (higher chance of death, higher chance of being hospitalized) in the treatment group.

Additionally we also look on the (counterfactual) time of hospitalization for patients discharged by day 28 in both groups and - to put it on the same scale, compare the mean of the log of the per-patient ratios of those times, i.e. estimate $> 0$ implies worse outcome (longer hospitalization) in the treatment group. This value is not reported in the main manuscript, only in the supplement.

For this model this results in the following estimates:

```
##                        hypothesis point_estimate  95% CI Low  95% CI High
## 1                       hcq_death    -1.05467091 -1.70906881 -0.401071326
## 2                    hcq_hospital    -0.27337333 -0.92041955  0.377648314
## 3         hcq_hospital (duration)     0.02579360 -0.07439438  0.123956431
## 4                        az_death    -0.12339784 -0.66308910  0.355873667
## 5                     az_hospital    -0.43183519 -1.13600369  0.172827301
## 6          az_hospital (duration)    -0.09635510 -0.21564664  0.002466843
## 7               favipiravir_death    -0.32718564 -2.31011301  0.506285400
## 8            favipiravir_hospital    -0.17069121 -1.43690031  0.872395978
## 9 favipiravir_hospital (duration)    -0.05239507 -0.28103470  0.086746002
```

##### 5.2.2 Treatments + age + sex

In this model the formula is expanded to include age and sex:

```
## ~.rate_id + (0 + best_supportive_care + took_hcq + took_az +
##     took_favipiravir + took_convalescent_plasma || rate_group) +
##     (0 + age_norm + is_male || rate_group) - 1
```

This is the summary of the model parameters

```
##
## Rate coefficients:
##                             Estimate Est.Error      Q2.5     Q97.5
## .rate_idOxygen_Death       -4.595120 0.4523254 -5.527329 -3.777001
## .rate_idVentilated_Death   -3.867152 0.4501387 -4.773306 -3.005340
## .rate_idAA_Discharged      -2.305250 0.1425151 -2.599523 -2.033638
```

```
## .rate_idOxygen_AA         -2.165778 0.1625109 -2.499840 -1.859840
## .rate_idVentilated_Oxygen -3.622863 0.3300599 -4.297243 -3.016692
## .rate_idAA_Oxygen         -2.540922 0.1973151 -2.951096 -2.175123
## .rate_idOxygen_Ventilated -4.103329 0.3200609 -4.747660 -3.493809
##
## Per rate group effects:
## , , best_supportive_care
##
##                Estimate Est.Error       Q2.5       Q97.5
## improve_one -0.01587312 0.1920216 -0.3967748  0.35640425
## death        2.29370212 0.4051330  1.5243670  3.11848750
## worsen_one  -0.63857373 0.3334192 -1.3212617 -0.02787409
##
## , , took_hcq
##
##               Estimate Est.Error       Q2.5       Q97.5
## improve_one  0.1694899 0.1580218 -0.1321744  0.47314237
## death       -0.9697202 0.4021104 -1.7552470 -0.21076892
## worsen_one  -0.5290618 0.2602246 -1.0563615 -0.03696747
##
## , , took_az
##
##                Estimate Est.Error       Q2.5     Q97.5
## improve_one  0.15749238 0.1568806 -0.1040102 0.4882247
## death       -0.01593485 0.2774355 -0.6285269 0.5496306
## worsen_one   0.25394723 0.2646194 -0.1489169 0.8352491
##
## , , took_favipiravir
##
##               Estimate Est.Error       Q2.5     Q97.5
## improve_one  0.1577786 0.2413230 -0.2885593 0.6583563
## death       -0.3433908 0.9055573 -2.4712990 0.7400935
## worsen_one   0.1298194 0.4511428 -0.7627087 1.1521558
##
## , , took_convalescent_plasma
##
##                Estimate Est.Error       Q2.5     Q97.5
## improve_one -0.29614808 0.3110168 -0.9884227 0.1628284
## death       -0.01995540 0.5968901 -1.4823200 1.1294855
## worsen_one   0.06616401 0.5018879 -1.0120010 1.1186048
##
## , , age_norm
##
##               Estimate  Est.Error        Q2.5      Q97.5
## improve_one -0.3765694 0.08166477 -0.53642700 -0.2132916
## death        0.6513867 0.33826402  0.04833222  1.3531750
## worsen_one   0.2188221 0.14326859 -0.04288454  0.5190722
##
## , , is_male
##
##                Estimate Est.Error       Q2.5         Q97.5
## improve_one -0.25862231 0.1383554 -0.5268616 -0.0006126674
## death       -0.06236278 0.2254023 -0.5474087  0.3911553750
## worsen_one  -0.06064242 0.1737792 -0.4327761  0.2747597500
```

And here are the estimates of the log ORs:

```
##                       hypothesis point_estimate  95% CI Low 95% CI High
## 1                      hcq_death   -0.913690966 -1.57954968 -0.31000485
## 2                   hcq_hospital   -0.312318653 -0.88878088  0.27623094
## 3        hcq_hospital (duration)    0.002098133 -0.09826513  0.10359941
## 4                       az_death   -0.007595880 -0.43416602  0.43364195
## 5                    az_hospital   -0.167089136 -0.78242095  0.35891466
## 6         az_hospital (duration)   -0.060907668 -0.18452748  0.02600353
## 7              favipiravir_death   -0.282246765 -1.88231559  0.46253880
## 8           favipiravir_hospital   -0.160823607 -1.21887125  0.80842336
## 9 favipiravir_hospital (duration)   -0.060525867 -0.27788725  0.07779017
```

##### 5.2.3 Treatments + age + sex + hospital

Here the model is further expanded to allow all the rates to vary between hospitals.

```
## ~.rate_id + (0 + best_supportive_care + took_hcq + took_az +
##     took_favipiravir + took_convalescent_plasma || rate_group) +
##     (0 + age_norm + is_male || rate_group) + (0 + .rate_id |
##     hospital_id) - 1
```

Summary of the coefficients:

```
##
## Rate coefficients:
##                           Estimate Est.Error      Q2.5     Q97.5
## .rate_idOxygen_Death      -5.288404 0.7913082 -6.912479 -3.928862
## .rate_idVentilated_Death  -4.436724 0.8968420 -6.407712 -2.841070
## .rate_idAA_Discharged     -2.109148 0.5202503 -3.115755 -1.061986
## .rate_idOxygen_AA         -1.923041 0.3784598 -2.629307 -1.114671
## .rate_idVentilated_Oxygen -3.738980 0.4914368 -4.785708 -2.814447
## .rate_idAA_Oxygen         -2.986250 0.5946552 -4.258511 -1.848681
## .rate_idOxygen_Ventilated -4.619424 1.0023164 -6.849667 -2.780599
##
## Per rate group effects:
## , , best_supportive_care
##
##              Estimate Est.Error       Q2.5     Q97.5
## improve_one -0.1562589 0.3198566 -0.7825889 0.4710578
## death        3.1391377 0.4839133  2.2145795 4.0956130
## worsen_one   0.8828553 0.5015347 -0.1356128 1.8333525
##
## , , took_hcq
##
##              Estimate Est.Error       Q2.5       Q97.5
## improve_one  0.1742087 0.1782372 -0.1689506  0.52529830
## death       -0.7890966 0.4544939 -1.7149362  0.02886456
## worsen_one  -0.6879515 0.3305803 -1.3520760 -0.07892419
##
## , , took_az
##
```

```
##              Estimate Est.Error        Q2.5     Q97.5
## improve_one 0.2095375 0.1740539 -0.07691849 0.5707501
## death       0.1593170 0.3381183 -0.44666638 0.9593603
## worsen_one  0.2792031 0.2999475 -0.18568182 0.9350054
##
## , , took_favipiravir
##
##                Estimate Est.Error       Q2.5     Q97.5
## improve_one  0.38904692 0.3053188 -0.1274124 1.0016167
## death       -0.63395281 1.0493471 -3.3323195 0.6721498
## worsen_one   0.08508405 0.4885467 -0.8898947 1.1128993
##
## , , took_convalescent_plasma
##
##                 Estimate Est.Error       Q2.5     Q97.5
## improve_one -0.153521220 0.2900068 -0.8290607 0.3475114
## death       -0.167341188 0.5462780 -1.5158962 0.8118710
## worsen_one   0.005623158 0.4650567 -1.0571740 0.9949555
##
## , , age_norm
##
##               Estimate  Est.Error       Q2.5      Q97.5
## improve_one -0.3770790 0.08450664 -0.5390817 -0.2095405
## death        0.4205931 0.33469961 -0.1270330  1.1941580
## worsen_one   0.1141534 0.14060286 -0.1446639  0.3953201
##
## , , is_male
##
##                Estimate Est.Error       Q2.5      Q97.5
## improve_one -0.22580499 0.1439910 -0.5065072 0.01893192
## death       -0.01452836 0.2160901 -0.4661775 0.42708385
## worsen_one  -0.08810612 0.1761377 -0.4851204 0.22865298
##
##
## Between-site differences:
##                               Estimate Est.Error       Q2.5    Q97.5
## sd(.rate_idOxygen_Death)      1.2289113 0.5386392 0.48918305 2.595732
## sd(.rate_idVentilated_Death)  1.4293045 0.7061698 0.42266033 3.117474
## sd(.rate_idAA_Discharged)     1.4289804 0.5500054 0.53006352 2.684665
## sd(.rate_idOxygen_AA)         0.8593896 0.3892128 0.34289228 1.818686
## sd(.rate_idVentilated_Oxygen) 0.5342227 0.4933090 0.01365477 1.808610
## sd(.rate_idAA_Oxygen)         1.4534712 0.5209365 0.71404895 2.718592
## sd(.rate_idOxygen_Ventilated) 2.3340190 0.9356141 0.47837583 4.385367
```

We see that the data are consistent with quite high between-site differences (measured in standard deviations of the varying intercepts).

And the resulting log ORs are:

```
##                        hypothesis point_estimate  95% CI Low 95% CI High
## 1                       hcq_death    -0.68228546 -1.30063635  -0.1287519
## 2                    hcq_hospital    -0.36519723 -0.99309072   0.2367156
## 3         hcq_hospital (duration)     0.02570142 -0.09548715   0.1492904
## 4                        az_death     0.06109829 -0.28392407   0.4895574
```

```
## 5                     az_hospital    -0.28824521 -0.95140309   0.2137366
## 6          az_hospital (duration)    -0.08806570 -0.22894466   0.0116890
## 7               favipiravir_death    -0.47604714 -2.22424418   0.2316306
## 8            favipiravir_hospital    -0.52201363 -1.74011388   0.4358066
## 9 favipiravir_hospital (duration)    -0.13318481 -0.42268181   0.0600798
```

##### 5.2.4 Treatments + age + sex + hospital, first wave only

This is the same model as above, but using only the patients from the first wave. Here is the summary of the coefficients.

```
##
## Rate coefficients:
##                             Estimate Est.Error      Q2.5     Q97.5
## .rate_idOxygen_Death      -5.425844 0.7904323 -7.031583 -4.002470
## .rate_idVentilated_Death  -4.420629 0.9182409 -6.337347 -2.808453
## .rate_idAA_Discharged     -2.183918 0.5235758 -3.226913 -1.130864
## .rate_idOxygen_AA         -2.146837 0.3585505 -2.815242 -1.339653
## .rate_idVentilated_Oxygen -3.771194 0.4698175 -4.729465 -2.898836
## .rate_idAA_Oxygen         -3.381452 0.7854534 -5.089521 -1.875075
## .rate_idOxygen_Ventilated -5.041890 0.9185589 -7.110962 -3.474770
##
## Per rate group effects:
## , , best_supportive_care
##
##                Estimate Est.Error       Q2.5     Q97.5
## improve_one -0.04844505 0.3200143 -0.6823124 0.5568627
## death        3.08256630 0.4671809  2.1799643 4.0212493
## worsen_one   1.13622662 0.5258807  0.1111883 2.1186020
##
## , , took_hcq
##
##               Estimate Est.Error       Q2.5       Q97.5
## improve_one  0.1997970 0.1834620 -0.1790296  0.55010910
## death       -0.7978675 0.4617082 -1.7418460  0.04873819
## worsen_one  -0.6749227 0.3419319 -1.3690883 -0.05550335
##
## , , took_az
##
##              Estimate Est.Error        Q2.5     Q97.5
## improve_one 0.2349010 0.1718462 -0.06548213 0.5792773
## death       0.1997852 0.3631851 -0.42524395 1.0295257
## worsen_one  0.3725259 0.3314040 -0.12686475 1.0788885
##
## , , took_favipiravir
##
##                Estimate Est.Error       Q2.5     Q97.5
## improve_one  0.37657942 0.3058163 -0.1324709 0.9880483
## death       -0.68051503 1.1365649 -3.8766932 0.6571443
## worsen_one   0.08840324 0.5022941 -0.9704352 1.1331013
##
## , , took_convalescent_plasma
##
```

```
##              Estimate Est.Error       Q2.5     Q97.5
## improve_one -0.1586224 0.2973284 -0.8587154 0.3431192
## death       -0.1852890 0.5726927 -1.7241615 0.7989332
## worsen_one   0.0145709 0.4599381 -1.0339470 0.9876523
##
## , , age_norm
##
##              Estimate  Est.Error       Q2.5      Q97.5
## improve_one -0.3623931 0.08693749 -0.5288145 -0.1830642
## death        0.4199841 0.32936812 -0.1182811  1.1465528
## worsen_one   0.1058817 0.14290598 -0.1525793  0.3983011
##
## , , is_male
##
##               Estimate Est.Error       Q2.5      Q97.5
## improve_one -0.18483383 0.1413366 -0.4837805 0.03481536
## death        0.01747059 0.2141515 -0.4253779 0.49763193
## worsen_one  -0.04652093 0.1673450 -0.4080518 0.28558253
##
##
## Between-site differences:
##                               Estimate Est.Error       Q2.5    Q97.5
## sd(.rate_idOxygen_Death)      1.3018593 0.5837528 0.50657595 2.733226
## sd(.rate_idVentilated_Death)  1.4835931 0.7022460 0.39415560 3.172421
## sd(.rate_idAA_Discharged)     1.4421834 0.5533401 0.53217343 2.740570
## sd(.rate_idOxygen_AA)         0.6938668 0.3305980 0.27536668 1.523655
## sd(.rate_idVentilated_Oxygen) 0.5476171 0.4965087 0.01444878 1.814049
## sd(.rate_idAA_Oxygen)         1.9287014 0.6649224 0.93792935 3.583924
## sd(.rate_idOxygen_Ventilated) 1.5566317 1.0170608 0.08606677 4.056435
```

And the resulting log ORs/log duration ratios:

```
##                        hypothesis point_estimate  95% CI Low 95% CI High
## 1                       hcq_death    -0.68739322 -1.36442434 -0.13707709
## 2                    hcq_hospital    -0.41032493 -1.02261696  0.22577136
## 3         hcq_hospital (duration)     0.02568462 -0.09249005  0.15122585
## 4                        az_death     0.08910622 -0.25218965  0.54870771
## 5                     az_hospital    -0.30755303 -0.93807537  0.22225830
## 6          az_hospital (duration)    -0.10031081 -0.23430642  0.01071815
## 7               favipiravir_death    -0.49352472 -2.45976948  0.20885468
## 8            favipiravir_hospital    -0.49881475 -1.66622737  0.45434954
## 9 favipiravir_hospital (duration)    -0.12090399 -0.40586571  0.07309198
```

##### 5.2.5 Treatment + age + sex + hospital + comorbidities

For this model, the sum of comorbidities is added as a continuous predictor, resulting in the formula:

```
## ~.rate_id + (0 + best_supportive_care + took_hcq + took_az +
##     took_favipiravir + took_convalescent_plasma || rate_group) +
##     (0 + age_norm + is_male || rate_group) + (0 + .rate_id |
##     hospital_id) + (comorbidities_sum || rate_group) - 1
```

Where `comorbidities_sum` is a comorbidity score as described in Section 1.1.

Below is the summary of the coefficients:

```
## Warning: There were 32 divergent transitions after warmup. Increasing adapt_delta above may help. See
## transitions-after-warmup
```

```
##
## Rate coefficients:
##                             Estimate Est.Error      Q2.5      Q97.5
## .rate_idOxygen_Death      -5.003059  1.506629 -7.905332 -1.6737755
## .rate_idVentilated_Death  -4.159502  1.551154 -7.088794 -0.7272401
## .rate_idAA_Discharged     -1.881390  1.233472 -4.236045  0.8263083
## .rate_idOxygen_AA         -1.721945  1.183734 -4.102720  0.8987637
## .rate_idVentilated_Oxygen -3.493895  1.219940 -6.001599 -0.7495154
## .rate_idAA_Oxygen         -2.790392  1.370815 -5.577954  0.3220440
## .rate_idOxygen_Ventilated -4.464245  1.546941 -7.668501 -1.0592650
##
## Per rate group effects:
## , , best_supportive_care
##
##               Estimate Est.Error       Q2.5     Q97.5
## improve_one -0.1634044 0.3418008 -0.8492947 0.4893899
## death        3.1339189 0.4779736  2.2220842 4.0949833
## worsen_one   0.9180495 0.5121690 -0.1222638 1.8875335
##
## , , took_hcq
##
##               Estimate Est.Error       Q2.5       Q97.5
## improve_one  0.1565026 0.1804471 -0.1956277  0.51813088
## death       -0.7639773 0.4538789 -1.7104110  0.03118275
## worsen_one  -0.7008200 0.3220263 -1.3396968 -0.08912141
##
## , , took_az
##
##              Estimate Est.Error        Q2.5     Q97.5
## improve_one 0.2211568 0.1709527 -0.05920854 0.5677571
## death       0.1727925 0.3458038 -0.45210650 0.9804095
## worsen_one  0.2831740 0.2953952 -0.16921415 0.9265029
##
## , , took_favipiravir
##
##                Estimate Est.Error       Q2.5     Q97.5
## improve_one  0.39449377 0.3195631 -0.1469466 1.0181813
## death       -0.64458069 1.0803184 -3.6167927 0.6093352
## worsen_one   0.07132656 0.4970256 -1.0087658 1.0819448
##
## , , took_convalescent_plasma
##
##                 Estimate Est.Error       Q2.5     Q97.5
## improve_one -0.174130770 0.3077502 -0.9086499 0.3365511
## death       -0.163504711 0.5450151 -1.5143320 0.8195531
## worsen_one  -0.005623113 0.4707515 -1.1086685 0.9696678
##
## , , age_norm
##
```

```
##               Estimate  Est.Error       Q2.5      Q97.5
## improve_one -0.3545234 0.08913313 -0.5262836 -0.1717522
## death        0.4148058 0.32645162 -0.1251076  1.1449213
## worsen_one   0.1088982 0.13965898 -0.1576557  0.3875688
##
## , , is_male
##
##                Estimate Est.Error       Q2.5      Q97.5
## improve_one -0.22372642 0.1413068 -0.5008994 0.01963622
## death       -0.00879197 0.2310201 -0.4970842 0.44378440
## worsen_one  -0.08521231 0.1737829 -0.4605349 0.23393433
##
## , , Intercept
##
##               Estimate Est.Error      Q2.5    Q97.5
## improve_one -0.1311343  1.112744 -2.624913 2.170673
## death       -0.3673608  1.275568 -3.546447 2.011785
## worsen_one  -0.2212276  1.214737 -3.126247 2.271002
##
## , , comorbidities_sum
##
##                 Estimate  Est.Error        Q2.5      Q97.5
## improve_one -0.027907766 0.03681556 -0.11044465 0.03275001
## death        0.025014457 0.06683442 -0.08347936 0.18980060
## worsen_one   0.007974458 0.04482535 -0.08596379 0.10723168
##
##
## Between-site differences:
##                                Estimate Est.Error       Q2.5    Q97.5
## sd(.rate_idOxygen_Death)      1.2453746 0.5414450 0.49606445 2.619669
## sd(.rate_idVentilated_Death)  1.4584735 0.7061733 0.38726348 3.167737
## sd(.rate_idAA_Discharged)     1.4121629 0.5255193 0.54206290 2.616734
## sd(.rate_idOxygen_AA)         0.8609561 0.3740244 0.34618180 1.799829
## sd(.rate_idVentilated_Oxygen) 0.5520420 0.5084830 0.01607347 1.863192
## sd(.rate_idAA_Oxygen)         1.4785950 0.5607568 0.68175853 2.816474
## sd(.rate_idOxygen_Ventilated) 2.3757252 0.8940666 0.76998700 4.358383
```

The sampler warns us of divergent transitions, indicating the posterior might not have been fully explored. This is most likely because we do not have enough data to estimate all the parameters. Here and in the following, we mark models that had such problems as "Suspicious" and do not report their results in the main text of the manuscript.

The resulting log ORs (which cannot be fully trusted):

```
##                        hypothesis point_estimate  95% CI Low 95% CI High
## 1                       hcq_death    -0.66287938 -1.30485402 -0.12126403
## 2                    hcq_hospital    -0.34089246 -0.94648155  0.27561451
## 3         hcq_hospital (duration)     0.03068621 -0.08646427  0.15797308
## 4                        az_death     0.06290151 -0.27781634  0.49720187
## 5                     az_hospital    -0.30692387 -0.95359789  0.19850875
## 6          az_hospital (duration)    -0.09291349 -0.23191008  0.01275098
## 7               favipiravir_death    -0.48815401 -2.27275749  0.25879940
## 8            favipiravir_hospital    -0.52948730 -1.76437954  0.42433240
## 9 favipiravir_hospital (duration)    -0.13611701 -0.45368579  0.05953480
```

###### 5.2.6 Treatment + age + sex + hospital + comorbidities, first wave

Once again, we run the same model but for the first wave data only. Similarly to the previous case this results in divergent transitions and the model should not be fully trusted.

```
## Warning: There were 74 divergent transitions after warmup. Increasing adapt_delta above may help. Se
## transitions-after-warmup
```

```
##
## Rate coefficients:
##                            Estimate Est.Error      Q2.5      Q97.5
## .rate_idOxygen_Death      -5.178315  1.518016 -7.885268 -1.6227890
## .rate_idVentilated_Death  -4.193625  1.654349 -7.268397 -0.3087524
## .rate_idAA_Discharged     -1.989850  1.225696 -4.601736  0.6086232
## .rate_idOxygen_AA         -2.016048  1.162646 -4.479803  0.6212253
## .rate_idVentilated_Oxygen -3.603910  1.194785 -6.092831 -1.0378907
## .rate_idAA_Oxygen         -3.122956  1.426002 -6.082225 -0.1323199
## .rate_idOxygen_Ventilated -4.788397  1.484043 -7.800869 -1.6476167
##
## Per rate group effects:
## , , best_supportive_care
##
##                Estimate Est.Error       Q2.5    Q97.5
## improve_one -0.07117904 0.3285126 -0.7305299 0.555677
## death        3.08531751 0.4782866  2.1943237 4.047337
## worsen_one   1.10456615 0.5265393  0.0242793 2.110785
##
## , , took_hcq
##
##               Estimate Est.Error      Q2.5       Q97.5
## improve_one  0.1799738 0.1810573 -0.171786  0.53702273
## death       -0.7496767 0.4627728 -1.730407  0.06268110
## worsen_one  -0.6759378 0.3373733 -1.379221 -0.06707749
##
## , , took_az
##
##              Estimate Est.Error        Q2.5     Q97.5
## improve_one 0.2309940 0.1726633 -0.07018311 0.5810422
## death       0.1794754 0.3575038 -0.43438938 0.9978248
## worsen_one  0.3627142 0.3226710 -0.12703585 1.0742100
##
## , , took_favipiravir
##
##                Estimate Est.Error       Q2.5     Q97.5
## improve_one  0.38172125 0.3143233 -0.1339441 1.0300175
## death       -0.68446386 1.1108874 -3.4970005 0.5487547
## worsen_one   0.09622206 0.4984177 -0.9445352 1.1119878
##
## , , took_convalescent_plasma
##
##                 Estimate Est.Error       Q2.5     Q97.5
## improve_one -0.174903124 0.3039170 -0.8960845 0.3239413
## death       -0.176633161 0.5564133 -1.6086010 0.7818895
## worsen_one   0.001091935 0.4732028 -1.0203105 1.0092643
```

```
##
## , , age_norm
##
##               Estimate  Est.Error       Q2.5      Q97.5
## improve_one -0.3338099 0.09233415 -0.5127078 -0.1533656
## death        0.4168532 0.34120326 -0.1275545  1.1796428
## worsen_one   0.1230877 0.14365447 -0.1366029  0.4221343
##
## , , is_male
##
##                Estimate Est.Error       Q2.5      Q97.5
## improve_one -0.18262833 0.1404099 -0.4691312 0.04007208
## death        0.01702002 0.2120414 -0.4109178 0.49251423
## worsen_one  -0.05780999 0.1648937 -0.4477814 0.25214860
##
## , , Intercept
##
##                Estimate Est.Error      Q2.5    Q97.5
## improve_one -0.04966823  1.101991 -2.648581 2.411438
## death       -0.36761564  1.373262 -3.890871 2.003843
## worsen_one  -0.18570266  1.248252 -2.991585 2.492426
##
## , , comorbidities_sum
##
##                Estimate  Est.Error       Q2.5      Q97.5
## improve_one -0.03647198 0.04075442 -0.1263832 0.03033277
## death        0.02351214 0.07243926 -0.1161888 0.20069855
## worsen_one  -0.02183021 0.04909566 -0.1298166 0.06886433
##
##
## Between-site differences:
##                               Estimate Est.Error       Q2.5    Q97.5
## sd(.rate_idOxygen_Death)      1.2990110 0.5832834 0.48976205 2.773890
## sd(.rate_idVentilated_Death)  1.4886673 0.7446322 0.45118990 3.337141
## sd(.rate_idAA_Discharged)     1.4528637 0.5199276 0.59790253 2.689811
## sd(.rate_idOxygen_AA)         0.6786886 0.3064324 0.28218585 1.493442
## sd(.rate_idVentilated_Oxygen) 0.5578127 0.4817167 0.02341953 1.768553
## sd(.rate_idAA_Oxygen)         1.8560361 0.6171273 0.93018110 3.331395
## sd(.rate_idOxygen_Ventilated) 1.5270773 0.9424739 0.14638760 3.650600
```

And here are the resulting log ORs

```
##                        hypothesis point_estimate  95% CI Low 95% CI High
## 1                       hcq_death    -0.65267491 -1.35657774 -0.09662939
## 2                    hcq_hospital    -0.38143701 -1.00458093  0.24477629
## 3         hcq_hospital (duration)     0.02682056 -0.08922121  0.14975178
## 4                        az_death     0.07899384 -0.28301213  0.52875360
## 5                     az_hospital    -0.30063742 -0.92779585  0.21678431
## 6          az_hospital (duration)    -0.09755432 -0.23267950  0.01207860
## 7               favipiravir_death    -0.49102424 -2.27197023  0.19584988
## 8            favipiravir_hospital    -0.49845191 -1.72334975  0.42426224
## 9 favipiravir_hospital (duration)    -0.12409341 -0.41918880  0.06917590
```

##### 5.3 Treatments - effect on individual rates

In this set of models the effects (especially the treatments) act directly on the individual rates.

###### 5.3.1 Only treatments

For treatments only, this boils down to the following `brms` formula:

```
## ~0 + .rate_id + (0 + best_supportive_care + took_hcq + took_az +
##     took_favipiravir + took_convalescent_plasma || .rate_id)
```

Resulting in the following fitted coefficients:

```
##
## Rate coefficients:
##                            Estimate Est.Error      Q2.5     Q97.5
## .rate_idOxygen_Death      -4.710917 0.4932821 -5.794824 -3.814654
## .rate_idVentilated_Death  -3.684519 0.4393418 -4.600087 -2.851791
## .rate_idAA_Discharged     -2.653855 0.1625897 -2.981882 -2.344768
## .rate_idOxygen_AA         -2.212690 0.1520514 -2.512143 -1.930578
## .rate_idVentilated_Oxygen -3.742285 0.4394393 -4.674879 -2.941104
## .rate_idAA_Oxygen         -2.556653 0.1688961 -2.886919 -2.233196
## .rate_idOxygen_Ventilated -4.149297 0.3772437 -4.924892 -3.452011
##
## Per rate effects:
## , , best_supportive_care
##
##                     Estimate Est.Error       Q2.5      Q97.5
## Oxygen_Death       3.0707512 0.5174304  2.1330547 4.18768200
## Ventilated_Death   0.5648538 0.8690468 -1.3189652 2.10877700
## AA_Discharged      0.1368959 0.2614552 -0.3781493 0.63077098
## Oxygen_AA         -0.4833282 0.2787818 -1.0786517 0.02666024
## Ventilated_Oxygen -0.1207476 0.9005172 -2.0604895 1.44622675
## AA_Oxygen         -0.5872780 0.3719977 -1.3568375 0.08327057
## Oxygen_Ventilated -0.3689920 0.7161280 -1.8870807 0.88218240
##
## , , took_hcq
##
##                      Estimate Est.Error       Q2.5     Q97.5
## Oxygen_Death      -0.63358431 0.4431023 -1.5821903 0.0570350
## Ventilated_Death  -0.33752877 0.4676477 -1.4111725 0.4297822
## AA_Discharged      0.20530008 0.2134231 -0.1858146 0.6467654
## Oxygen_AA          0.12781838 0.1983559 -0.2539504 0.5432899
## Ventilated_Oxygen  0.07657732 0.4254109 -0.7476083 0.9981709
## AA_Oxygen         -0.29881154 0.2518210 -0.8372465 0.1377619
## Oxygen_Ventilated -0.40784531 0.4519609 -1.4403170 0.2767020
##
## , , took_az
##
##                      Estimate Est.Error         Q2.5     Q97.5
## Oxygen_Death      -0.27411132 0.3653198 -1.104882000 0.3106007
## Ventilated_Death   0.09497623 0.3586235 -0.593571250 0.8733190
## AA_Discharged      0.40961364 0.2382889 -0.005734403 0.8792566
```

```
## Oxygen_AA          0.10771694 0.1928445 -0.260814900 0.5162031
## Ventilated_Oxygen  0.01035815 0.3502848 -0.749380800 0.7280137
## AA_Oxygen          0.10469630 0.2410129 -0.348633975 0.6031122
## Oxygen_Ventilated  0.09527752 0.3606181 -0.579247125 0.8859487
##
## , , took_favipiravir
##
##                     Estimate Est.Error       Q2.5     Q97.5
## Oxygen_Death      -0.20461383 0.7088968 -1.9820917 1.0011298
## Ventilated_Death  -0.27134741 0.6858853 -1.9771035 0.8244588
## AA_Discharged      0.58392640 0.4831536 -0.1566555 1.5706285
## Oxygen_AA         -0.13034856 0.3074461 -0.7968250 0.4505565
## Ventilated_Oxygen -0.01971708 0.5839542 -1.3125457 1.1434143
## AA_Oxygen          0.22850567 0.5323345 -0.7698495 1.4376990
## Oxygen_Ventilated -0.11515105 0.5677672 -1.4727957 0.9626361
##
## , , took_convalescent_plasma
##
##                     Estimate Est.Error       Q2.5     Q97.5
## Oxygen_Death      -0.17547466 0.7430829 -2.0615247 1.0752915
## Ventilated_Death  -0.17171870 0.5779514 -1.5469098 0.8678844
## AA_Discharged      0.17772311 0.4687861 -0.7345606 1.2175978
## Oxygen_AA         -0.49142194 0.4585706 -1.5320643 0.1380505
## Ventilated_Oxygen -0.03858658 0.5099387 -1.2047930 0.9983755
## AA_Oxygen         -0.33854506 0.7447156 -2.3453288 0.7839105
## Oxygen_Ventilated  0.27252742 0.5540765 -0.7023956 1.5572780
```

###### 5.3.2 Treatments + age + sex

```
##
## Rate coefficients:
##                           Estimate Est.Error      Q2.5     Q97.5
## .rate_idOxygen_Death      -4.917443 0.5412247 -6.101910 -3.973653
## .rate_idVentilated_Death  -3.745023 0.5253339 -4.809566 -2.728086
## .rate_idAA_Discharged     -2.560363 0.1841711 -2.916976 -2.203021
## .rate_idOxygen_AA         -2.018383 0.2034479 -2.410569 -1.618755
## .rate_idVentilated_Oxygen -3.690229 0.4944754 -4.735267 -2.766196
## .rate_idAA_Oxygen         -2.597937 0.1975056 -2.994222 -2.215048
## .rate_idOxygen_Ventilated -4.071362 0.4228756 -4.903604 -3.237389
##
## Per rate effects:
## , , best_supportive_care
##
##                     Estimate Est.Error       Q2.5      Q97.5
## Oxygen_Death       2.78964405 0.5370790  1.8443397 3.90200925
## Ventilated_Death   0.38085101 0.8168677 -1.3502572 1.84826150
## AA_Discharged      0.24202708 0.2572615 -0.2709340 0.73292305
## Oxygen_AA         -0.31107987 0.2887041 -0.8981212 0.23243885
## Ventilated_Oxygen -0.02013952 0.9113024 -1.9352320 1.63950100
## AA_Oxygen         -0.67832907 0.3737117 -1.4347390 0.03622249
## Oxygen_Ventilated -0.31419496 0.7081167 -1.8539767 0.97979100
##
## , , took_hcq
```

```
##
##                      Estimate Est.Error       Q2.5      Q97.5
## Oxygen_Death      -0.68673740 0.4112094 -1.5432885 0.01878089
## Ventilated_Death  -0.28901352 0.4934293 -1.3616067 0.57161455
## AA_Discharged      0.38831073 0.2246133 -0.0303905 0.83168368
## Oxygen_AA          0.04951408 0.2097188 -0.3574608 0.46466285
## Ventilated_Oxygen  0.01856717 0.4600850 -0.8950958 0.95012280
## AA_Oxygen         -0.39701833 0.2498635 -0.9017023 0.06427778
## Oxygen_Ventilated -0.46462779 0.4726181 -1.4998740 0.33543838
##
## , , took_az
##
##                      Estimate Est.Error        Q2.5     Q97.5
## Oxygen_Death      -0.15402636 0.2970833 -0.91574477 0.3221084
## Ventilated_Death   0.06604257 0.2903987 -0.49048102 0.7575517
## AA_Discharged      0.21998497 0.2057200 -0.07547659 0.6727024
## Oxygen_AA          0.04685749 0.1686515 -0.28450437 0.4094478
## Ventilated_Oxygen  0.01049010 0.2775804 -0.58297712 0.5913261
## AA_Oxygen          0.13084005 0.2204752 -0.23447370 0.6509747
## Oxygen_Ventilated  0.06789767 0.2840663 -0.48418380 0.7135475
##
## , , took_favipiravir
##
##                      Estimate Est.Error       Q2.5     Q97.5
## Oxygen_Death      -0.18260480 0.7210180 -2.0104012 1.0468495
## Ventilated_Death  -0.31322479 0.7522010 -2.2285393 0.8139802
## AA_Discharged      0.53193110 0.4747803 -0.1954153 1.5327353
## Oxygen_AA         -0.11160293 0.3087067 -0.7802361 0.4778761
## Ventilated_Oxygen  0.04532529 0.5974899 -1.2371173 1.3274505
## AA_Oxygen          0.27901027 0.5626660 -0.6850619 1.5366308
## Oxygen_Ventilated -0.09459512 0.5503762 -1.4142042 0.9404677
##
## , , took_convalescent_plasma
##
##                      Estimate Est.Error       Q2.5     Q97.5
## Oxygen_Death      -0.16399515 0.7769859 -1.9956422 1.2511865
## Ventilated_Death  -0.15309728 0.5846870 -1.4814575 0.8931275
## AA_Discharged      0.26885422 0.5065367 -0.6416918 1.3645363
## Oxygen_AA         -0.52352251 0.4601044 -1.5550993 0.1349168
## Ventilated_Oxygen -0.04206701 0.5151476 -1.1886513 0.9978853
## AA_Oxygen         -0.33918585 0.7617072 -2.3497337 0.7907749
## Oxygen_Ventilated  0.29771191 0.5727628 -0.7193219 1.6368370
##
## , , age_norm
##
##                      Estimate Est.Error        Q2.5       Q97.5
## Oxygen_Death       0.49965699 0.3426666 -0.10503115  1.24735125
## Ventilated_Death   0.64579254 0.5509351 -0.22806115  1.92848625
## AA_Discharged     -0.45982820 0.1027826 -0.65177843 -0.25326142
## Oxygen_AA         -0.22773218 0.1348843 -0.49410992  0.03134707
## Ventilated_Oxygen -0.34121968 0.4371353 -1.31214475  0.42815228
## AA_Oxygen          0.25192866 0.1541410 -0.03741344  0.55786800
## Oxygen_Ventilated -0.03579914 0.3199588 -0.66938602  0.60755678
##
```

```
## , , is_male
##
##                      Estimate Est.Error       Q2.5      Q97.5
## Oxygen_Death      -0.05029578 0.2064924 -0.5128710 0.35090808
## Ventilated_Death  -0.11449379 0.2872267 -0.8443961 0.37453678
## AA_Discharged     -0.18766728 0.1721552 -0.5649861 0.07389984
## Oxygen_AA         -0.18068527 0.1745203 -0.5537462 0.09141911
## Ventilated_Oxygen -0.07175978 0.2707298 -0.7109350 0.42923835
## AA_Oxygen         -0.01467936 0.1649133 -0.3588074 0.32062713
## Oxygen_Ventilated -0.09966910 0.2653013 -0.7626645 0.35003218
```

And those log ORs for the outcomes:

```
##                        hypothesis point_estimate 95% CI Low 95% CI High
## 1                       hcq_death    -0.57779855 -1.1852859 -0.05516884
## 2                    hcq_hospital    -0.54040117 -1.1434829  0.02924813
## 3         hcq_hospital (duration)    -0.07277003 -0.1949307  0.04567395
## 4                        az_death    -0.05725552 -0.4261843  0.23094038
## 5                     az_hospital    -0.21840360 -0.8153749  0.19546909
## 6          az_hospital (duration)    -0.06468453 -0.2060263  0.02138196
## 7               favipiravir_death    -0.12737546 -1.2271531  0.51024298
## 8            favipiravir_hospital    -0.25668831 -1.2821246  0.62815470
## 9 favipiravir_hospital (duration)    -0.17362335 -0.5612307  0.04685571
```

##### 5.3.3 Treatments + age + sex + hospital

We allow the rates to differ by hospital site with this formula:

```
## ~.rate_id + (0 + best_supportive_care + took_hcq + took_az +
##     took_favipiravir + took_convalescent_plasma || .rate_id) +
##     (0 + age_norm + is_male || .rate_id) + (0 + .rate_id | hospital_id) -
##     1
```

Those are the resulting fitted coefficients:

```
##
## Rate coefficients:
##                             Estimate Est.Error      Q2.5     Q97.5
## .rate_idOxygen_Death       -5.389686 0.7435511 -6.986782 -4.033718
## .rate_idVentilated_Death   -3.990028 0.8018825 -5.849321 -2.575978
## .rate_idAA_Discharged      -2.337314 0.5313184 -3.364098 -1.254099
## .rate_idOxygen_AA          -1.819361 0.3634750 -2.517701 -1.104145
## .rate_idVentilated_Oxygen -3.715204 0.6290809 -4.993261 -2.488173
## .rate_idAA_Oxygen          -3.149736 0.6562190 -4.590977 -1.906234
## .rate_idOxygen_Ventilated -4.313558 1.0467673 -6.636208 -2.392459
##
## Per rate effects:
## , , best_supportive_care
##
##                     Estimate Est.Error       Q2.5     Q97.5
## Oxygen_Death      3.56693175 0.5694473  2.5278950 4.7940485
## Ventilated_Death  0.87581418 1.1688660 -1.4804277 3.1016113
```

```
## AA_Discharged      0.18485242 0.5925790 -0.9805677 1.2514023
## Oxygen_AA         -0.37427380 0.4144038 -1.2277067 0.4175796
## Ventilated_Oxygen -0.02903078 1.1874371 -2.5372538 2.2211420
## AA_Oxygen          1.34639158 0.5297476  0.2723395 2.3710238
## Oxygen_Ventilated -0.90005315 0.9997158 -3.0093903 1.0162060
##
## , , took_hcq
##
##                     Estimate Est.Error       Q2.5      Q97.5
## Oxygen_Death      -0.60108826 0.4582221 -1.5842800 0.17004233
## Ventilated_Death  -0.40550206 0.5453086 -1.6342115 0.50179533
## AA_Discharged      0.39182850 0.2602462 -0.1002337 0.91151008
## Oxygen_AA          0.08001687 0.2305538 -0.3770460 0.52701612
## Ventilated_Oxygen  0.02270885 0.4722070 -0.8905145 1.04300300
## AA_Oxygen         -0.50520327 0.3112867 -1.1393082 0.02916149
## Oxygen_Ventilated -0.44754674 0.4853255 -1.5679612 0.34352890
##
## , , took_az
##
##                     Estimate Est.Error        Q2.5     Q97.5
## Oxygen_Death      0.006160372 0.2781069 -0.58407775 0.5921735
## Ventilated_Death  0.065374596 0.3093939 -0.51693055 0.7880368
## AA_Discharged     0.263593843 0.2254026 -0.07280137 0.7619368
## Oxygen_AA         0.037277579 0.1864125 -0.32841215 0.4621667
## Ventilated_Oxygen 0.017530102 0.2918092 -0.57718570 0.6408244
## AA_Oxygen         0.165622781 0.2419668 -0.23558902 0.7233703
## Oxygen_Ventilated 0.036307074 0.3054630 -0.58925098 0.7268362
##
## , , took_favipiravir
##
##                     Estimate Est.Error       Q2.5     Q97.5
## Oxygen_Death      -0.16439532 0.7094092 -1.8763385 1.0454045
## Ventilated_Death  -0.34520662 0.7124642 -2.2108748 0.6329145
## AA_Discharged      0.47672075 0.4562060 -0.2200845 1.4638853
## Oxygen_AA          0.11022745 0.3262145 -0.5352054 0.8120634
## Ventilated_Oxygen  0.01596325 0.5717970 -1.1918095 1.2508430
## AA_Oxygen          0.17780896 0.5026829 -0.7554800 1.3135455
## Oxygen_Ventilated -0.16626114 0.5624969 -1.5518885 0.7934875
##
## , , took_convalescent_plasma
##
##                      Estimate Est.Error       Q2.5     Q97.5
## Oxygen_Death      -0.107613978 0.6256989 -1.5248740 1.0105213
## Ventilated_Death  -0.159417219 0.5127778 -1.4683825 0.6818925
## AA_Discharged      0.169029329 0.4114131 -0.5669959 1.1254470
## Oxygen_AA         -0.243983408 0.3720606 -1.1500297 0.3090424
## Ventilated_Oxygen  0.001099001 0.4525372 -0.9655146 0.9517381
## AA_Oxygen         -0.260887621 0.6542224 -2.0831642 0.6498421
## Oxygen_Ventilated  0.149556839 0.4934053 -0.7510609 1.3298443
##
## , , age_norm
##
##                     Estimate Est.Error       Q2.5        Q97.5
## Oxygen_Death       0.17630798 0.2835660 -0.3201222  0.802644250
```

```
## Ventilated_Death   0.52799684 0.5395435 -0.2421271  1.871090250
## AA_Discharged     -0.43640967 0.1111243 -0.6582790 -0.224338375
## Oxygen_AA         -0.25779579 0.1349132 -0.5277786 -0.003185585
## Ventilated_Oxygen -0.25270937 0.4193652 -1.2222748  0.498308775
## AA_Oxygen          0.13388618 0.1411668 -0.1383609  0.418708900
## Oxygen_Ventilated -0.03951724 0.3270301 -0.6943617  0.623940450
##
## , , is_male
##
##                      Estimate Est.Error       Q2.5      Q97.5
## Oxygen_Death       0.004960797 0.2053053 -0.4314690 0.42555020
## Ventilated_Death  -0.103483369 0.2851092 -0.8175518 0.38158435
## AA_Discharged     -0.174629033 0.1699750 -0.5582279 0.07913845
## Oxygen_AA         -0.156269446 0.1692538 -0.5353732 0.10029863
## Ventilated_Oxygen -0.050973528 0.2602506 -0.6579274 0.44256355
## AA_Oxygen         -0.009746496 0.1561747 -0.3471183 0.30244165
## Oxygen_Ventilated -0.130756200 0.2771215 -0.8197268 0.31741540
##
##
## Between-site differences:
##                               Estimate Est.Error       Q2.5    Q97.5
## sd(.rate_idOxygen_Death)      1.2304310 0.5273803 0.49239158 2.478523
## sd(.rate_idVentilated_Death)  0.8815374 0.6703753 0.04321626 2.546297
## sd(.rate_idAA_Discharged)     1.4737399 0.5377209 0.58544493 2.702120
## sd(.rate_idOxygen_AA)         0.7300807 0.3285253 0.27589603 1.555482
## sd(.rate_idVentilated_Oxygen) 0.6448478 0.5535480 0.01717033 2.079109
## sd(.rate_idAA_Oxygen)         1.5894125 0.5646597 0.76599605 2.973641
## sd(.rate_idOxygen_Ventilated) 2.4328888 0.9262774 0.82711545 4.525248
```

And the resulting estimates:

```
##                       hypothesis point_estimate 95% CI Low 95% CI High
## 1                      hcq_death    -0.48757458 -1.0681906 -0.02738464
## 2                   hcq_hospital    -0.52976674 -1.1446511  0.05010341
## 3        hcq_hospital (duration)    -0.04194849 -0.1767536  0.09344148
## 4                       az_death    -0.00316350 -0.2595869  0.26865261
## 5                    az_hospital    -0.24914713 -0.8316412  0.17447269
## 6         az_hospital (duration)    -0.07687852 -0.2313394  0.01769447
## 7              favipiravir_death    -0.19423778 -1.0682390  0.28696623
## 8           favipiravir_hospital    -0.39152943 -1.4738497  0.40329512
## 9 favipiravir_hospital (duration)    -0.14904743 -0.5124777  0.05678035
```

##### 5.3.4 Treatments + age + sex + hospital, first_Wave

This is same as the previous model, but limiting only to first wave patients, the fitted coefficients:

```
##
## Rate coefficients:
##                           Estimate Est.Error      Q2.5     Q97.5
## .rate_idOxygen_Death      -5.558064 0.7688775 -7.193924 -4.104326
## .rate_idVentilated_Death  -4.020669 0.8497940 -5.891527 -2.542316
## .rate_idAA_Discharged     -2.459103 0.5518109 -3.557794 -1.349107
## .rate_idOxygen_AA         -2.013020 0.3675082 -2.692042 -1.250283
```

```
## .rate_idVentilated_Oxygen -3.707845 0.6983528 -5.154222 -2.358363
## .rate_idAA_Oxygen         -3.596343 0.7914659 -5.354980 -2.136126
## .rate_idOxygen_Ventilated -4.658559 0.9495634 -6.874206 -2.986199
##
## Per rate effects:
## , , best_supportive_care
##
##                      Estimate Est.Error       Q2.5     Q97.5
## Oxygen_Death       3.51548467 0.6006276  2.4351505 4.8532748
## Ventilated_Death   0.89057219 1.1485363 -1.3637400 3.1508568
## AA_Discharged      0.50606012 0.6262913 -0.7400189 1.6766858
## Oxygen_AA         -0.28577081 0.4117326 -1.1180788 0.4817166
## Ventilated_Oxygen -0.06762629 1.2293326 -2.6900060 2.2514570
## AA_Oxygen          1.72842200 0.5674330  0.5821529 2.7774278
## Oxygen_Ventilated -0.70531554 1.0090954 -2.8372152 1.0349543
##
## , , took_hcq
##
##                      Estimate Est.Error        Q2.5      Q97.5
## Oxygen_Death      -0.55290253 0.4464280 -1.52283400 0.14184355
## Ventilated_Death  -0.39868927 0.5407851 -1.63557675 0.48150113
## AA_Discharged      0.40835653 0.2703726 -0.06546104 0.95343980
## Oxygen_AA          0.12905357 0.2318877 -0.32337610 0.60095490
## Ventilated_Oxygen  0.01553081 0.4519471 -0.91060640 0.95299685
## AA_Oxygen         -0.44698361 0.3263774 -1.16211775 0.09700043
## Oxygen_Ventilated -0.38280874 0.4885557 -1.46948375 0.44499675
##
## , , took_az
##
##                      Estimate Est.Error        Q2.5     Q97.5
## Oxygen_Death      0.001629615 0.3029407 -0.66848427 0.6623135
## Ventilated_Death  0.070325518 0.3200217 -0.53832075 0.7976556
## AA_Discharged     0.280508412 0.2348394 -0.06910628 0.7808002
## Oxygen_AA         0.056943694 0.1912176 -0.30497210 0.4592269
## Ventilated_Oxygen 0.031250935 0.3264856 -0.63531987 0.7303115
## AA_Oxygen         0.201809137 0.2663024 -0.20244220 0.8195795
## Oxygen_Ventilated 0.080396542 0.3196373 -0.49509400 0.8264097
##
## , , took_favipiravir
##
##                      Estimate Est.Error       Q2.5     Q97.5
## Oxygen_Death      -0.15908668 0.6234287 -1.6935123 0.9047831
## Ventilated_Death  -0.31067605 0.6841309 -2.0816612 0.6763376
## AA_Discharged      0.44963889 0.4455647 -0.2128740 1.4227933
## Oxygen_AA          0.08986942 0.3106824 -0.5298750 0.7514286
## Ventilated_Oxygen  0.02075968 0.5659742 -1.2697360 1.1880500
## AA_Oxygen          0.14961027 0.4914800 -0.7835338 1.2648940
## Oxygen_Ventilated -0.15499672 0.5549720 -1.5267202 0.8381233
##
## , , took_convalescent_plasma
##
##                      Estimate Est.Error       Q2.5     Q97.5
## Oxygen_Death      -0.13285886 0.6390748 -1.6798877 0.9660236
## Ventilated_Death  -0.17497627 0.5505695 -1.5902085 0.7854215
```

```
## AA_Discharged      0.18503685 0.4386137 -0.6075900 1.2510840
## Oxygen_AA         -0.27985931 0.3818354 -1.1574202 0.3144782
## Ventilated_Oxygen -0.01055283 0.4550537 -1.0273735 0.9150556
## AA_Oxygen         -0.27275281 0.6436307 -1.9435930 0.6459034
## Oxygen_Ventilated  0.17401297 0.4830431 -0.7156234 1.3089240
##
## , , age_norm
##
##                        Estimate Est.Error       Q2.5       Q97.5
## Oxygen_Death       0.1717435912 0.2843401 -0.3166451  0.81479848
## Ventilated_Death   0.5219027502 0.5331660 -0.2631424  1.80113975
## AA_Discharged     -0.4228588909 0.1150788 -0.6410277 -0.19209092
## Oxygen_AA         -0.2293489620 0.1388029 -0.4984668  0.04681031
## Ventilated_Oxygen -0.2411437448 0.4100415 -1.1500745  0.46583075
## AA_Oxygen          0.1075468869 0.1427321 -0.1557263  0.39299068
## Oxygen_Ventilated -0.0005218652 0.3190502 -0.5986968  0.63793358
##
## , , is_male
##
##                       Estimate Est.Error       Q2.5     Q97.5
## Oxygen_Death       0.024164534 0.1877965 -0.3697024 0.4471552
## Ventilated_Death  -0.083039799 0.2659572 -0.7691731 0.3705726
## AA_Discharged     -0.127946866 0.1585664 -0.4939566 0.1043084
## Oxygen_AA         -0.118127958 0.1598894 -0.4857158 0.1410480
## Ventilated_Oxygen -0.050430272 0.2337566 -0.6103729 0.3804343
## AA_Oxygen          0.007351256 0.1512244 -0.3175645 0.3227494
## Oxygen_Ventilated -0.093096302 0.2566234 -0.7357207 0.3376813
##
##
## Between-site differences:
##                               Estimate Est.Error       Q2.5    Q97.5
## sd(.rate_idOxygen_Death)      1.2581178 0.5315577 0.49681468 2.558173
## sd(.rate_idVentilated_Death)  0.9081897 0.7113655 0.03189651 2.733293
## sd(.rate_idAA_Discharged)     1.5156049 0.5463138 0.63043848 2.748029
## sd(.rate_idOxygen_AA)         0.6296656 0.3242888 0.21381750 1.459377
## sd(.rate_idVentilated_Oxygen) 0.6956001 0.5923318 0.01911589 2.244946
## sd(.rate_idAA_Oxygen)         2.0341926 0.6602188 1.04675625 3.560530
## sd(.rate_idOxygen_Ventilated) 1.7388723 1.0153005 0.15112568 3.989833
```

And the log(OR) estimates:

```
##                        hypothesis point_estimate 95% CI Low  95% CI High
## 1                       hcq_death   -0.460579609 -1.0418478 -0.005708268
## 2                    hcq_hospital   -0.557987844 -1.2195722  0.042063608
## 3         hcq_hospital (duration)   -0.044401685 -0.1780163  0.086813326
## 4                        az_death   -0.004596722 -0.2788405  0.294683231
## 5                     az_hospital   -0.258263394 -0.8743869  0.175085558
## 6          az_hospital (duration)   -0.078328790 -0.2251293  0.019829036
## 7               favipiravir_death   -0.179032872 -0.9503765  0.232911690
## 8            favipiravir_hospital   -0.358111059 -1.3836482  0.400022541
## 9 favipiravir_hospital (duration)   -0.131201891 -0.4832106  0.063229979
```

##### 5.3.5 Treatment + age + sex + hospital + comorbidities

Adding comorbidities, the complete formula:

```
## ~.rate_id + (0 + best_supportive_care + took_hcq + took_az +
##     took_favipiravir + took_convalescent_plasma || .rate_id) +
##     (0 + age_norm + is_male || .rate_id) + (0 + .rate_id | hospital_id) +
##     (comorbidities_sum || .rate_id) - 1
```

Where `comorbidities_sum` is a comorbidity score as described in Section 1.1.

Coefficient estimates - note the divergent transitions, indicating the results are not completely trustworthy:

```
## Warning: There were 101 divergent transitions after warmup. Increasing adapt_delta above may help. Se
## transitions-after-warmup
```

```
##
## Rate coefficients:
##                             Estimate Est.Error      Q2.5      Q97.5
## .rate_idOxygen_Death      -5.244947  1.475958 -8.164087 -2.0257655
## .rate_idVentilated_Death  -4.011087  1.551564 -7.178683 -0.6246043
## .rate_idAA_Discharged     -2.177257  1.358630 -4.954089  0.6802268
## .rate_idOxygen_AA         -1.809291  1.214719 -4.476222  0.8336526
## .rate_idVentilated_Oxygen -3.601467  1.421527 -6.542673 -0.4122783
## .rate_idAA_Oxygen         -3.008075  1.343670 -5.631302  0.1132083
## .rate_idOxygen_Ventilated -4.439611  1.685857 -7.735733 -1.2845812
##
## Per rate effects:
## , , best_supportive_care
##
##                      Estimate Est.Error       Q2.5     Q97.5
## Oxygen_Death       3.57487001 0.6104319  2.4488977 4.9045225
## Ventilated_Death   0.87397007 1.1167422 -1.3824767 3.0338312
## AA_Discharged      0.14151331 0.5876885 -1.1323117 1.2119560
## Oxygen_AA         -0.37105760 0.3977317 -1.1511742 0.3869154
## Ventilated_Oxygen -0.05625635 1.2223624 -2.6190330 2.2627890
## AA_Oxygen          1.33511245 0.5434398  0.1906756 2.3702538
## Oxygen_Ventilated -0.91209621 0.9966997 -3.0742510 0.8584295
##
## , , took_hcq
##
##                      Estimate Est.Error       Q2.5      Q97.5
## Oxygen_Death      -0.55437832 0.4457477 -1.5167905 0.15403343
## Ventilated_Death  -0.40048566 0.5629833 -1.6630385 0.56122705
## AA_Discharged      0.38018674 0.2607824 -0.1070806 0.87564288
## Oxygen_AA          0.07311985 0.2332779 -0.3785148 0.53668645
## Ventilated_Oxygen  0.01510268 0.4881665 -0.9839724 1.05066800
## AA_Oxygen         -0.49941605 0.3139473 -1.1458755 0.02649509
## Oxygen_Ventilated -0.41859898 0.4930247 -1.5192670 0.39618820
##
## , , took_az
##
##                      Estimate Est.Error       Q2.5     Q97.5
## Oxygen_Death      0.006663826 0.2927697 -0.5910591 0.6540233
```

```
## Ventilated_Death  0.076094461 0.3065705 -0.5132843 0.7855648
## AA_Discharged     0.257859069 0.2260377 -0.0733782 0.7393680
## Oxygen_AA         0.034113279 0.1808163 -0.3063976 0.4281857
## Ventilated_Oxygen 0.032197816 0.2929461 -0.5429803 0.6941894
## AA_Oxygen         0.161256290 0.2415409 -0.2288742 0.7153399
## Oxygen_Ventilated 0.055268444 0.3054040 -0.5271629 0.7524016
##
## , , took_favipiravir
##
##                      Estimate Est.Error       Q2.5     Q97.5
## Oxygen_Death      -0.17812284 0.7565367 -2.0388845 1.0444945
## Ventilated_Death  -0.32946788 0.7158238 -2.2446200 0.7076996
## AA_Discharged      0.47109035 0.4458086 -0.2006253 1.4241348
## Oxygen_AA          0.12129265 0.3342282 -0.5341281 0.8240631
## Ventilated_Oxygen  0.02561385 0.5750298 -1.2429200 1.2453903
## AA_Oxygen          0.17239683 0.5023469 -0.7813866 1.3242993
## Oxygen_Ventilated -0.16480453 0.5557129 -1.4783587 0.8137536
##
## , , took_convalescent_plasma
##
##                       Estimate Est.Error       Q2.5     Q97.5
## Oxygen_Death      -0.130661173 0.6451339 -1.7445077 1.0217083
## Ventilated_Death  -0.159791004 0.5075368 -1.4132437 0.7001265
## AA_Discharged      0.165037973 0.4280145 -0.6135059 1.1311555
## Oxygen_AA         -0.264587323 0.3832700 -1.1676273 0.3348971
## Ventilated_Oxygen -0.009238966 0.4689791 -1.0795350 1.0049055
## AA_Oxygen         -0.266436991 0.6704849 -2.1420892 0.7136004
## Oxygen_Ventilated  0.177207717 0.5167253 -0.7562704 1.3928378
##
## , , age_norm
##
##                      Estimate Est.Error       Q2.5       Q97.5
## Oxygen_Death       0.17486900 0.2747240 -0.2999521  0.78614213
## Ventilated_Death   0.49546884 0.5198531 -0.2498284  1.83583875
## AA_Discharged     -0.40696146 0.1203356 -0.6382027 -0.16193180
## Oxygen_AA         -0.25550903 0.1423311 -0.5395766  0.02317489
## Ventilated_Oxygen -0.23617097 0.4009956 -1.1558875  0.47436383
## AA_Oxygen          0.14088965 0.1459314 -0.1336113  0.43350703
## Oxygen_Ventilated -0.07632298 0.3103216 -0.6906774  0.53109418
##
## , , is_male
##
##                        Estimate Est.Error       Q2.5      Q97.5
## Oxygen_Death      -0.0008886269 0.2211279 -0.4564423 0.49634153
## Ventilated_Death  -0.1128074098 0.3022539 -0.8588735 0.39919180
## AA_Discharged     -0.1973241153 0.1789589 -0.5824407 0.08314512
## Oxygen_AA         -0.1669185995 0.1737022 -0.5592952 0.11035625
## Ventilated_Oxygen -0.0690256955 0.2838501 -0.7497127 0.45967245
## AA_Oxygen         -0.0139229158 0.1625966 -0.3675874 0.30892378
## Oxygen_Ventilated -0.1562128261 0.2968567 -0.9068018 0.30406280
##
## , , Intercept
##
##                      Estimate Est.Error      Q2.5    Q97.5
```

```
## Oxygen_Death      -0.27721980  1.264851 -3.226755 2.200145
## Ventilated_Death  -0.12527721  1.310545 -3.107099 2.757234
## AA_Discharged     -0.01976889  1.236418 -2.706491 2.583650
## Oxygen_AA          0.01776315  1.134570 -2.492704 2.502678
## Ventilated_Oxygen -0.11318032  1.266219 -3.062307 2.440703
## AA_Oxygen         -0.11209423  1.206086 -2.946750 2.366909
## Oxygen_Ventilated -0.15272612  1.343860 -3.347977 2.551297
##
## , , comorbidities_sum
##
##                       Estimate  Est.Error        Q2.5      Q97.5
## Oxygen_Death       0.032235109 0.07706698 -0.09932803 0.20764110
## Ventilated_Death   0.034829883 0.09680026 -0.12742382 0.28006310
## AA_Discharged     -0.056671492 0.05618073 -0.18054290 0.02214423
## Oxygen_AA         -0.005097332 0.04941716 -0.11324848 0.09096859
## Ventilated_Oxygen  0.003309318 0.09206774 -0.19058492 0.21215330
## AA_Oxygen         -0.008173292 0.04889598 -0.11253917 0.09657224
## Oxygen_Ventilated  0.085742550 0.12590118 -0.05833219 0.42518513
##
##
## Between-site differences:
##                                Estimate Est.Error       Q2.5    Q97.5
## sd(.rate_idOxygen_Death)      1.2755505 0.5360095 0.50238070 2.613885
## sd(.rate_idVentilated_Death)  0.9405574 0.7190523 0.03373263 2.771543
## sd(.rate_idAA_Discharged)     1.4660463 0.5522342 0.55656030 2.708392
## sd(.rate_idOxygen_AA)         0.7571247 0.3552808 0.26877175 1.624937
## sd(.rate_idVentilated_Oxygen) 0.6885347 0.5813338 0.03216758 2.146322
## sd(.rate_idAA_Oxygen)         1.5952085 0.5676666 0.77712123 2.991458
## sd(.rate_idOxygen_Ventilated) 2.3420957 0.9715424 0.49322490 4.486446
```

Log odds-ratio estimates:

```
##                        hypothesis point_estimate 95% CI Low  95% CI High
## 1                       hcq_death    -0.46102387 -1.0545742 -0.006623639
## 2                    hcq_hospital    -0.50480808 -1.1130403  0.083773838
## 3         hcq_hospital (duration)    -0.04283602 -0.1757391  0.093607033
## 4                        az_death     0.00146455 -0.2780391  0.289795207
## 5                     az_hospital    -0.24348145 -0.8176950  0.140142042
## 6          az_hospital (duration)    -0.07550106 -0.2289876  0.017212164
## 7               favipiravir_death    -0.20028512 -1.1014467  0.278425437
## 8            favipiravir_hospital    -0.39393568 -1.4433875  0.342658505
## 9 favipiravir_hospital (duration)    -0.14839078 -0.5199554  0.051875937
```

#### 5.4 Markers

In the marker models, we use the simple model and investigate effects on rate groups. We filter only for patients that at had the marker measured at least once. We use the peak of the marker so far as a predictor in the model and investigate its estimated coefficient. Unfortunately, none of the results are trustworthy as the sampler encountered divergent transitions.

##### 5.4.1 D-dimer + hospital

The formula is:

```
## ~0 + .rate_id + (0 + best_supportive_care || rate_group) + (0 +
##     .rate_id | hospital_id) + (log_peak_d_dimer || rate_group)
```

And here are the coefficient estimates - note the divergent transitions, indicating the results are not completely trustworthy:

```
## Warning: There were 35 divergent transitions after warmup. Increasing adapt_delta above may help. Se
## transitions-after-warmup
```

```
##
## Rate coefficients:
##                            Estimate Est.Error      Q2.5      Q97.5
## .rate_idOxygen_Death      -5.686684  1.681796 -8.752808 -1.9845427
## .rate_idVentilated_Death  -5.662366  1.771079 -8.930380 -1.8014360
## .rate_idAA_Discharged     -2.084210  1.447450 -4.939433  1.1522155
## .rate_idOxygen_AA         -1.977260  1.301413 -4.682300  1.1874173
## .rate_idVentilated_Oxygen -3.357165  1.399299 -6.279436 -0.2091585
## .rate_idAA_Oxygen         -2.259000  1.497885 -5.443232  1.0549093
## .rate_idOxygen_Ventilated -4.119981  1.691360 -7.569851 -0.3862285
##
## Per rate group effects:
## , , best_supportive_care
##
##              Estimate Est.Error     Q2.5      Q97.5
## improve_one -0.628935 0.3996689 -1.48703 0.08110057
## death        3.263292 0.5987387  2.19123 4.46408100
## worsen_one  -0.992466 1.0135280 -3.05148 0.94321065
##
## , , Intercept
##
##               Estimate Est.Error      Q2.5    Q97.5
## improve_one -0.1903327  1.289654 -3.349236 2.498886
## death       -0.5469543  1.449655 -4.281889 1.807992
## worsen_one  -0.1891857  1.373464 -3.362560 2.692680
##
## , , log_peak_d_dimer
##
##                Estimate  Est.Error        Q2.5      Q97.5
## improve_one -0.04602089 0.04149957 -0.12867910 0.02876492
## death       -0.15747827 0.11998038 -0.43465340 0.02636271
## worsen_one   0.12693012 0.05379901  0.01698733 0.22800403
##
##
## Between-site differences:
##                                Estimate Est.Error       Q2.5    Q97.5
## sd(.rate_idOxygen_Death)      1.7251438 0.6686232 0.74323383 3.363668
## sd(.rate_idVentilated_Death)  1.4352315 0.9329348 0.10557360 3.566617
## sd(.rate_idAA_Discharged)     1.9716891 0.6704624 0.99256180 3.571352
## sd(.rate_idOxygen_AA)         0.3898263 0.2766182 0.02872911 1.094586
## sd(.rate_idVentilated_Oxygen) 0.7402473 0.7070797 0.02143050 2.584072
## sd(.rate_idAA_Oxygen)         1.2221890 0.6527602 0.24493965 2.718367
## sd(.rate_idOxygen_Ventilated) 2.0717461 1.1854526 0.15466283 4.590559
```

##### 5.4.2 D-dimer + age + sex + comorbidities

Including comorbidites in the model gives the formula:

```
## ~.rate_id + (0 + best_supportive_care || rate_group) + (0 + .rate_id |
##     hospital_id) + (log_peak_d_dimer || rate_group) + (0 + age_norm +
##     is_male || rate_group) - 1
```

Where `comorbidities_sum` is a comorbidity score as described in Section 1.1.

And here are the coefficient estimates - note the divergent transitions, indicating the results are not completely trustworthy:

```
## Warning: There were 51 divergent transitions after warmup. Increasing adapt_delta above may help. Se
## transitions-after-warmup
```

```
##
## Rate coefficients:
##                            Estimate Est.Error      Q2.5      Q97.5
## .rate_idOxygen_Death      -5.786380  1.728314 -8.882839 -1.8114747
## .rate_idVentilated_Death  -5.591031  1.783587 -8.814782 -1.4510245
## .rate_idAA_Discharged     -2.138416  1.331188 -4.723346  0.7102087
## .rate_idOxygen_AA         -1.961496  1.155860 -4.261095  0.5609019
## .rate_idVentilated_Oxygen -3.375181  1.264702 -5.948095 -0.6515390
## .rate_idAA_Oxygen         -2.235513  1.400131 -5.025286  0.9797593
## .rate_idOxygen_Ventilated -4.177598  1.580615 -7.354728 -0.8425036
##
## Per rate group effects:
## , , best_supportive_care
##
##               Estimate Est.Error      Q2.5     Q97.5
## improve_one -0.5372003 0.4107315 -1.380835 0.2308639
## death        3.1071974 0.6367095  1.932140 4.4241250
## worsen_one  -1.0703833 1.0275331 -3.052311 0.9455844
##
## , , Intercept
##
##                Estimate Est.Error      Q2.5    Q97.5
## improve_one -0.09974711  1.115248 -2.533432 2.126199
## death       -0.58787190  1.465290 -4.413249 1.677610
## worsen_one  -0.19278994  1.274491 -3.283183 2.420785
##
## , , log_peak_d_dimer
##
##                Estimate  Est.Error        Q2.5      Q97.5
## improve_one -0.04813276 0.04114208 -0.13089685 0.02820987
## death       -0.15689851 0.11702691 -0.42593060 0.02350400
## worsen_one   0.13105543 0.05453345  0.01246391 0.23253645
##
## , , age_norm
##
##               Estimate Est.Error       Q2.5       Q97.5
## improve_one -0.1954312 0.1086113 -0.4140918 0.006305619
## death        0.2607975 0.3316756 -0.2164683 1.070904500
```

```
## worsen_one   0.1319566 0.1600189 -0.1467335 0.475408200
##
## , , is_male
##
##                Estimate Est.Error       Q2.5      Q97.5
## improve_one -0.14199609 0.1496371 -0.4753307 0.09237117
## death        0.01287642 0.2608843 -0.5197301 0.63202628
## worsen_one   0.04156971 0.1827565 -0.3271852 0.43992663
##
##
## Between-site differences:
##                                Estimate Est.Error       Q2.5    Q97.5
## sd(.rate_idOxygen_Death)      1.7418145 0.6769936 0.74483243 3.354283
## sd(.rate_idVentilated_Death)  1.3559212 0.9278030 0.07073830 3.660373
## sd(.rate_idAA_Discharged)     1.9367455 0.6641094 0.93941447 3.517571
## sd(.rate_idOxygen_AA)         0.4892701 0.3421394 0.04410227 1.314289
## sd(.rate_idVentilated_Oxygen) 0.7300325 0.6855340 0.02908739 2.552338
## sd(.rate_idAA_Oxygen)         1.1886254 0.6713604 0.17299803 2.835810
## sd(.rate_idOxygen_Ventilated) 2.0845851 1.1728739 0.15790025 4.661818
```

##### 5.4.3 IL-6 + hospital

We further look into Interleukin 6 using the formula:

```
## ~0 + .rate_id + (0 + best_supportive_care || rate_group) + (0 +
##     .rate_id | hospital_id) + (log_peak_IL_6 || rate_group)
```

Giving us the following coefficients - note the divergent transitions, indicating the results are not completely trustworthy:

```
## Warning: There were 8 divergent transitions after warmup. Increasing adapt_delta above may help. See
## transitions-after-warmup
```

```
##
## Rate coefficients:
##                            Estimate Est.Error      Q2.5       Q97.5
## .rate_idOxygen_Death      -4.935438  1.736411 -8.618153 -1.41896450
## .rate_idVentilated_Death  -3.519498  1.580666 -6.544897  0.02109801
## .rate_idAA_Discharged     -2.285582  1.206916 -4.770434  0.37043028
## .rate_idOxygen_AA         -1.762646  1.341674 -4.402457  1.17979025
## .rate_idVentilated_Oxygen -3.399736  1.345279 -6.082186 -0.57450127
## .rate_idAA_Oxygen         -2.145116  1.384053 -4.758182  1.27804700
## .rate_idOxygen_Ventilated -4.610499  1.602632 -7.914137 -1.07673000
##
## Per rate group effects:
## , , best_supportive_care
##
##              Estimate Est.Error      Q2.5      Q97.5
## improve_one -1.162006 0.7143182 -2.738502 0.02780235
## death        1.659778 1.0588241 -0.370437 3.78476825
## worsen_one  -2.219944 1.7737369 -6.370176 0.28937438
##
```

```
## , , Intercept
##
##               Estimate Est.Error      Q2.5    Q97.5
## improve_one -0.1055650  1.134484 -2.709342 2.196629
## death       -0.2951620  1.401003 -3.492336 2.450826
## worsen_one  -0.2122986  1.286219 -3.419607 2.240803
##
## , , log_peak_IL_6
##
##               Estimate Est.Error       Q2.5        Q97.5
## improve_one -0.2313676 0.1268443 -0.5074526 -0.001749414
## death       -0.8074576 0.9010183 -3.3065727  0.021600240
## worsen_one   0.1709045 0.1422230 -0.1140742  0.436010000
##
##
## Between-site differences:
##                                Estimate Est.Error       Q2.5    Q97.5
## sd(.rate_idOxygen_Death)      1.1984340 0.9143703 0.06606922 3.444852
## sd(.rate_idVentilated_Death)  0.8336388 0.7521870 0.03038907 2.810832
## sd(.rate_idAA_Discharged)     0.7694387 0.5566954 0.08236453 2.224663
## sd(.rate_idOxygen_AA)         1.2330010 0.8079948 0.13481373 3.185192
## sd(.rate_idVentilated_Oxygen) 0.9167199 0.7798884 0.04005720 2.912383
## sd(.rate_idAA_Oxygen)         0.7302227 0.7266465 0.01958305 2.675232
## sd(.rate_idOxygen_Ventilated) 1.1014647 0.9085542 0.04860189 3.393584
```

##### 5.4.4 IL_6 + age + sex + comorbidities

And finally adding comorbidities with the formula:

```
## ~.rate_id + (0 + best_supportive_care || rate_group) + (0 + .rate_id |
##     hospital_id) + (log_peak_IL_6 || rate_group) + (0 + age_norm +
##     is_male || rate_group) - 1
```

Where `comorbidities_sum` is a comorbidity score as described in Section 1.1.

Gives us those estimates - note the divergent transitions, indicating the results are not completely trustworthy:

```
## Warning: There were 51 divergent transitions after warmup. Increasing adapt_delta above may help. Se
## transitions-after-warmup
```

```
##
## Rate coefficients:
##                            Estimate Est.Error      Q2.5      Q97.5
## .rate_idOxygen_Death      -5.871774  1.989303 -9.972071 -1.4895877
## .rate_idVentilated_Death  -3.734294  1.727788 -6.765419  0.2363629
## .rate_idAA_Discharged     -2.209650  1.261348 -4.816824  0.6048484
## .rate_idOxygen_AA         -1.644860  1.360893 -4.516631  1.2861670
## .rate_idVentilated_Oxygen -3.269354  1.382249 -6.131176 -0.2668680
## .rate_idAA_Oxygen         -2.317448  1.636550 -5.421046  1.3604913
## .rate_idOxygen_Ventilated -4.841171  1.682046 -8.301401 -1.2049985
##
## Per rate group effects:
```

```
## , , best_supportive_care
##
##               Estimate Est.Error       Q2.5     Q97.5
## improve_one -0.796892 0.7176621 -2.4445595 0.2765177
## death        1.112679 1.0773408 -0.6517291 3.3530443
## worsen_one  -1.859738 1.8075064 -6.4521995 0.4764803
##
## , , Intercept
##
##                Estimate Est.Error      Q2.5    Q97.5
## improve_one -0.1553079  1.167868 -2.901281 2.319167
## death       -0.4761043  1.472594 -4.105513 2.063560
## worsen_one  -0.2751205  1.404256 -3.737125 2.385616
##
## , , log_peak_IL_6
##
##                Estimate Est.Error       Q2.5       Q97.5
## improve_one -0.2474300 0.1277285 -0.5283995 -0.02734142
## death       -0.7423145 0.8506601 -3.0125130  0.03221664
## worsen_one   0.1814508 0.1404321 -0.1108498  0.43481267
##
## , , age_norm
##
##                Estimate Est.Error        Q2.5       Q97.5
## improve_one -0.2992318 0.1433122 -0.58367217 -0.01703348
## death        1.4669714 0.8599029  0.03676329  3.28415400
## worsen_one   0.6418314 0.3200053  0.08090958  1.31276450
##
## , , is_male
##
##                 Estimate Est.Error       Q2.5     Q97.5
## improve_one -0.08696444 0.1713591 -0.4718728 0.2202813
## death       -0.03542763 0.3447884 -0.8542363 0.7145321
## worsen_one   0.08216941 0.2244197 -0.3361775 0.5760288
##
##
## Between-site differences:
##                                 Estimate Est.Error       Q2.5    Q97.5
## sd(.rate_idOxygen_Death)      1.1168974 0.9027111 0.05233202 3.292391
## sd(.rate_idVentilated_Death)  0.8544930 0.7337388 0.02379513 2.704761
## sd(.rate_idAA_Discharged)     0.6241296 0.5132422 0.02724517 2.036218
## sd(.rate_idOxygen_AA)         1.2625284 0.8289799 0.19292380 3.349247
## sd(.rate_idVentilated_Oxygen) 0.9316523 0.8089931 0.02668968 3.018260
## sd(.rate_idAA_Oxygen)         1.3627786 1.0270817 0.04842824 3.870647
## sd(.rate_idOxygen_Ventilated) 1.2721222 0.9180428 0.04502789 3.541438
```

##### 5.5 Complex: Treatments - effects on rate groups

Here we use the complex model and let treatments have effect on the rate groups. The models have the same formulae as the simple models, only the rate (and state) definitions change. The rates used are shown in Table 2.

Table 2: Rates used in the complex model.

| .from | .to | rate_group | .rate_id |
|---|---|---|---|
| Oxygen_worsening | Death | death | Oxygen_worsening_Death |
| Ventilated_worsening | Death | death | Ventilated_worsening_Death |
| AA_improving | Discharged | improve_one | AA_improving_Discharged |
| Oxygen_improving | AA_improving | improve_one | Oxygen_improving_AA_improving |
| Ventilated_improving | Oxygen_improving | improve_one | Ventilated_improving_Oxygen_improving |
| AA_worsening | Oxygen_worsening | worsen_one | AA_worsening_Oxygen_worsening |
| Oxygen_worsening | Ventilated_worsening | worsen_one | Oxygen_worsening_Ventilated_worsening |
| AA_worsening | AA_improving | to_improving | AA_worsening_AA_improving |
| Oxygen_worsening | Oxygen_improving | to_improving | Oxygen_worsening_Oxygen_improving |
| Ventilated_worsening | Ventilated_improving | to_improving | Ventilated_worsening_Ventilated_improving |
| AA_improving | AA_worsening | to_worsening | AA_improving_AA_worsening |
| Oxygen_improving | Oxygen_worsening | to_worsening | Oxygen_improving_Oxygen_worsening |
| Ventilated_improving | Ventilated_worsening | to_worsening | Ventilated_improving_Ventilated_worsening |

##### 5.5.1 Only treatments

Using treatments only with the formula:

```
## ~0 + .rate_id + (0 + best_supportive_care + took_hcq + took_az +
##     took_favipiravir + took_convalescent_plasma || rate_group)
```

Fitted coefficients:

```
##
## Rate coefficients:
##                                                   Estimate Est.Error       Q2.5      Q97.5
## .rate_idOxygen_worsening_Death                  -2.8011057 1.5220407  -4.7502910  1.2135365
## .rate_idVentilated_worsening_Death              -3.3325593 0.8917613  -4.4554887 -0.9935105
## .rate_idAA_improving_Discharged                 -0.2446059 0.5968957  -1.1919742  1.1643825
## .rate_idOxygen_improving_AA_improving           -1.7191746 0.5417125  -2.3996665 -0.4360857
## .rate_idVentilated_improving_Oxygen_improving   -2.2664370 1.2003228  -3.6604325 -0.3625244
## .rate_idAA_worsening_Oxygen_worsening           -2.4729954 0.1691224  -2.8087375 -2.1293847
## .rate_idOxygen_worsening_Ventilated_worsening   -2.3424824 1.4994974  -4.0764045  1.6885943
## .rate_idAA_worsening_AA_improving               -1.2578009 0.3988031  -1.9997935 -0.4178084
## .rate_idOxygen_worsening_Oxygen_improving        2.3908010 2.0954389  -0.4016284  7.4928530
## .rate_idVentilated_worsening_Ventilated_improving -2.2970691 2.1122054  -4.1137977  4.6104888
## .rate_idAA_improving_AA_worsening                0.9772228 0.7764428  -0.2157075  2.8458723
## .rate_idOxygen_improving_Oxygen_worsening        1.1991217 1.7237415  -1.0515292  5.5846700
## .rate_idVentilated_improving_Ventilated_worsening -3.8190063 3.2216145 -10.9012900  2.7665953
##
## Per rate group effects:
## , , best_supportive_care
##
##                Estimate Est.Error        Q2.5     Q97.5
## improve_one  -0.4666020 0.4027144 -1.40699150 0.2370365
## death         3.1630218 0.6314680  2.05496375 4.4573740
## to_improving  2.0052383 1.3751113  0.03884126 5.5351253
## to_worsening  1.5066502 1.3144288 -0.50225337 4.9392587
```

```
## worsen_one   -0.3585449 0.3916631 -1.12156975 0.4246911
##
## , , took_hcq
##
##                 Estimate Est.Error       Q2.5     Q97.5
## improve_one  -0.06529555 0.2577387 -0.6551664 0.3805429
## death        -0.51149557 0.3944402 -1.3603870 0.1158783
## to_improving  0.26561590 0.3415812 -0.3554500 0.9834590
## to_worsening -0.10349088 0.3906632 -0.9133417 0.7092998
## worsen_one   -0.29863627 0.2445028 -0.8212678 0.1135969
##
## , , took_az
##
##                 Estimate Est.Error       Q2.5     Q97.5
## improve_one   0.14696769 0.1838623 -0.1943123 0.5333071
## death        -0.03190590 0.2704141 -0.6190071 0.5016246
## to_improving  0.14205101 0.2565600 -0.3222712 0.7208123
## to_worsening -0.08594225 0.2744886 -0.7048421 0.4522178
## worsen_one    0.17539029 0.2353470 -0.1991987 0.7181398
##
## , , took_favipiravir
##
##                 Estimate Est.Error      Q2.5     Q97.5
## improve_one  -0.34359485 0.4478565 -1.333038 0.3679446
## death        -0.81800751 1.6847104 -5.061972 1.7192795
## to_improving  0.05792143 1.2258370 -2.435275 2.5812600
## to_worsening -2.16727551 1.8924174 -6.324574 0.5053806
## worsen_one    1.29656904 1.0651028 -0.359415 3.3628880
##
## , , took_convalescent_plasma
##
##                 Estimate Est.Error       Q2.5     Q97.5
## improve_one  -0.43652498 0.4073780 -1.3507255 0.2043995
## death         0.03684769 0.6599066 -1.4873490 1.4270315
## to_improving -0.20199471 0.6795431 -1.8927037 0.9857279
## to_worsening -0.35628598 1.1147200 -3.6117353 1.0922263
## worsen_one    0.48745396 1.0144595 -0.7531574 3.3641818
```

Estimates:

```
##                       hypothesis point_estimate  95% CI Low  95% CI High
## 1                      hcq_death   -0.698893637 -1.39232033 -0.025068480
## 2                   hcq_hospital   -0.303820569 -0.97087078  0.315636141
## 3        hcq_hospital (duration)   -0.001460008 -0.09660221  0.100857697
## 4                       az_death   -0.212494210 -0.73975276  0.206097910
## 5                    az_hospital   -0.457986608 -1.15761335  0.114576464
## 6         az_hospital (duration)   -0.083307556 -0.20330216  0.007936137
## 7              favipiravir_death   -1.535872363 -5.89254256  0.641789838
## 8           favipiravir_hospital   -0.278904579 -1.76275564  1.119431572
## 9 favipiravir_hospital (duration)   -0.103403333 -0.47069146  0.192273660
```

##### 5.5.2 Treatments + age + sex

Adding age and sex with formula:

```
## ~.rate_id + (0 + best_supportive_care + took_hcq + took_az +
##     took_favipiravir + took_convalescent_plasma || rate_group) +
##     (0 + age_norm + is_male || rate_group) - 1
```

Fitted coefficients:

```
##
## Rate coefficients:
##                                                      Estimate Est.Error        Q2.5      Q97.5
## .rate_idOxygen_worsening_Death                     -2.9248140 1.5514559  -4.9798520  1.1600263
## .rate_idVentilated_worsening_Death                 -3.2287712 1.2413520  -4.6048107  0.2399800
## .rate_idAA_improving_Discharged                    -0.2173491 0.6643947  -1.3205547  1.1670805
## .rate_idOxygen_improving_AA_improving              -1.5480580 0.5672526  -2.2284440 -0.2814364
## .rate_idVentilated_improving_Oxygen_improving      -1.9713217 1.6355945  -3.6150780  2.5287780
## .rate_idAA_worsening_Oxygen_worsening              -2.4596386 0.2245430  -2.8754215 -2.0099662
## .rate_idOxygen_worsening_Ventilated_worsening      -2.3284530 1.5161109  -4.1336090  1.8623098
## .rate_idAA_worsening_AA_improving                  -1.1942485 0.4077653  -1.9365168 -0.3527128
## .rate_idOxygen_worsening_Oxygen_improving           2.0021174 1.9929519  -0.6329437  6.9144303
## .rate_idVentilated_worsening_Ventilated_improving -1.5310033 2.7888109  -4.1274595  6.3949765
## .rate_idAA_improving_AA_worsening                   0.6826421 0.8699886  -0.7445479  2.5099313
## .rate_idOxygen_improving_Oxygen_worsening           0.4607750 1.5154737  -1.5320158  4.8499623
## .rate_idVentilated_improving_Ventilated_worsening -2.5614044 3.6579069 -10.6322925  4.9171470
##
## Per rate group effects:
## , , best_supportive_care
##
##                Estimate Est.Error       Q2.5     Q97.5
## improve_one  -0.1661703 0.3879809 -0.9225606 0.5443709
## death         2.6090518 0.5934805  1.4979300 3.8784520
## to_improving  1.1877077 1.0536997 -0.3516382 3.7678393
## to_worsening  0.9420166 1.1231497 -0.8923870 3.6516270
## worsen_one   -0.5036897 0.3736056 -1.2727303 0.2066496
##
## , , took_hcq
##
##                Estimate Est.Error       Q2.5     Q97.5
## improve_one  -0.1243320 0.2869953 -0.7740559 0.3678138
## death        -0.4838828 0.3936454 -1.3507170 0.1710815
## to_improving  0.4540906 0.3754696 -0.1878885 1.2895425
## to_worsening  0.0342680 0.4540029 -0.7810468 1.0269555
## worsen_one   -0.3274113 0.2535616 -0.8553957 0.1281431
##
## , , took_az
##
##                 Estimate Est.Error       Q2.5     Q97.5
## improve_one   0.07909821 0.1750943 -0.2523629 0.4790798
## death        -0.02198240 0.2480810 -0.5923864 0.4908062
## to_improving  0.09956398 0.2484748 -0.3501448 0.7098204
## to_worsening -0.01122418 0.2804075 -0.5997408 0.6020301
## worsen_one    0.19311483 0.2396621 -0.1403568 0.7734866
##
## , , took_favipiravir
##
##                Estimate Est.Error       Q2.5     Q97.5
```

```
## improve_one  -0.2102632 0.3997395 -1.1037960 0.4611888
## death        -0.9037579 1.7715682 -5.5419352 1.7620075
## to_improving -0.1934976 1.1277453 -2.6984260 1.9827355
## to_worsening -2.3374697 1.9466470 -6.5134200 0.2812808
## worsen_one    1.3614269 1.1003095 -0.3396947 3.5385583
##
## , , took_convalescent_plasma
##
##                 Estimate Est.Error       Q2.5     Q97.5
## improve_one  -0.46367402 0.4239742 -1.4149800 0.2251099
## death         0.07811338 0.7505283 -1.5679885 1.6732223
## to_improving -0.34090426 0.9007881 -2.5583698 1.0867813
## to_worsening -0.53935693 1.4496873 -4.6958493 1.2011262
## worsen_one    0.66614760 1.1943282 -0.7942403 4.0285240
##
## , , age_norm
##
##                 Estimate Est.Error       Q2.5      Q97.5
## improve_one  -0.28676123 0.1728067 -0.6349286 0.04669976
## death         0.47748570 0.3502603 -0.1096538 1.21616275
## to_improving  0.09633123 0.2594560 -0.3781483 0.67089623
## to_worsening  0.42934229 0.3328211 -0.1264738 1.18153075
## worsen_one    0.17565885 0.1507426 -0.1041732 0.48088050
##
## , , is_male
##
##                 Estimate Est.Error       Q2.5      Q97.5
## improve_one  -0.25033711 0.1857885 -0.6623992 0.03566686
## death        -0.05443236 0.2247945 -0.5349350 0.38968170
## to_improving -0.09285905 0.2286320 -0.6143771 0.34753168
## to_worsening -0.03701985 0.2731237 -0.6994239 0.44682000
## worsen_one   -0.04051842 0.1676326 -0.3803358 0.29286128
```

Estimates:

```
##                       hypothesis point_estimate 95% CI Low 95% CI High
## 1                      hcq_death   -0.674543740 -1.2726695 -0.12692807
## 2                   hcq_hospital   -0.200286255 -0.8204069  0.38776696
## 3        hcq_hospital (duration)   -0.003375891 -0.1092269  0.10398342
## 4                       az_death   -0.062428185 -0.4934511  0.34323557
## 5                    az_hospital   -0.180772898 -0.7725520  0.31467540
## 6         az_hospital (duration)   -0.048662816 -0.1678519  0.03112189
## 7              favipiravir_death   -1.371056798 -5.9219056  0.69318701
## 8           favipiravir_hospital   -0.216326449 -1.6963201  1.18916487
## 9 favipiravir_hospital (duration)   -0.089075302 -0.4465107  0.21359607
```

##### 5.5.3 Treatments + age + sex + hospital

Including hospital as well with the formula:

```
## ~.rate_id + (0 + best_supportive_care + took_hcq + took_az +
##     took_favipiravir + took_convalescent_plasma || rate_group) +
##     (0 + age_norm + is_male || rate_group) + (0 + .rate_id |
##     hospital_id) - 1
```

Fitted coefficients:

```
## Warning: There were 1 divergent transitions after warmup. Increasing adapt_delta above may help. See
## transitions-after-warmup

##
## Rate coefficients:
##                                                   Estimate Est.Error         Q2.5      Q97.5
## .rate_idOxygen_worsening_Death                  -3.3457077 1.7680626  -6.36770425  0.6176781
## .rate_idVentilated_worsening_Death              -3.2125522 1.9452179  -5.98855575  1.9884835
## .rate_idAA_improving_Discharged                 -1.5364634 0.6014110  -2.65929650 -0.2211763
## .rate_idOxygen_improving_AA_improving           -1.5513041 0.7621594  -2.39838950 -0.3653768
## .rate_idVentilated_improving_Oxygen_improving   -1.6739196 2.5214647  -4.24737350  6.0196215
## .rate_idAA_worsening_Oxygen_worsening           -1.6373921 0.5940806  -2.88473025 -0.5084440
## .rate_idOxygen_worsening_Ventilated_worsening   -2.5466386 1.8340885  -5.53556450  1.9275043
## .rate_idAA_worsening_AA_improving               -0.1830593 0.9174307  -1.63685325  1.9466985
## .rate_idOxygen_worsening_Oxygen_improving        2.9692892 2.1422544  -0.04552768  8.1342763
## .rate_idVentilated_worsening_Ventilated_improving -0.9634564 3.3132433  -4.42024475  7.4292188
## .rate_idAA_improving_AA_worsening               -1.8997077 1.1350877  -4.17494700  0.2966953
## .rate_idOxygen_improving_Oxygen_worsening        0.4573909 1.8764244  -2.19079025  5.3072038
## .rate_idVentilated_improving_Ventilated_worsening -3.3232477 3.7121932 -11.62597000  3.4717655
##
## Per rate group effects:
## , , best_supportive_care
##
##                 Estimate Est.Error       Q2.5    Q97.5
## improve_one  0.02648498 0.4431107 -0.7657004 1.025213
## death        2.83834342 0.7896966  1.4889948 4.558967
## to_improving 0.20058875 1.0781643 -1.7114900 2.571180
## to_worsening 1.40335182 0.9966471 -0.3844644 3.496062
## worsen_one   0.62487144 0.6957639 -0.6587533 2.081048
##
## , , took_hcq
##
##                  Estimate Est.Error       Q2.5     Q97.5
## improve_one   0.04173776 0.2388077 -0.3999859 0.5638985
## death        -0.37003650 0.4270902 -1.3830852 0.2795503
## to_improving  0.19885632 0.3778831 -0.4737580 1.0491325
## to_worsening -0.09455076 0.5232793 -1.0360798 1.2583012
## worsen_one   -0.13522809 0.3876533 -1.1076885 0.5163693
##
## , , took_az
##
##                  Estimate Est.Error        Q2.5     Q97.5
## improve_one   0.19204424 0.1884906 -0.09479205 0.5947244
## death         0.06047344 0.2825811 -0.47497873 0.6830322
## to_improving -0.01737463 0.2692653 -0.59171632 0.5542831
## to_worsening  0.08107206 0.3002278 -0.49268952 0.7839257
## worsen_one    0.08785209 0.2622044 -0.39812670 0.6859433
##
## , , took_favipiravir
##
##                  Estimate Est.Error       Q2.5     Q97.5
## improve_one   0.24617500 0.4204936 -0.6324374 1.0872747
```

```
## death        -0.55981150 1.1333195 -3.5178272 0.9912665
## to_improving -0.06325144 0.7466014 -1.8481018 1.3081023
## to_worsening -0.76766311 1.3522221 -4.4875540 0.7761913
## worsen_one    0.44567735 0.7604263 -0.6524054 2.2955313
##
## , , took_convalescent_plasma
##
##                Estimate Est.Error       Q2.5     Q97.5
## improve_one  -0.2166006 0.3821986 -1.0912195 0.3674980
## death        -0.1761091 0.6417822 -1.6852700 0.8769131
## to_improving -0.1053559 0.6857868 -1.5495190 1.0710033
## to_worsening -0.1604712 0.9814074 -2.9566313 1.2164870
## worsen_one    0.3073903 0.8149366 -0.7340391 2.6940278
##
## , , age_norm
##
##                 Estimate Est.Error       Q2.5       Q97.5
## improve_one  -0.37132507 0.1643435 -0.6938281 -0.02013974
## death         0.41259561 0.3897991 -0.1911011  1.32009775
## to_improving -0.08793716 0.2603217 -0.6359255  0.42850500
## to_worsening  0.09873341 0.3827572 -0.6774420  0.94552968
## worsen_one    0.17418756 0.2496782 -0.3432759  0.65391067
##
## , , is_male
##
##                 Estimate Est.Error       Q2.5      Q97.5
## improve_one  -0.21337104 0.1623229 -0.5496841 0.05399288
## death        -0.02804944 0.2517906 -0.5975649 0.47459118
## to_improving -0.15491107 0.2801531 -0.8441210 0.32506153
## to_worsening -0.01852471 0.2957749 -0.6992949 0.53846213
## worsen_one   -0.09926653 0.2294770 -0.6581773 0.30729990
##
##
## Between-site differences:
##                                                    Estimate Est.Error       Q2.5    Q97.5
## sd(.rate_idOxygen_worsening_Death)                1.2208364 0.7422819 0.11121325 3.009137
## sd(.rate_idVentilated_worsening_Death)            1.3299497 0.8216459 0.09238057 3.221577
## sd(.rate_idAA_improving_Discharged)               1.4252515 0.5637165 0.53570353 2.737330
## sd(.rate_idOxygen_improving_AA_improving)         0.7557002 0.4145837 0.13720853 1.786893
## sd(.rate_idVentilated_improving_Oxygen_improving) 0.8722083 0.8541053 0.02903513 3.221572
## sd(.rate_idAA_worsening_Oxygen_worsening)         0.8126704 0.5983436 0.03958624 2.265452
## sd(.rate_idOxygen_worsening_Ventilated_worsening) 2.0823642 1.0044061 0.29803603 4.280899
## sd(.rate_idAA_worsening_AA_improving)             1.3022277 0.7172009 0.17195380 2.937576
## sd(.rate_idOxygen_worsening_Oxygen_improving)     1.0605282 0.7603336 0.05295536 2.867806
## sd(.rate_idVentilated_worsening_Ventilated_improving) 0.9793489 0.8242693 0.03753788 3.078653
## sd(.rate_idAA_improving_AA_worsening)             2.0388853 0.9041092 0.36128303 3.983822
## sd(.rate_idOxygen_improving_Oxygen_worsening)     0.9098574 0.6968910 0.03036363 2.574595
## sd(.rate_idVentilated_improving_Ventilated_worsening) 1.3068708 1.0293888 0.04124696 3.911036
```

Estimates:

```
##                   hypothesis point_estimate 95% CI Low 95% CI High
## 1                  hcq_death   -0.364448739 -0.9444101  0.03585455
## 2               hcq_hospital   -0.143394072 -0.7543138  0.40623632
```

```
## 3        hcq_hospital (duration)    0.008278186 -0.1007723  0.12734414
## 4                      az_death    0.026667234 -0.3128180  0.39397300
## 5                   az_hospital   -0.249872229 -0.8432581  0.18637410
## 6         az_hospital (duration)   -0.061878533 -0.1929223  0.02308699
## 7              favipiravir_death   -0.563224044 -2.7845672  0.37366910
## 8           favipiravir_hospital   -0.370756275 -1.5794132  0.72573754
## 9 favipiravir_hospital (duration)   -0.079024696 -0.4035508  0.22359891
```

###### 5.5.4 Treatments + age + sex + hospital, first wave

The same model as above, but using only first wave patients. Fitted coefficients:

```
##
## Rate coefficients:
##                                                   Estimate Est.Error         Q2.5       Q97.5
## .rate_idOxygen_worsening_Death                  -3.7992080 1.4586612  -6.62034600 -0.62253780
## .rate_idVentilated_worsening_Death              -3.1724357 2.0453790  -5.89331800  2.45442750
## .rate_idAA_improving_Discharged                 -1.8041775 0.5988531  -2.95426275 -0.49274080
## .rate_idOxygen_improving_AA_improving           -1.8348122 0.4114496  -2.52364650 -0.91057285
## .rate_idVentilated_improving_Oxygen_improving   -1.7301115 2.5356479  -4.35264150  5.41067125
## .rate_idAA_worsening_Oxygen_worsening           -1.3635752 0.7080073  -2.91018625 -0.01201981
## .rate_idOxygen_worsening_Ventilated_worsening   -3.3441921 1.2987321  -5.64182150 -0.37129432
## .rate_idAA_worsening_AA_improving                0.5983139 1.1252163  -1.33785825  3.07730425
## .rate_idOxygen_worsening_Oxygen_improving        2.4488819 1.7980610  -0.09943714  6.80459150
## .rate_idVentilated_worsening_Ventilated_improving -1.0226250 3.2755290  -4.28600575  7.40224875
## .rate_idAA_improving_AA_worsening               -2.1183125 1.2438500  -4.69601925  0.22258545
## .rate_idOxygen_improving_Oxygen_worsening        0.1103539 1.7181021  -2.46961900  4.41371675
## .rate_idVentilated_improving_Ventilated_worsening -3.6702491 3.7849152 -11.88146250  3.28675750
##
## Per rate group effects:
## , , best_supportive_care
##
##                Estimate Est.Error       Q2.5    Q97.5
## improve_one   0.3451175 0.5345357 -0.5665884 1.509135
## death         2.5116870 0.7165074  1.2086292 4.040737
## to_improving -0.1624667 1.0492120 -1.9936698 2.134799
## to_worsening  1.6039104 1.0230280 -0.1596997 3.829607
## worsen_one    0.5621762 0.6899578 -0.7569026 1.921780
##
## , , took_hcq
##
##                 Estimate Est.Error       Q2.5     Q97.5
## improve_one   0.14861235 0.2622138 -0.2792686 0.8021607
## death        -0.46239161 0.4875248 -1.5574177 0.1971349
## to_improving  0.12289133 0.3934176 -0.6398229 1.0051850
## to_worsening  0.03319059 0.5820995 -0.9463529 1.5302783
## worsen_one   -0.22569341 0.4647464 -1.3540533 0.5081727
##
## , , took_az
##
##                 Estimate Est.Error        Q2.5     Q97.5
## improve_one   0.24305574 0.2044252 -0.07409251 0.6923382
## death         0.04971633 0.3079312 -0.61132325 0.7390486
```

```
## to_improving -0.06424495 0.3052390 -0.72976825 0.5648990
## to_worsening  0.15672457 0.3648828 -0.45917110 1.0404868
## worsen_one    0.10879694 0.2824199 -0.39767537 0.7481821
##
## , , took_favipiravir
##
##                 Estimate Est.Error       Q2.5     Q97.5
## improve_one   0.21508142 0.4326294 -0.7509071 1.0210550
## death        -0.60799356 1.0970755 -3.7871747 0.7383430
## to_improving -0.07702334 0.7336233 -1.7068212 1.2992955
## to_worsening -0.73038507 1.3496402 -4.7020840 0.7758429
## worsen_one    0.34274911 0.7077397 -0.8435814 2.1831390
##
## , , took_convalescent_plasma
##
##                Estimate Est.Error       Q2.5     Q97.5
## improve_one  -0.2476183 0.4117554 -1.2093443 0.4304566
## death        -0.1936122 0.6611017 -1.8174165 0.8813617
## to_improving -0.0942724 0.6590844 -1.5611605 1.0993550
## to_worsening -0.2155936 1.1053369 -3.5859700 1.2942788
## worsen_one    0.3288153 0.8563670 -0.8150719 2.7961645
##
## , , age_norm
##
##                 Estimate Est.Error       Q2.5      Q97.5
## improve_one  -0.30033258 0.1575897 -0.5851459 0.04365038
## death         0.35229342 0.3618677 -0.1903767 1.20924050
## to_improving -0.15516380 0.2668419 -0.7030080 0.37651295
## to_worsening  0.17329088 0.3867007 -0.5445281 1.02983375
## worsen_one    0.09185824 0.2627922 -0.4206736 0.58664340
##
## , , is_male
##
##                 Estimate Est.Error       Q2.5     Q97.5
## improve_one  -0.15441153 0.1539459 -0.4767641 0.1093294
## death        -0.01634454 0.2214315 -0.5240798 0.4159339
## to_improving -0.11835029 0.2609859 -0.7453792 0.3323907
## to_worsening  0.05597625 0.2565231 -0.4873509 0.6113818
## worsen_one   -0.08695559 0.2192292 -0.6425902 0.2954768
##
##
## Between-site differences:
##                                                       Estimate Est.Error       Q2.5    Q97.5
## sd(.rate_idOxygen_worsening_Death)                   1.3900037 0.7540813 0.21286508 3.269448
## sd(.rate_idVentilated_worsening_Death)               1.3254973 0.8457480 0.08218110 3.305770
## sd(.rate_idAA_improving_Discharged)                  1.5072219 0.5589219 0.63693830 2.884958
## sd(.rate_idOxygen_improving_AA_improving)            0.5119832 0.3511906 0.04261472 1.472361
## sd(.rate_idVentilated_improving_Oxygen_improving)    0.9063286 0.8408822 0.02515527 3.097122
## sd(.rate_idAA_worsening_Oxygen_worsening)            0.8472221 0.7062050 0.02963533 2.537675
## sd(.rate_idOxygen_worsening_Ventilated_worsening)    1.2568497 0.9767322 0.03668150 3.721257
## sd(.rate_idAA_worsening_AA_improving)                1.8449084 0.8426429 0.45351575 3.832430
## sd(.rate_idOxygen_worsening_Oxygen_improving)        1.1476102 0.7541073 0.07283942 2.952539
## sd(.rate_idVentilated_worsening_Ventilated_improving) 0.9549941 0.8462401 0.03084616 3.118087
## sd(.rate_idAA_improving_AA_worsening)                2.1449508 1.0017126 0.24206138 4.390733
```

```
## sd(.rate_idOxygen_improving_Oxygen_worsening)         0.9184481 0.7216616 0.03836311 2.675351
## sd(.rate_idVentilated_improving_Ventilated_worsening) 1.2779537 1.0168273 0.03542762 3.821026
```

Estimates:

```
##                        hypothesis point_estimate 95% CI Low 95% CI High
## 1                       hcq_death   -0.362926232 -0.9868015  0.04750491
## 2                    hcq_hospital   -0.215839393 -0.8377178  0.34818860
## 3         hcq_hospital (duration)   -0.004294224 -0.1161632  0.11436899
## 4                        az_death    0.060586196 -0.2934862  0.47190013
## 5                     az_hospital   -0.273345448 -0.8731011  0.19095203
## 6          az_hospital (duration)   -0.074946653 -0.2134063  0.01846581
## 7               favipiravir_death   -0.568970363 -3.0040485  0.27841789
## 8            favipiravir_hospital   -0.342079983 -1.5276130  0.79731307
## 9 favipiravir_hospital (duration)   -0.061583800 -0.3771753  0.25102164
```

##### 5.5.5 Treatment + age + sex + hospital + comorbidities

Adding comorbidities with the formula:

```
## ~.rate_id + (0 + best_supportive_care + took_hcq + took_az +
##     took_favipiravir + took_convalescent_plasma || rate_group) +
##     (0 + age_norm + is_male || rate_group) + (0 + .rate_id |
##     hospital_id) + (comorbidities_sum || rate_group) - 1
```

Where `comorbidities_sum` is a comorbidity score as described in Section 1.1.

Fitted coefficients - note the divergent transitions, indicating the results are not completely trustworthy:

```
## Warning: There were 25 divergent transitions after warmup. Increasing adapt_delta above may help. Se
## transitions-after-warmup
```

```
##
## Rate coefficients:
##                                                     Estimate Est.Error       Q2.5     Q97.5
## .rate_idOxygen_worsening_Death                    -3.3702592  2.152823  -7.510988 1.3422363
## .rate_idVentilated_worsening_Death                -3.2031656  2.400269  -7.144477 2.6463793
## .rate_idAA_improving_Discharged                   -1.5061389  1.279066  -4.093200 1.2422108
## .rate_idOxygen_improving_AA_improving             -1.4304065  1.318944  -4.125794 1.5604030
## .rate_idVentilated_improving_Oxygen_improving     -1.6518820  2.675793  -5.297630 5.9875145
## .rate_idAA_worsening_Oxygen_worsening             -1.7917019  1.336484  -4.826058 0.9650288
## .rate_idOxygen_worsening_Ventilated_worsening     -2.7756560  2.133518  -6.724102 1.8260400
## .rate_idAA_worsening_AA_improving                 -0.5639164  1.533358  -3.963077 2.3096640
## .rate_idOxygen_worsening_Oxygen_improving          2.7492899  2.426670  -1.460351 8.1569808
## .rate_idVentilated_worsening_Ventilated_improving -0.8198872  3.453065  -5.516137 7.0211355
## .rate_idAA_improving_AA_worsening                 -1.9348715  1.561121  -5.164001 1.1503550
## .rate_idOxygen_improving_Oxygen_worsening          0.6799676  2.219280  -3.059431 6.1140482
## .rate_idVentilated_improving_Ventilated_worsening -2.9500644  3.739101 -11.358780 3.7275708
##
## Per rate group effects:
## , , best_supportive_care
##
```

```
##                Estimate Est.Error       Q2.5    Q97.5
## improve_one  0.05167171 0.4679020 -0.7711655 1.089884
## death        2.81535201 0.8007426  1.4213955 4.699739
## to_improving 0.16416284 1.0789955 -1.7097442 2.484940
## to_worsening 1.46385904 0.9847763 -0.2621487 3.605791
## worsen_one   0.57980222 0.7084007 -0.7448406 2.029441
##
## , , took_hcq
##
##                 Estimate Est.Error       Q2.5     Q97.5
## improve_one   0.02438265 0.2069174 -0.3794155 0.4729512
## death        -0.29006237 0.4122813 -1.3059240 0.3242005
## to_improving  0.15462789 0.3428094 -0.4915457 0.9064798
## to_worsening -0.10832369 0.4169333 -0.9414365 0.8450387
## worsen_one   -0.10493831 0.3347226 -0.9547952 0.5174091
##
## , , took_az
##
##                 Estimate Est.Error        Q2.5     Q97.5
## improve_one   0.19636348 0.1870453 -0.09532769 0.6011053
## death         0.04766377 0.2902467 -0.52213820 0.7208174
## to_improving -0.03448963 0.2836839 -0.64793200 0.5264866
## to_worsening  0.08288478 0.3273653 -0.50206905 0.8917839
## worsen_one    0.09662132 0.2683842 -0.41582253 0.7502404
##
## , , took_favipiravir
##
##                Estimate Est.Error       Q2.5     Q97.5
## improve_one   0.3190806 0.3876548 -0.3977633 1.0854492
## death        -0.5960340 1.1415221 -3.5875840 0.9465509
## to_improving -0.0969700 0.7491493 -1.9470315 1.2918705
## to_worsening -0.7583448 1.2212229 -4.0245825 0.6856091
## worsen_one    0.4279498 0.7156565 -0.8094016 2.0709478
##
## , , took_convalescent_plasma
##
##                Estimate Est.Error       Q2.5     Q97.5
## improve_one  -0.2325311 0.3814084 -1.1351070 0.3964487
## death        -0.1877247 0.6225068 -1.6783935 0.8766320
## to_improving -0.1163346 0.6473340 -1.5416787 0.9921753
## to_worsening -0.1120986 0.9653880 -2.6563037 1.3183905
## worsen_one    0.3206859 0.7851961 -0.7141766 2.6062970
##
## , , age_norm
##
##                 Estimate Est.Error       Q2.5       Q97.5
## improve_one  -0.34102734 0.1592352 -0.6270696 0.005550484
## death         0.38694842 0.3849380 -0.1989353 1.321935000
## to_improving -0.05381607 0.2705899 -0.5910787 0.487180775
## to_worsening  0.12557234 0.3443922 -0.5524985 0.900856950
## worsen_one    0.13248513 0.2709348 -0.4853750 0.627241325
##
## , , is_male
##
```

```
##                 Estimate Est.Error       Q2.5      Q97.5
## improve_one  -0.22602846 0.1591376 -0.5501086 0.04174039
## death        -0.02799677 0.2558825 -0.5705421 0.49890673
## to_improving -0.17024993 0.2942298 -0.8820199 0.33260110
## to_worsening -0.02705008 0.2911476 -0.6933474 0.54419480
## worsen_one   -0.11949092 0.2463275 -0.6934125 0.31054940
##
## , , Intercept
##
##                 Estimate Est.Error      Q2.5    Q97.5
## improve_one   0.08010700  1.159288 -2.518968 2.564986
## death        -0.13728195  1.265271 -2.893620 2.561662
## to_improving  0.53281565  1.333640 -1.627321 3.963345
## to_worsening  0.11167609  1.207914 -2.384741 2.948727
## worsen_one    0.04246846  1.186022 -2.518964 2.742138
##
## , , comorbidities_sum
##
##                 Estimate  Est.Error       Q2.5      Q97.5
## improve_one  -0.05854528 0.07316544 -0.2475062 0.04638908
## death         0.05870428 0.11965841 -0.1435329 0.34445100
## to_improving -0.08210750 0.13614937 -0.4216466 0.13138178
## to_worsening -0.10277263 0.19152452 -0.5734875 0.15192008
## worsen_one    0.06360942 0.14427702 -0.1603423 0.41945385
##
##
## Between-site differences:
##                                                        Estimate Est.Error       Q2.5    Q97.5
## sd(.rate_idOxygen_worsening_Death)                    1.2089762 0.7401264 0.09103341 3.033405
## sd(.rate_idVentilated_worsening_Death)                1.3395908 0.8093099 0.08939575 3.189737
## sd(.rate_idAA_improving_Discharged)                   1.3539030 0.5166801 0.48961695 2.534090
## sd(.rate_idOxygen_improving_AA_improving)             0.8231443 0.4617276 0.15817215 1.967023
## sd(.rate_idVentilated_improving_Oxygen_improving)     0.8761621 0.8468631 0.02782630 3.213061
## sd(.rate_idAA_worsening_Oxygen_worsening)             0.9016138 0.6119438 0.04231071 2.316944
## sd(.rate_idOxygen_worsening_Ventilated_worsening)     2.0566520 0.9939326 0.27742428 4.229188
## sd(.rate_idAA_worsening_AA_improving)                 1.2653953 0.7206368 0.12806870 2.938958
## sd(.rate_idOxygen_worsening_Oxygen_improving)         1.0315829 0.7208641 0.05358955 2.762706
## sd(.rate_idVentilated_worsening_Ventilated_improving) 1.0187563 0.8570704 0.02746255 3.157189
## sd(.rate_idAA_improving_AA_worsening)                 1.9562286 0.8793267 0.29771203 3.912508
## sd(.rate_idOxygen_improving_Oxygen_worsening)         0.9370704 0.7089551 0.03710803 2.650372
## sd(.rate_idVentilated_improving_Ventilated_worsening) 1.2847621 1.0398747 0.04405940 3.913308
```

Estimates (not completely trustworthy due to the divergent transitions):

```
##                        hypothesis point_estimate  95% CI Low 95% CI High
## 1                       hcq_death   -0.301638343 -0.90426299  0.06461316
## 2                    hcq_hospital   -0.100708595 -0.68011471  0.45463573
## 3         hcq_hospital (duration)    0.009441702 -0.08968162  0.12892896
## 4                        az_death    0.032465196 -0.28225635  0.43405299
## 5                     az_hospital   -0.245627974 -0.83969611  0.19427970
## 6          az_hospital (duration)   -0.062060872 -0.18824891  0.02419643
## 7               favipiravir_death   -0.610255980 -3.15738941  0.34667968
## 8            favipiravir_hospital   -0.451484421 -1.60800912  0.59286350
## 9 favipiravir_hospital (duration)   -0.099173136 -0.41065741  0.16223596
```

### 6 Joint longitudinal and time-to-event models

Joint models have two components:

- *longitudinal model* describes the evolution of some quantity/quantities over time (in our case a biological marker)
- *time-to-event model* describes the risk of event over time assuming proportional hazards. This risk can be affected by the value of the marker.

Modelling those two jointly means that the marker values are imputed between measurements according to the longitudinal model, but information also flows in the other direction. We use the implemention from the `rstanarm` package (Brilleman et al. 2018) - consult the reference for an introduction to this class of models.

We use those models only for association between markers (IL-6 and D-dimer) and disease progression. Those analyses ended up not being reported in the main paper as the results are inconclusive.

We evaluate the associations directly via the coefficient describing the association between the longitudinal and time-to-event component. We focus solely on the "Death" event.

#### 6.1 D-dimer

##### 6.1.1 Age + sex

The longitudinal formula is `log_d_dimer ~ day + (1  | patient_id)` and the time-to-event formula is `Surv(daym1, day, event_death) ~ age + sex`. The fitted coefficients are:

```
## stan_jm
##  formula (Long1): log_d_dimer ~ day + (1 | patient_id)
##  family  (Long1): gaussian [identity]
##  formula (Event): Surv(daym1, day, event_death) ~ age + sex
##  baseline hazard: bs
##  assoc:           etavalue (Long1)
## ------
##
## Longitudinal submodel: log_d_dimer
##             Median MAD_SD
## (Intercept) 1.171  0.267
## day         0.040  0.005
## sigma       0.502  0.024
##
## Event submodel:
##                 Median  MAD_SD  exp(Median)
## (Intercept)      -9.377   1.769   0.000
## age               0.053   0.025   1.054
## sexM              0.442   0.590   1.555
## Long1|etavalue    0.471   0.264   1.601
## b-splines-coef1 -17.497  10.858      NA
## b-splines-coef2   1.198   1.861      NA
## b-splines-coef3  -1.275   1.141      NA
## b-splines-coef4  -1.070   3.129      NA
## b-splines-coef5  -0.504   8.056      NA
## b-splines-coef6 -23.339  22.178      NA
##
```

```
## Group-level error terms:
##  Groups     Name              Std.Dev.
##  patient_id Long1|(Intercept) 2.538
## Num. levels: patient_id 91
##
## Sample avg. posterior predictive distribution
## of longitudinal outcomes:
##                Median MAD_SD
## Long1|mean_PPD 1.902  0.041
##
## ------
## For info on the priors used see help('prior_summary.stanreg').
```

###### 6.1.2 Age + sex + hydroxychloroquine

The time-to-event formula is expanded to `Surv(daym1, day, event_death) ~ age + sex + took_hcq`. The fitted coefficients are:

```
## stan_jm
##  formula (Long1): log_d_dimer ~ day + (1 | patient_id)
##  family  (Long1): gaussian [identity]
##  formula (Event): Surv(daym1, day, event_death) ~ age + sex + took_hcq
##  baseline hazard: bs
##  assoc:           etavalue (Long1)
## ------
##
## Longitudinal submodel: log_d_dimer
##             Median MAD_SD
## (Intercept) 1.157  0.263
## day         0.041  0.005
## sigma       0.501  0.027
##
## Event submodel:
##                  Median  MAD_SD  exp(Median)
## (Intercept)      -7.663   1.912   0.000
## age               0.038   0.025   1.038
## sexM              0.216   0.627   1.241
## took_hcqTRUE     -1.089   0.644   0.336
## Long1|etavalue    0.557   0.259   1.746
## b-splines-coef1 -17.957  11.497      NA
## b-splines-coef2   1.044   1.948      NA
## b-splines-coef3  -1.397   1.177      NA
## b-splines-coef4  -1.217   2.971      NA
## b-splines-coef5  -0.515   7.633      NA
## b-splines-coef6 -21.450  20.560      NA
##
## Group-level error terms:
##  Groups     Name              Std.Dev.
##  patient_id Long1|(Intercept) 2.529
## Num. levels: patient_id 91
##
## Sample avg. posterior predictive distribution
## of longitudinal outcomes:
```

```
##                 Median MAD_SD
## Long1|mean_PPD 1.904  0.042
##
## ------
## For info on the priors used see help('prior_summary.stanreg').
```

The problems in the model are visible when visualising the fitted longitudinal trajectories for some patients: there are very few measurements and thus huge uncertainties:

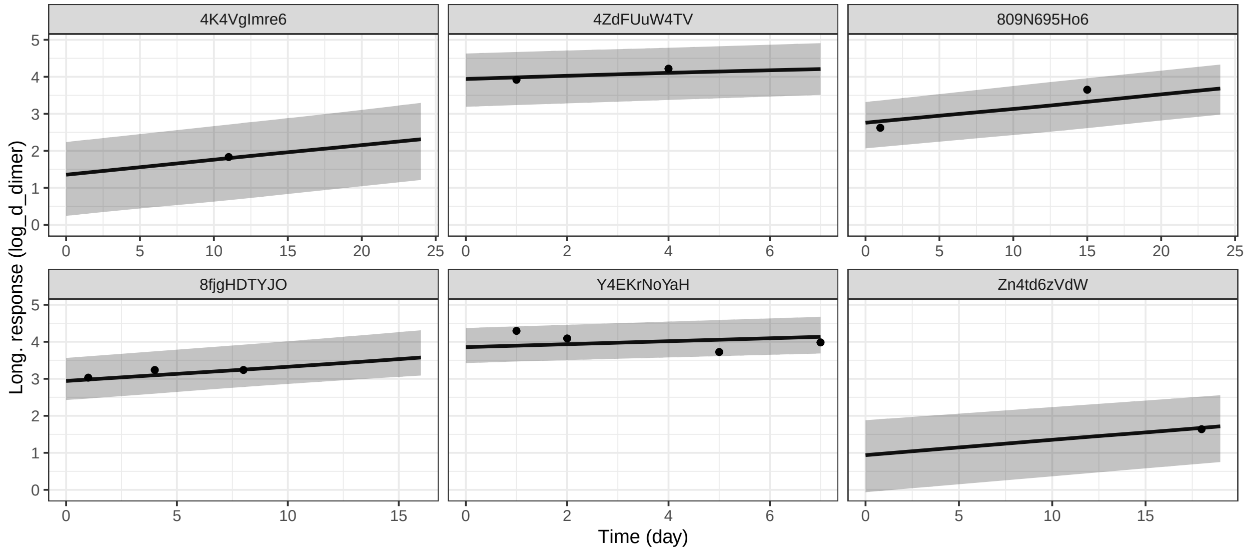

## 6.2 IL-6

##### 6.2.1 Age + sex

The longitudinal formula is `log_IL_6 ~ day + (1 + day | patient_id)` and the time-to-event formula is `Surv(daym1, day, event_death) ~ age + sex`. The fitted coefficients are:

```
## stan_jm
##  formula (Long1): log_IL_6 ~ day + (1 + day | patient_id)
##  family  (Long1): gaussian [identity]
##  formula (Event): Surv(daym1, day, event_death) ~ age + sex
##  baseline hazard: bs
##  assoc:           etavalue (Long1)
## ------
##
## Longitudinal submodel: log_IL_6
##             Median MAD_SD
## (Intercept)  3.978  0.310
## day         -0.047  0.023
## sigma        0.954  0.070
##
## Event submodel:
##                 Median  MAD_SD  exp(Median)
## (Intercept)     -21.804   6.611   0.000
## age               0.171   0.069   1.187
```

```
## sexM              0.308   1.000   1.361
## Long1|etavalue    1.608   0.714   4.995
## b-splines-coef1 -62.087  40.211      NA
## b-splines-coef2   1.396   3.024      NA
## b-splines-coef3 -11.160   4.064      NA
## b-splines-coef4   5.016   6.183      NA
## b-splines-coef5  -8.220  10.505      NA
## b-splines-coef6 -30.435  36.398      NA
##
## Group-level error terms:
##  Groups     Name              Std.Dev. Corr
##  patient_id Long1|(Intercept) 1.86745
##             Long1|day         0.08869  -0.30
## Num. levels: patient_id 50
##
## Sample avg. posterior predictive distribution
## of longitudinal outcomes:
##                Median MAD_SD
## Long1|mean_PPD 4.113  0.103
##
## ------
## For info on the priors used see help('prior_summary.stanreg').
```

###### 6.2.2 Age + sex + hydroxychloroquine

The time-to-event formula becomes `Surv(daym1, day, event_death) ~ age + sex + took_hcq`. The fitted model is:

```
## stan_jm
##  formula (Long1): log_IL_6 ~ day + (1 + day | patient_id)
##  family  (Long1): gaussian [identity]
##  formula (Event): Surv(daym1, day, event_death) ~ age + sex + took_hcq
##  baseline hazard: bs
##  assoc:           etavalue (Long1)
## ------
##
## Longitudinal submodel: log_IL_6
##             Median MAD_SD
## (Intercept)  3.946  0.323
## day         -0.046  0.023
## sigma        0.952  0.070
##
## Event submodel:
##                  Median  MAD_SD  exp(Median)
## (Intercept)     -23.103   7.637   0.000
## age               0.182   0.079   1.200
## sexM              0.227   1.045   1.254
## took_hcqTRUE     -0.051   1.064   0.950
## Long1|etavalue    1.777   0.790   5.910
## b-splines-coef1 -64.309  40.888      NA
## b-splines-coef2   0.854   3.030      NA
## b-splines-coef3 -11.417   4.219      NA
## b-splines-coef4   4.291   5.907      NA
```

```
## b-splines-coef5  -7.232   9.790      NA
## b-splines-coef6 -33.031  37.449      NA
##
## Group-level error terms:
##  Groups     Name              Std.Dev. Corr
##  patient_id Long1|(Intercept) 1.88367
##             Long1|day         0.08822  -0.32
## Num. levels: patient_id 50
##
## Sample avg. posterior predictive distribution
## of longitudinal outcomes:
##                Median MAD_SD
## Long1|mean_PPD 4.109  0.100
##
## ------
## For info on the priors used see help('prior_summary.stanreg').
```

Similarly, we can explore the posterior trajectories, showing similar issues as before: too wide uncertainty.

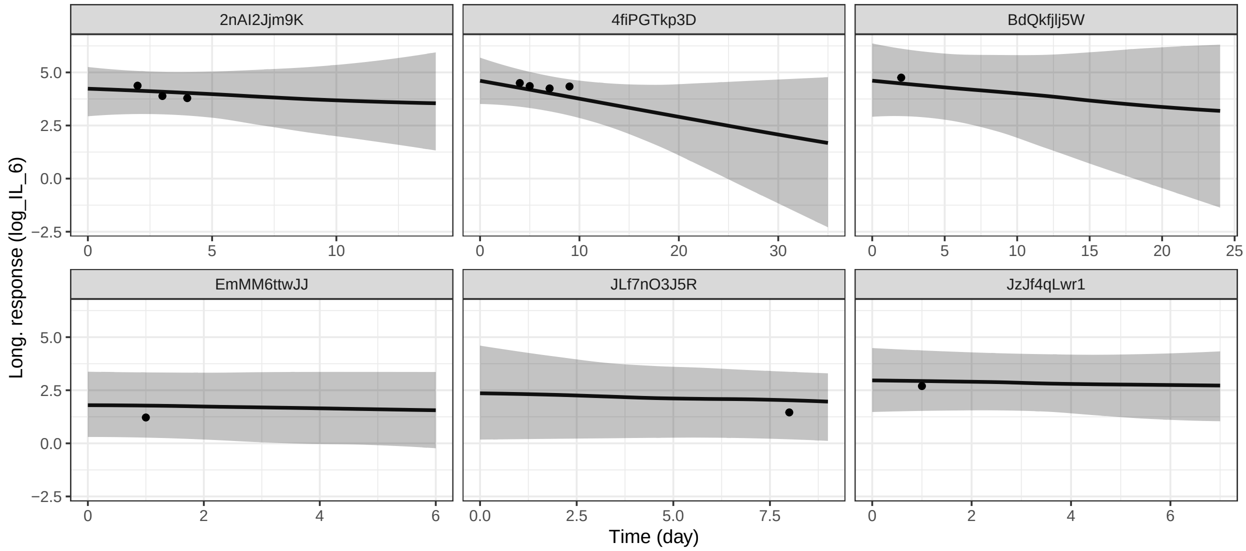

Table 3: Comparing the 12-day mortality based on ACP score (where available) with the 95% confidence intervals (CI) for 12-day mortality reported by Lu et al.

| ACP grade | patients | 12-day mortality | Li et al. 95% CI |
|---|---|---|---|
| 1 | 23 | 0 (0%) | 0% |
| 2 | 67 | 6 (9%) | 0 - 11.3% |
| 3 | 85 | 21 (25%) | 19.8 - 44.3% |

### 7 Validating published prognostic models

In all of the following we focus on the area under curve (AUC), also often called C-statistic. We compute AUC using the `pROC` package (Robin et al. 2011), which also reports bootstrapped confidence intervals.

For all risk scores we look at 12-day mortality and 30-day mortality - simply because those are the time horizons reported in the studies we looked at. When needed or when it is uncertain how the outcome was coded in the original study, we can also look at "indefinite horizon" where we only include patients that were not censored and had a known outcome.

#### 7.1 ACP

The ACP index (Lu et al. 2020) is a simple score that categorizes the patients into 3 categories based on age and C-reactive protein (CRP): grade 1 (age $<$ 60 years and CRP $<$ 34 mg/L), grade 2 (age $\geq$ 60 years and CRP $<$ 34 mg/L or age $<$ 60 years and CRP $\geq$ 34 mg/L), grade 3 (age $\geq$ 60 years and CRP $\geq$ 34 mg/L) and was evaluated primarily on 12-day mortality. Despite the score being developed on patients in Wuhan, China, the 12-day mortality in our dataset is in rough agreement with the values in the original manuscript as shown in Table 3.

The original authors do not report AUC (C-statistic), but they report the overall counts of alive and death patients, letting us compute it. The counts are:

```
## # A tibble: 5 x 3
##   ACP_grade alive_12     N
##       <dbl> <lgl>    <int>
## 1         1 TRUE       194
## 2         2 FALSE        8
## 3         2 TRUE       137
## 4         3 FALSE       32
## 5         3 TRUE        63
```

Giving us this AUC:

| AUC | 95% CI |
|---|---|
| 0.869 | (0.831, 0.907) |

So we can now compare AUC computed on our data for both 12-day mortality (as in the original study) and 30-day mortality.

We see that the score noticeably underperforms in the present dataset.

#### 7.2 Chen and Liu

Chen and Liu (Chen and Liu 2020) propose a score based on a logistic regression model as follows.

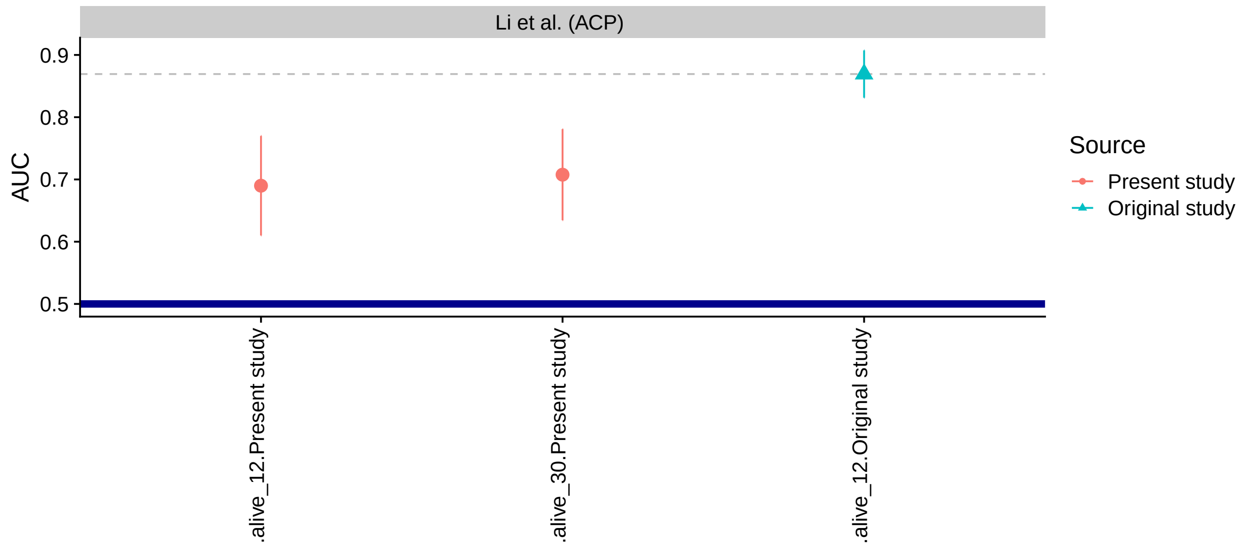

Figure 2: The AUC and it's 95% credible intervals for the ACP score. The thick blue line marks AUC of 0.5, i.e. the point where reverting the score would yield better predictions. The thin gray dashed line marks the AUC from the original study for easier comparison.

$$Score = -4.211 + 0.013 \times CRP + 0.059 \times Age + 0.112 \times DD - 1.984 \times LYM$$

Where CRP is C-reactive protein (mg/l) DD is D-dimer (the paper does not have units, but the associated web app is clear that it expects D-dimer as ug/ml FEU) and LYM is the Lympohocyte count (10^9 / L). The paper states:

> patients who were with >10% missing values, stayed in the hospital < 7 days, afflicted by a severe disease before admission (e.g., cancer, aplastic anemia, and uremia), were unconscious at admission, and were directly admitted to the intensive care unit (ICU) were excluded.

> Non-normally distributed continuous variables were transformed using a Box-Cox transformation.

However, reverse engineering the asociated web app (https://phenomics.fudan.edu.cn/risk_scores/), no transform appears to have been done to the variables that were finally selected. It is also slightly unclear how they computed the outcome (i.e. how they treated censoring).

The units in our dataset are:

```
## # A tibble: 3 x 2
##   marker          unit
##   <chr>           <chr>
## 1 CRP             mg/l
## 2 lymphocyte_count 10^9/l
## 3 d_dimer         ng/ml DDU
```

Which matches the model except for D-dimer (we use ng/ml DDU, FEU value is double the DDU).

A plot of the computed values for our dataset.

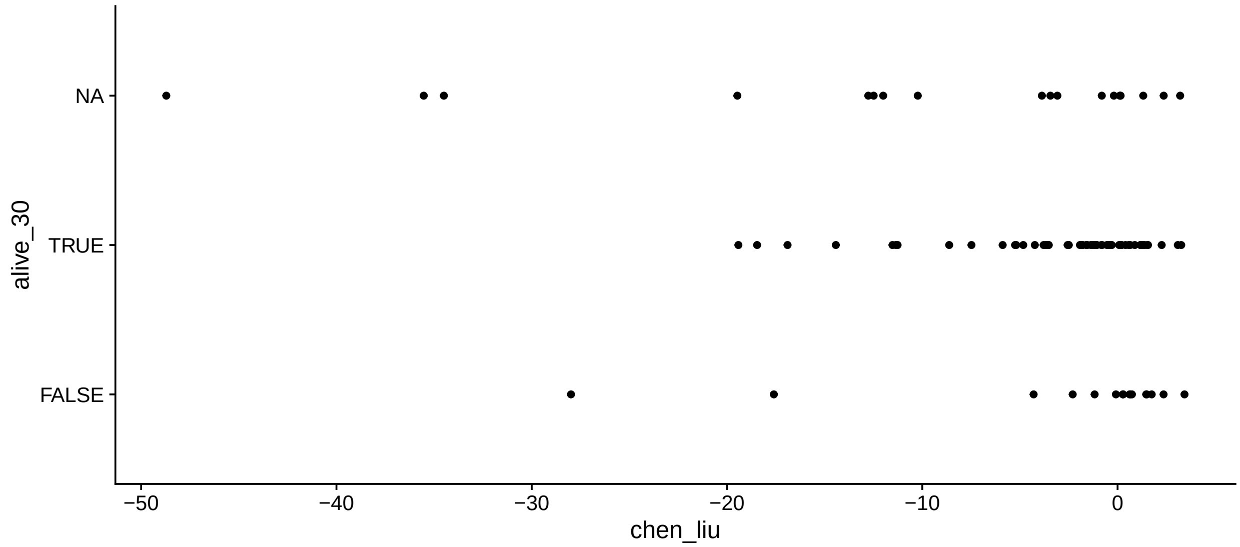

We look at survival since first day when all of the values required for the score are available, or we ignore when the values were computed and look at survival since admission. Finally we also tried emulating the inclusion criteria from Chen & Lie

Chen & Liu report AUC = 0.881 for the validation dataset and don't provide enough information to let us recompute it with credible intervals.

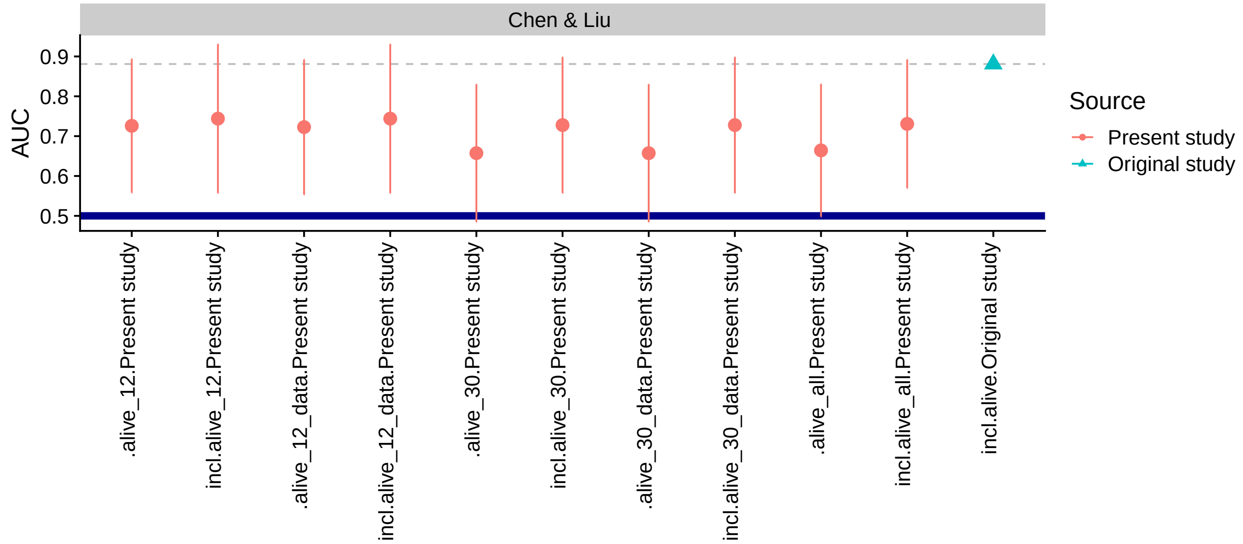

Figure 3: The AUC and it's 95% credible intervals for the Chen & Liu score. The thick blue line marks AUC of 0.5, i.e. the point where reverting the score would yield better predictions. The thin gray dashed line marks the AUC from the original study for easier comparison. The outcomes ending with `_data` mean mortality was counted since the first day all the lab markers were measured, otherwise it is since hospitalization. `alive_all` mens that only patients that were not censored were included.

##### 7.3 Shi et al.

Shi et al. (Shi et al. 2020) report a simple score: 1 point for Age >= 50, Male and Hypertension each. They try to predict "severe" illness, but it is not clear what "severe" means. Here, we test both "not ventilated" (or worse, including death) and “not needing supplementary oxygen” as proxy for “severe”. They also create a risk score for both “severe” on admission and “severe” during hospitalization. As for other scores we also look at 12- and 30- day mortality.

We extract the original data from Fig1 B,C using WebPlotDigitizer.

| score | mild_cases_admission | all_cases_admission | mild_cases_hospitalization | all_cases_hospitalization |
|---|---|---|---|---|
| 0 | 118 | 118 | 11 | 12 |
| 1 | 199 | 211 | 25 | 29 |
| 2 | 98 | 121 | 11 | 18 |
| 3 | 21 | 35 | 4 | 7 |

Here is the comparison to original data.

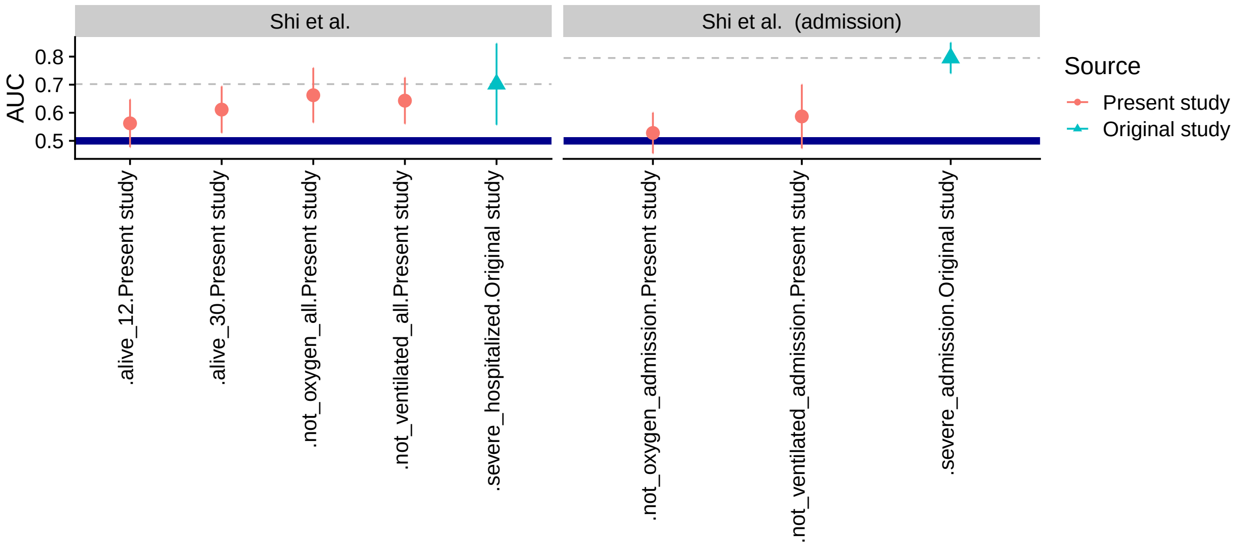

Figure 4: The AUC and it’s 95% credible intervals for the Shi et al. score. The thick blue line marks AUC of 0.5, i.e. the point where reverting the score would yield better predictions. The thin gray dashed line marks the AUC from the original study for easier comparison.

We see that the score consistently underperforms, although some of the CIs are quite wide.

We can also compare the ratios of “severe” cases reported by Shi et al. to ratios in our dataset.

| score | Shi et al. (admission) | Ventilated (admission) | Oxygen (admission) | N |
|---|---|---|---|---|
| 0 | 0.000 | 0.0000000 | 0.3636364 | 11 |
| 1 | 0.057 | 0.1052632 | 0.6052632 | 38 |
| 2 | 0.190 | 0.0970874 | 0.5436893 | 103 |
| 3 | 0.400 | 0.1639344 | 0.6065574 | 61 |

| score | Shi et al. (hospitalization) | Ventilated (hospitalization) | Oxygen (hospitalization) | N |
|---|---|---|---|---|
| 0 | 0.083 | 0.1000000 | 0.1666667 | 10 |
| 1 | 0.138 | 0.0645161 | 0.1538462 | 31 |
| 2 | 0.389 | 0.3205128 | 0.4807692 | 78 |
| 3 | 0.429 | 0.3684211 | 0.6363636 | 38 |

#### 7.4 Caramelo et al.

Caramelo et al. (Caramelo, Ferreira, and Oliveiros 2020) uses simulations to get patient-level characterstics from aggregate data and then fit a logistic regression model. We use the coefficients they report for the model (the model can't be used directly, because the intercept is not reported). The covariates in the model are age (by decades), sex, presence of hypertension, presence of diabetes, presence of cardiac disease, presence of chronic respiratory disease and presence of cancer. It is unclear whether NYHA > 1 would be considered "cardiac disease" and similarly whether asthma is chronic respiratory disease, so we compute two versions of the score. Our dataset also does not contain cancer, so we ignore it (cancer has one of the lowest reported OR in the model).

Caramelo et al. have not computed AUC in their paper and do not give us any mean to reconstruct it, so we only present our results for the risk score.

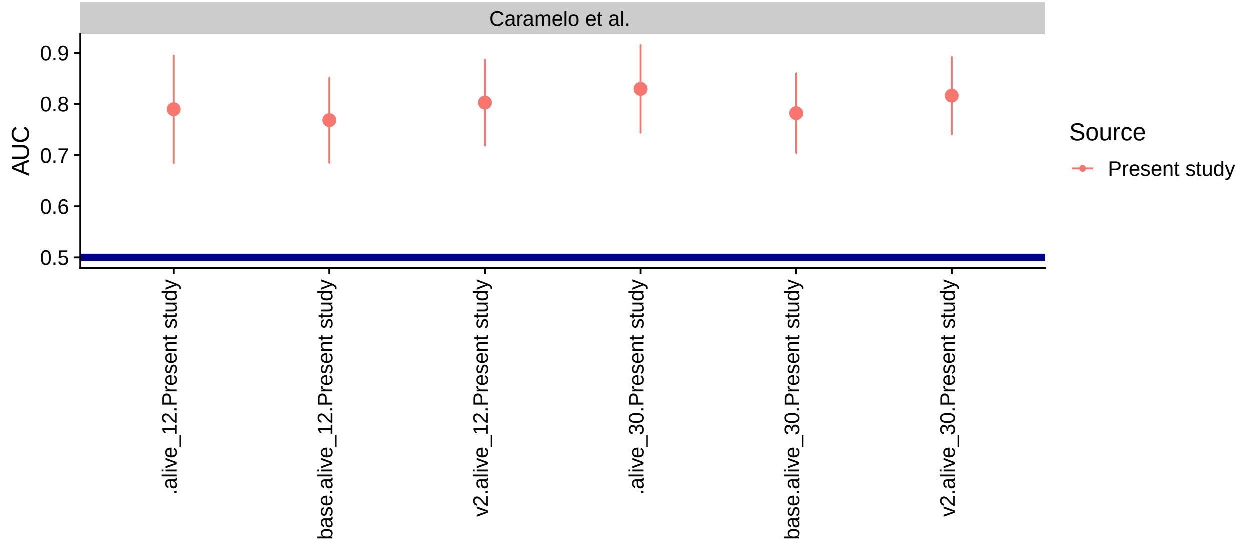

Figure 5: The AUC and it's 95% credible intervals for the Caramelo et al. score. The thick blue line marks AUC of 0.5, i.e. the point where reverting the score would yield better predictions.

The results look somewhat encouraging for the score - the AUC values float around 0.8.

#### 7.5 Bello-Chavolla et al.

The index given in (Bello-Chavolla et al. 2020) is built from following elements:

```
##                      factor score
## 1          Age >= 65 years     3
## 2                 Diabetes     1
## 3 Diabetes*Age < 40 years     5
## 4           Age < 40 years    -6
## 5                  Obesity     1
## 6                Pneumonia     7
## 7  Chronic Kidney Disease     3
## 8                     COPD     1
## 9        Immunosuppression     1
```

Of those, our data does not contain Immunosuppresion and Pneumonia. We can however use taking antibiotics within 1 day of hospitalization as an imperfect proxy for pneumonia. We will ignore immunosuppresion in our test. Both of those choices could reduce the performance of the score.

The authors do not define obesity, so we will use BMI > 30 as threshold for obesity. Our data contains an indicator for "Renal disease" which might be classified slightly differently than "Chronic Kidney Disease".

The authors report C-statistic = 0.830 for mortality on validation dataset - it is unclear how they treat censoring, but they mention 30-day mortality in other part of the paper, so we will focus on 30-day mortality.

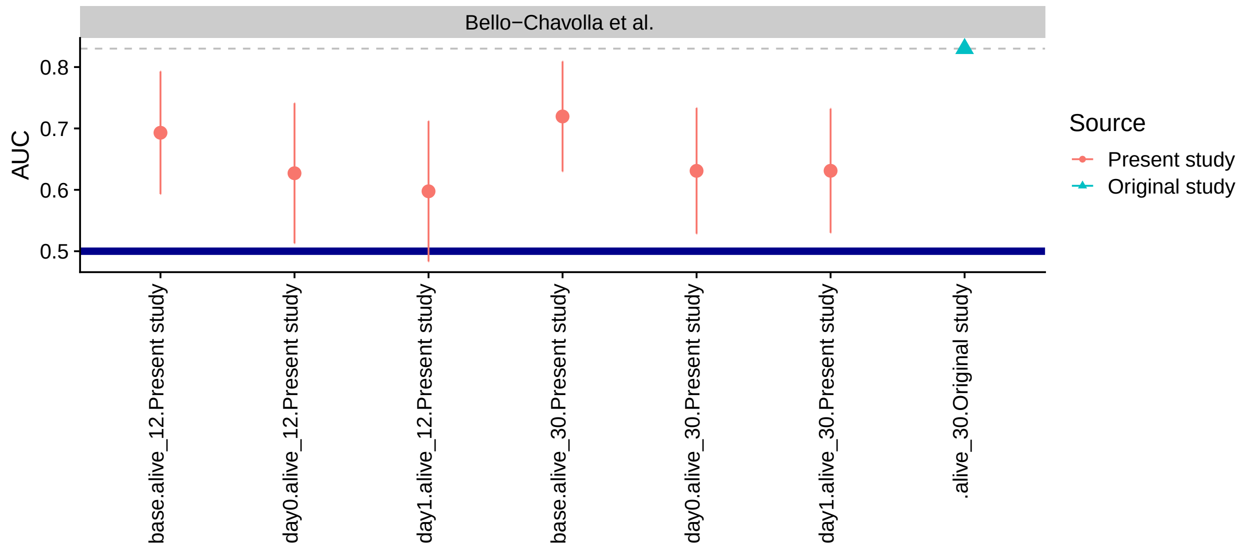

Figure 6: The AUC and it's 95% credible intervals for the Bello-Chavolla et al. score. The thick blue line marks AUC of 0.5, i.e. the point where reverting the score would yield better predictions. The thin gray dashed line marks the AUC from the original study for easier comparison. We use 3 variants of the score - `base` means not inferring pneumonia at all, `day0` means that pneumonia is inferred from being admitted for covid and antibiotics at hospitalization, `day1` - inferring pneumonia from antibiotics within 1 day of hospitalization

The main surprise here is that not inferring pneumonia (shown as "base" in the figure) works best. This indicates that our data may not be well suited to the use of the score.

##### 7.6 Age only

As a simple baseline we use age in years and the decade of age as the sole predictor and compute AUC for this.

| auc | auc_low | auc_high | outcome | score | note | subgroup |
|---|---|---|---|---|---|---|
| 0.8240157 | 0.7496651 | 0.8983664 | alive_30 | age | | |
| 0.8285019 | 0.7502675 | 0.9067362 | alive_12 | age | | |
| 0.7950787 | 0.7200672 | 0.8700903 | alive_30 | age_decade | decade | |
| 0.7945269 | 0.7137577 | 0.8752961 | alive_12 | age_decade | decade | |

##### 7.7 Summary

We see that the "age only" baseline performs similarly to the best performing score (Caramelo et al.) and likely better than all of the other scores - there is however some uncertainty prohibiting us to be very sure

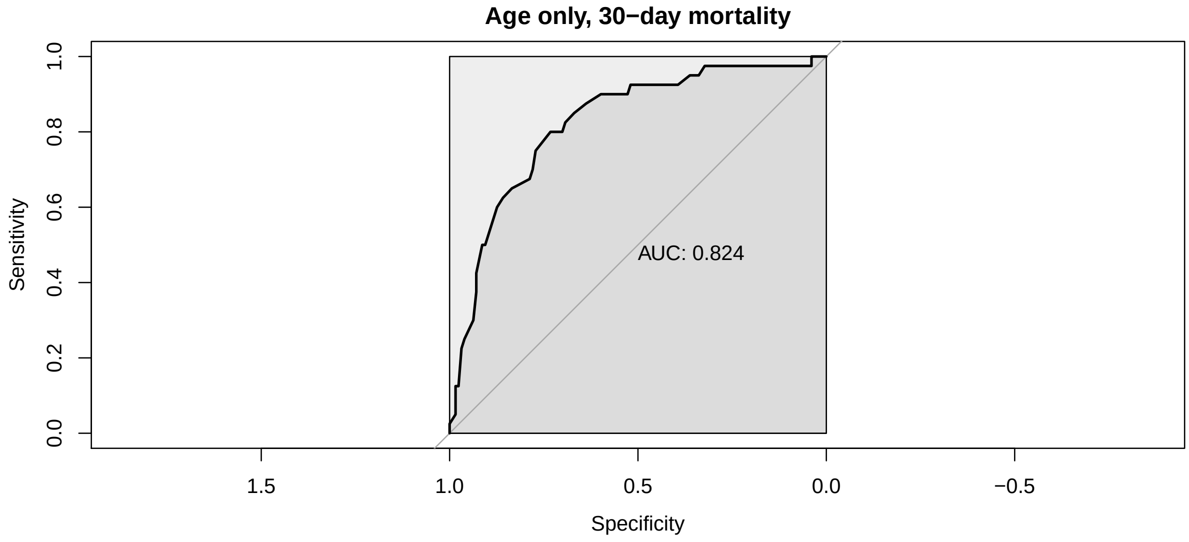

Figure 7: The ROC curve for age in years as the sole predictor of 30-day mortality.

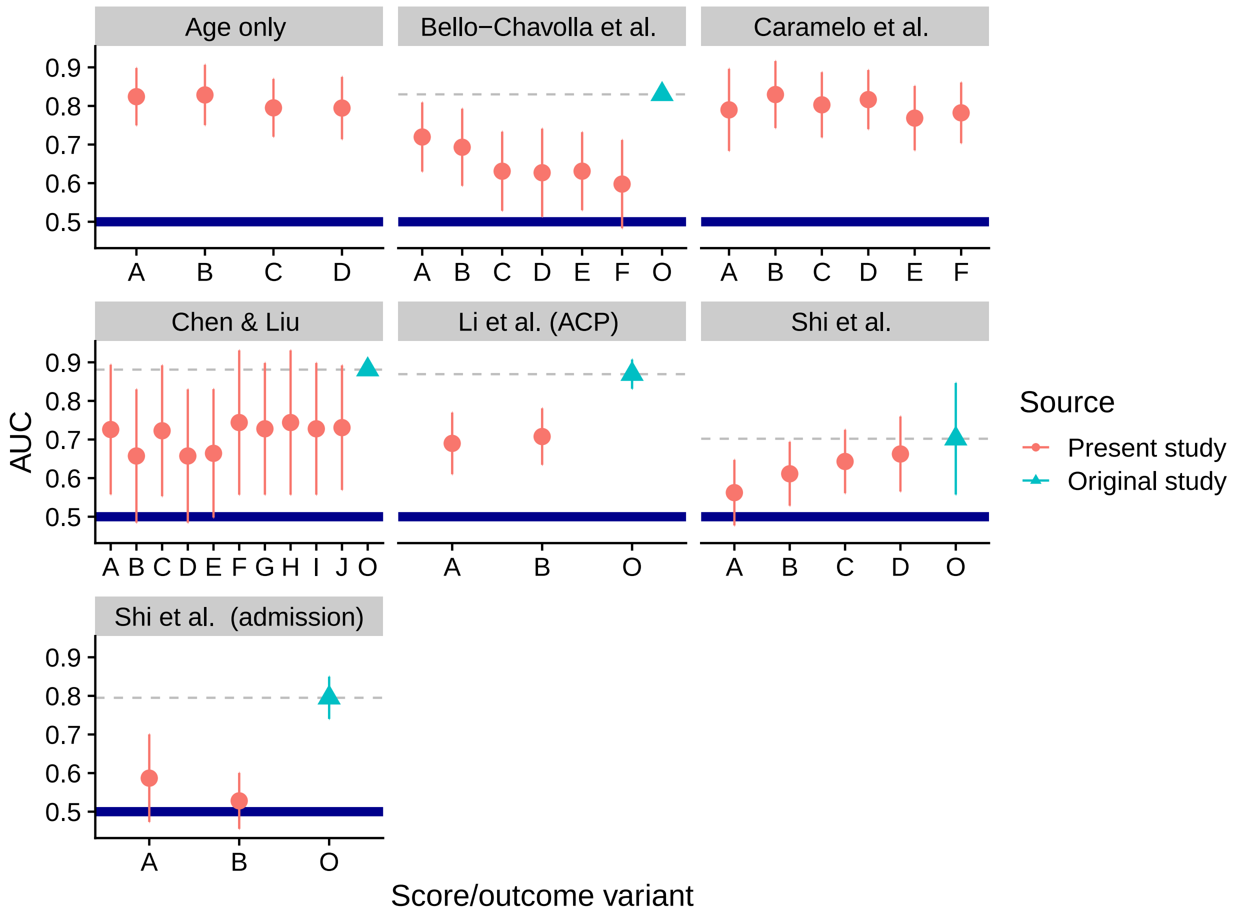

Figure 8: The AUC and it's 95% credible intervals for all of the scores considered so far. The thick blue line marks AUC of 0.5, i.e. the point where reverting the score would yield better predictions. The thin gray dashed line marks the AUC from the original study for easier comparison.

about the ordering. This holds even if we only take the decade of age. Note that the score by (Caramelo et al.) is actually heavily based on the decade of age. Table of the results is shown below as the small differences are not well visible in the plot.

| score | subgroup | note | best_auc |
|---|---|---|---|
| Caramelo et al. | | | 0.8295238 |
| Age only | | | 0.8285019 |
| Caramelo et al. | | v2 | 0.8164370 |
| Age only | | decade | 0.7950787 |
| Caramelo et al. | | base | 0.7821850 |
| Chen & Liu | | incl | 0.7439394 |
| Chen & Liu | | | 0.7258454 |
| Bello-Chavolla et al. | | base | 0.7195122 |
| Li et al. (ACP) | | | 0.7076380 |
| Shi et al. | | | 0.6624650 |
| Bello-Chavolla et al. | | day1 | 0.6310378 |
| Bello-Chavolla et al. | | day0 | 0.6309182 |
| Shi et al. | (admission) | | 0.5868607 |

### 8 Original computing environment

```
## R version 4.0.3 (2020-10-10)
## Platform: x86_64-w64-mingw32/x64 (64-bit)
## Running under: Windows 10 x64 (build 18363)
##
## Matrix products: default
##
## locale:
## [1] LC_COLLATE=Slovak_Slovakia.1250  LC_CTYPE=Slovak_Slovakia.1250    LC_MONETARY=Slovak_Slovakia.125
## [5] LC_TIME=Slovak_Slovakia.1250
##
## attached base packages:
## [1] stats     graphics  grDevices utils     datasets  methods   base
##
## other attached packages:
##  [1] patchwork_1.0.1                  covid19retrospective_0.0.0.9000 rstanarm_2.21.1
##  [5] Rcpp_1.0.5                       tidybayes_2.3.1                 survival_3.2-7
##  [9] stringr_1.4.0                    dplyr_1.0.2                     purrr_0.3.4
## [13] tidyr_1.1.2                      tibble_3.0.4                    ggplot2_3.3.2
## [17] testthat_2.3.2
##
## loaded via a namespace (and not attached):
##   [1] readxl_1.3.1         backports_1.1.10     RcppEigen_0.3.3.7.0  plyr_1.8.6           igraph_1.2
##   [8] operator.tools_1.6.3 crosstalk_1.1.0.1    usethis_1.6.3        rstantools_2.1.1     inline_0.3
##  [15] rsconnect_0.8.16     fansi_0.4.1          magrittr_1.5         memoise_1.1.0        remotes_2.
##  [22] matrixStats_0.57.0   formula.tools_1.7.1  MCMCpack_1.4-9       xts_0.12.1           prettyunit
##  [29] blob_1.2.1           ggdist_2.3.0         haven_2.3.1          xfun_0.19            callr_3.5.
##  [36] lme4_1.1-25          zoo_1.8-8            glue_1.4.2           gtable_0.3.0         MatrixMode
##  [43] pkgbuild_1.1.0       rstan_2.21.2         abind_1.4-5          SparseM_1.78         scales_1.1
##  [50] miniUI_0.1.1.1       xtable_1.8-4         stats4_4.0.3         splines2_0.3.1       StanHeader
##  [57] htmlwidgets_1.5.2    threejs_0.3.3        arrayhelpers_1.1-0   ellipsis_0.3.1       farver_2.0
##  [64] dbplyr_1.4.4         utf8_1.1.4           janitor_2.0.1        here_0.1             labeling_0
```

```
##  [71] reshape2_1.4.4      later_1.1.0.1       munsell_0.5.0       cellranger_1.1.0    tools_4.0.3
##  [78] devtools_2.3.2      broom_0.7.2         ggridges_0.5.2      evaluate_0.14       fastmap_1.0
##  [85] processx_3.4.4      knitr_1.30          fs_1.5.0            nlme_3.1-149        mime_0.9
##  [92] compiler_4.0.3      bayesplot_1.7.2     shinythemes_1.1.2   rstudioapi_0.11     curl_4.3
##  [99] stringi_1.5.3       highr_0.8           ps_1.4.0            desc_1.2.0          Brobdingnag
## [106] nloptr_1.2.2.2      markdown_1.1        shinyjs_2.0.0       vctrs_0.3.4         pillar_1.4
## [113] data.table_1.13.2   cowplot_1.1.0       conquer_1.0.2       httpuv_1.5.4        R6_2.4.1
## [120] gridExtra_2.3       sessioninfo_1.1.1   codetools_0.2-16    boot_1.3-25         colourpicke
## [127] assertthat_0.2.1    pkgload_1.1.0       rprojroot_1.3-2     withr_2.3.0         shinystan_
## [134] parallel_4.0.3      hms_0.5.3           grid_4.0.3          minqa_1.2.4         coda_0.19-
## [141] snakecase_0.11.0    pROC_1.16.2         shiny_1.5.0         lubridate_1.7.9     base64enc_
```
