## Supplementary material for "Detailed disease progression of 213 patients hospitalized with Covid-19 in the Czech Republic: An exploratory analysis": Data Collection Protocol

### Protocol for the CovidRetro observational study

Version 1.4, 5. 6. 2020

A retrospective observational study - only data already entered in the patient's documentation will be used. No additional examinations, tests or treatments will be administered beyond those already provided as a part of standard care.

### Inclusion criteria

All patients admitted to inpatient hospital care who tested PCR positive for SARS-CoV2 will be included **except** for patients who:

- Are participating in a clinical trial of a drug for Covid-19
- Are less than 18 years old.

If it is for some reason impossible to collect data from all patients admitted in the given time window, please contact the study coordinator.

### Main principles of data collection

- There are two main modes of data collection: *centralized* for the whole site or *per department*. Centralized collection is preferred if it is feasible.
    - In centralized data collection, all data shall be entered into a single copy of the attached Excel spreadsheet "Data_Collection.xlsx", tracking each patient's history across all departments they were admitted to.
    - In per department data collection, each department fills in a separate copy of the "Data_Collection.xlsx" sheet. When a patient is transferred between departments, they have a separate record in each department. This method should be only used if centralized data collection is not practical.
- Only cells within thick green borders should be edited.
- The spreadsheet will be processed automatically, please do not alter its structure.
- All dates should be entered in the YYYY-MM-DD format (e.g. 2020-04-28)
- Values outside detection limits should be entered with "<" or ">" respectively, e.g. ">2000", "<1"
- If data is not available, leave the cell empty or, if more clarity is desired, explicitly fill in "NA"
- Whenever a "Yes" or "No" answer is applicable, you can also enter just "Y" or "N".
- All sheets contain example data to show the desired structure of collected information.

### Site data

- All coauthors should enter an email to let us contact them directly to comment on drafts of the manuscript and to enter them into journal submission systems. Other than that, the emails will not be shared or used.
- Please list the ORCID number, if the coauthors have one.
- Conflict of interest has to be declared - fill “None” if you don’t have any.

### Patient data

- For the purposes of the study, patients within each site are indexed by an ID starting from 1. Due to anonymization requirements, the site should not store the mapping between patient ID and individual patients.
- **Please do not enter any personal identifying information in the table.**
- The sheet contains example data and empty patient records. Example patient data should be kept in the spreadsheet for easier updating of the data.
- Each row represents a single patient
- Details for individual columns:
    - **Basic information:**
        - **Age:** Age in years
        - **Sex**: For the purposes of this study, sex should - to the best of your knowledge - correspond to karyotype.
        - **Date of symptom onset:** Date when the patient or their carer subjectively first noticed any symptoms associated with Covid-19. If unknown, set to “NA”.
        - **Date of admission:** Date of first admission to the hospital (if collecting data centrally per site) or of first admission to the department (if collecting data per department)
        - **Transferred from:** If the patient was admitted to the site/department after being transferred from elsewhere, please identify the site/department they were hospitalized previously. Otherwise leave empty.
        - **Covid Medications before admission:** If the patient took any specifically anti-Covid medications (Azithromycin, Hydroxychloroquine, Kaletra, Tocilizumab, Favipiravir, Interferon alpha, …) list them. Enter “No” only if it is known that the patient didn’t take any such medications. If the information is not available, please enter “NA”
        - **Admitted for Covid:** “Yes” - The patient was first admitted to a hospital in connection with Covid-19, “No” - they were first hospitalized for other reasons and Covid infection was discovered incidentally.
    - **Medical history:**
        - **Ischemic heart disease:** “Yes” or “No”

- **Hypertension drugs:** "No" if the patient is not diagnosed with hypertension or does not require any anti-hypertensive drugs, otherwise the number of different anti-hypertensive drugs the patient uses regularly.
- **Heart failure:** "Yes" or "No"
- **COPD:** Chronic obstructive pulmonary disease. "Yes" or "No"
- **Asthma Bronchiale:** "Yes" or "No"
- **Other lung disease:** Any other lung disease excluding COPD and Asthma. "Yes" or "No"
- **Diabetes** diagnosed with diabetes of any type, "Yes" or "No"
- **Renal Disease:** "Yes" or "No"
- **Liver Disease:** "Yes" or "No"
- **Smoking:** "Yes" or "No"

- **Clinical status on admission:** All data should be entered as evaluated upon first hospital admission, if not available enter "NA".
  - **BMI:** The body mass index, up to one decimal digit.
  - **NYHA:** New York Heart Association score for heart failure, if available or deducible from documentation ("NA" otherwise). The score has 4 levels:
    - 1: No limitation of physical activity. Ordinary physical activity does not cause undue fatigue, palpitation, dyspnea
    - 2: Slight limitation of physical activity. Comfortable at rest. Ordinary physical activity results in fatigue, palpitation, dyspnea.
    - 3: Marked limitation of physical activity. Comfortable at rest. Less than ordinary activity causes fatigue, palpitation, or dyspnea.
    - 4: Unable to carry on any physical activity without discomfort. Symptoms of heart failure at rest. If any physical activity is undertaken, discomfort increases.
  - **Creatinin:** Concentration of creatinine in serum (μmol/L)
  - **PT INR (Quick):** Prothrombin time (Quick test) as International Normalized Ratio, enter as ratio (NOT as percent).
  - **Albumin:** Concentration of albumin in serum/plasma (g/l)
- **Outcomes:**
  - **Discontinued medication:** Consider only medication that was used primarily to address Covid-19, do not include medication for any underlying comorbidities of the patient. Enter "No" if the patient used no anti-Covid drugs or if all drugs were administered for their planned course. If use of a drug was discontinued, enter "Yes" and briefly describe which drug and why. Especially mention any adverse effects of the medication if they led to discontinuation.
  - **Best supportive care from:** If the patient was determined to be too frail for some treatments (e.g. mechanical ventilation), enter the first day when treatment that would otherwise be chosen was avoided and best supportive care was initiated.

    - **Date of last record:** Last date for which patient data are available in the hospital/department.
    - **Outcome:** The final outcome as of the day of data collection. One of "Discharged", "Hospitalized", "Transferred" (when transferred to a different hospital) and "Death"
    - **Transferred to:** If the outcome is "Transferred" where was the patient transferred to?
  - **Note:** Any other information deemed relevant for the study.

### Disease progression

- This sheet collects details on disease progression. It is the most complex part of data entry, but we can make it together. If in doubt on how to fill it in, contact the study coordinator, we will gladly guide you through any issues you have. A short video showing the process of entering a patient is available at
- There are pre-filled headers for each patient, the "Patient ID" corresponds to "Patient ID" on the "Patients" sheet, please make sure you are entering data for the correct patient.
- Multiple rows correspond to each patient, columns correspond to days of disease
- Start by filling the "First_Day" for a patient, this is the first date for which you have any data about the patient, it should not be later than the date of hospital admission. If data are known after symptom onset and before hospitalization, please enter the date of the first event as "First_Day" (e.g. if a PCR test collected on 2020-04-01 was positive and the patient was admitted on 2020-04-04, enter "2020-04-01" as first day)
- After filling "First_Day" the "Date" row for each patient will update to show dates from the first day onward. This is only for easier data entry.
- Continue by filling in each "Indicator" for each day it is available for the patient. We expect that most indicators will not be available for most days. Leave cells with no data empty or enter "NA" to indicate you are sure the data is not available (instead of say forgetting to enter the data)
- Details for individual indicators
  - **Breathing:** One of:
    - "AA" - Ambient air, no breathing support required
    - "Oxygen" - Supplemental oxygen is provided. If yes, please also fill "Supp. O2" below
    - "NIPPV" - Noninvasive positive-pressure ventilation
    - "MV" - Mechanical ventilation
    - "ECMO" - Extracorporeal membrane oxygenation
  - **Supp. O2**: If supplementary oxygen is provided, what is the flow (litres/minute)?
  - **SpO2:** peripheral capillary oxygen saturation, in percent, measured with the breathing support indicated above

- **SpO2 native**: peripheral capillary oxygen saturation, in percent, measured without any breathing support/oxygen. Enter “NT” if absence of breathing support/oxygen is not tolerated by the patient.
- **Horowitz index**: $PaO_2/FiO_2$, relevant only for patients on MV.
- **PEEP** Applied positive end-expiratory pressure, in $cmH_2O$. Relevant only for patients on MV.
- **Laboratory results**
    - All tests (especially PCR) should be entered for the day the biological samples were collected, not the day the results arrived.
    - **PCR:** Result of a PCR test for SARS-CoV2. If possible, enter the Ct number measured in the test. Otherwise, enter “NEG” for negative and “POS” for positive.
    - **Ferritin, D-dimer, CRP, IL-6:** Ferritin, D-dimer, C-reactive protein and Interleukin 6 levels in blood/serum/plasma.
    - **Lymphocyte count** number of lymphocytes in blood (10^9/l)
    - If you measure some of the indicators in other units than shown, you can either transform to the units shown (this is preferable) or you can simply change the units.

- **Adverse events:** Any adverse events observed for the day, both potentially linked to medication and without a visible link. Events that should definitely be recorded are:
    - Diarrhea
    - Renal failure
    - Blurred vision
    - Chest pain or angina
    - Venous thromboembolic (pulmonary embolism and deep vein thrombosis) events
    - Acute renal failure
    - End stage renal disease
    - Hepatic failure
    - Acute pancreatitis
    - Heart failure
    - Acute myocardial infarction
    - Sudden cardiac death
    - Cardiac arrhythmia
    - Bradycardia
    - Gastrointestinal bleeding
    - Transient ischemic attack
    - Stroke (ischemic or hemorrhagic)
- **Other markers** If you routinely measure other markers and consider them informative of disease progression (and have time to enter them), feel free to enter them into one of the empty rows.

- **Drugs**
    - Enter dosage per day, one drug per row. You can remove drugs the patient never received and add others that were used. Please include only drugs specifically targeting Covid-19 or it's symptoms. This includes experimental therapies (Hydroxychloroquine, Azithromycin, Kaletra, Remdesivir, Favipiravir, Tocilizumab, ...) and any antibiotics used against bacterial superinfection. Please do not include drugs targeting underlying conditions of the patient not related to Covid-19 (e.g. antihypertensive).

### Care

- To make interpretation of data easier and to allow comparison between study sites, please briefly summarise how application of experimental drugs is decided and few other details about your site.
